# Dietary amino acids and risk of stroke subtypes: a prospective analysis of 356,000 participants in seven European countries

**DOI:** 10.1101/2023.08.15.23294122

**Authors:** Tammy Y.N. Tong, Robert Clarke, Julie A. Schmidt, Inge Huybrechts, Urwah Noor, Nita G. Forouhi, Fumiaki Imamura, Ruth C. Travis, Elisabete Weiderpass, Krasimira Aleksandrova, Christina C. Dahm, Yvonne T. van der Schouw, Kim Overvad, Cecilie Kyrø, Anne Tjønneland, Rudolf Kaaks, Verena Katzke, Catarina Schiborn, Matthias B. Schulze, Ana-Lucia Mayen-Chacon, Giovanna Masala, Sabina Sieri, Maria Santucci de Magistris, Rosario Tumino, Carlotta Sacerdote, Jolanda M.A. Boer, W.M. Monique Verschuren, Magritt Brustad, Therese Haugdahl Nøst, Marta Crous-Bou, Dafina Petrova, Pilar Amiano, José María Huerta, Conchi Moreno-Iribas, Gunnar Engström, Olle Melander, Kristina Johansson, Kristina Lindvall, Elom K. Aglago, Alicia K. Heath, Adam S. Butterworth, John Danesh, Timothy J. Key

**Affiliations:** Cancer Epidemiology Unit, Nuffield Department of Population Health, University of Oxford, Oxford, United Kingdom; Clinical Trial Service Unit and Epidemiological Studies Unit, Nuffield Department of Population Health, Oxford, United Kingdom; Department of Clinical Epidemiology, Department of Clinical Medicine, Aarhus University Hospital and Aarhus University, Aarhus, Denmark; Nutrition and Metabolism Branch, International Agency for Research on Cancer (IARC), World Health Organization (WHO), Lyon, France; MRC Epidemiology Unit, University of Cambridge School of Clinical Medicine, Cambridge, United Kingdom; International Agency for Research on Cancer (IARC), World Health Organization (WHO), Lyon, France; Department Epidemiological Methods and Etiological Research, Leibniz Institute for Prevention Research and Epidemiology - BIPS, Bremen, Germany; Faculty of Human and Health Sciences, University of Bremen, Grazer Straße 2, 28359, Bremen, Germany; Department of Public Health, Aarhus University, Aarhus, Denmark; Julius Center for Health Sciences and Primary Care, University Medical Center Utrecht, Utrecht University, Utrecht, The Netherlands; Danish Cancer Society Research Center, Copenhagen, Denmark; Division of Cancer Epidemiology, German Cancer Research Center (DKFZ), Heidelberg, Germany; Department of Molecular Epidemiology, German Institute of Human Nutrition Potsdam-Rehbruecke, Nuthetal, Germany; German Center for Diabetes Research (DZD), Neuherberg, Germany; Institute for Nutritional Science, University of Potsdam, Nuthetal, Germany; Institute for cancer research, prevention and clinical network (ISPRO), Florence, Italy; Epidemiology and Prevention Unit, Fondazione IRCCS Istituto Nazionale dei Tumori di Milano, Milan, Italy; Azienda Ospedaliera Universitaria Federico II, Napoli, Italy; Hyblean Association for Epidemiological Research AIRE-ONLUS, Ragusa, Italy; Unit of Cancer Epidemiology, Città della Salute e della Scienza University-Hospital, Turin, Italy; National Institute for Public Health and the Environment, Bilthoven, the Netherlands; Department of Community Medicine, Faculty of Health Sciences, UiT The Arctic University of Norway, Tromsø, Norway; The Public Dental Service Competence Centre of Northern Norway (TkNN), Tromsø, Norway; K.G. Jebsen Centre for Genetic Epidemiology, Department of Public Health and Nursing, Norwegian University of Science and Technology, Trondheim, Norway; Unit of Nutrition and Cancer, Cancer Epidemiology Research Program, Catalan Institute of Oncology (ICO) - Bellvitge Biomedical Research Institute (IDIBELL). L’Hospitalet de Llobregat, Barcelona, Spain; Department of Epidemiology, Harvard T.H. Chan School of Public Health. Boston, MA, USA; Escuela Andaluza de Salud Pública (EASP), Granada, Spain; Instituto de Investigación Biosanitaria ibs.GRANADA, Granada, Spain; Centro de Investigación Biomédica en Red de Epidemiología y Salud Pública (CIBERESP), Instituto de Salud Carlos III, C. de Melchor Fernández Almagro, Madrid, Spain; Ministry of Health of the Basque Government, Sub Directorate for Public Health and Addictions of Gipuzkoa, San Sebastian, Spain; Biodonostia Health Research Institute, Epidemiology of Chronic and Communicable Diseases Group, San Sebastián, Spain; Department of Epidemiology, Murcia Regional Health Council, IMIB-Arrixaca, Murcia, Spain; Navarra Institute for Health Research (IdiSNA), Pamplona, SpainInstituto de Salud Pública de Navarra, IdiSNA, Navarre Institute for Health Research, Pamplona, Spain; Department of Clinical Science in Malmö, Lund University, Clinical Research Center, Malmö, Sweden; Department of Emergency and Internal Medicine, Skåne University Hospital, Malmö, Sweden; Department of Public Health and Clinical medicine, Umeå University, Umeå, Sweden; Department of Epidemiology and Global Health, Umeå University, Umeå, Sweden; Department of Epidemiology and Biostatistics, School of Public Health, Imperial College London, London, United Kingdom; British Heart Foundation Cardiovascular Epidemiology Unit, Department of Public Health and Primary Care, University of Cambridge, Cambridge, United Kingdom; National Institute for Health Research Blood and Transplant Research Unit in Donor Health and Genomics, University of Cambridge, Cambridge, United Kingdom; British Heart Foundation Centre of Research Excellence, University of Cambridge, Hills Road, Cambridge, United Kingdom; Health Data Research UK Cambridge, Wellcome Genome Campus and University of Cambridge, United Kingdom; Department of Human Genetics, Wellcome Sanger Institute, Hinxton, United Kingdom

**Keywords:** amino acids, dietary protein, ischaemic stroke, haemorrhagic stroke, nutritional epidemiology, prospective cohort

## Abstract

**Background:** Previously reported associations of protein-rich foods with stroke subtypes have prompted interest in assessment of individual amino acids. We examined the associations of dietary amino acids with risks of ischaemic and haemorrhagic stroke in the EPIC study.

**Methods:** We analysed data on 356,142 participants from seven European countries. Dietary intakes of 19 individual amino acids were assessed using validated country-specific dietary questionnaires, calibrated using additional 24-hour dietary recalls. Multivariable-adjusted Cox regression models were used to estimate hazard ratios (HRs) and 95% confidence intervals (CIs) of ischaemic and haemorrhagic stroke in relation to intake of each amino acid. The role of blood pressure as a potential mechanism was assessed in 267,642 (75%) participants.

**Results:** After a median follow-up of 12.9 years, 4,295 participants had an ischaemic stroke and 1,375 participants had a haemorrhagic stroke. After correction for multiple testing, higher intake of proline (as percent of total protein) was associated with 12% lower risk of ischaemic stroke (HR per 1 SD higher intake 0.88; 95% CI 0.82, 0.94). The association persisted after mutual adjustment for all other amino acids, systolic and diastolic blood pressure. The inverse associations of isoleucine, leucine, valine, phenylalanine, threonine, tryptophan, glutamic acid, serine and tyrosine with ischaemic stroke were each attenuated with adjustment for proline intake. For haemorrhagic stroke, no statistically significant associations were observed in the continuous analyses after correcting for multiple testing.

**Conclusion:** Higher proline intake was associated with a lower risk of ischaemic stroke, independent of other dietary amino acids and blood pressure.

## Background

While stroke is the second leading cause of death worldwide, the incidence of stroke varies substantially between countries reflecting differences in risk factors including variations in dietary intakes [1]. Differences in dietary protein has been suggested to be important, but previous studies of the associations of dietary protein intake with risk of stroke have reported conflicting results, possibly reflecting heterogeneity by pathological stroke types and dietary sources of protein [2, 3]. The different sources of dietary protein reflect a different profile of individual dietary amino acids. For example, meat and meat products are a main source of the essential amino acids and glycine, while grain products are a main source of dietary cysteine [4]. A previous prospective study reported an inverse association of dietary cysteine with risk of total stroke [5], and another reported an inverse association of glutamic acid but positive association of glycine with stroke mortality [6], but overall there is very limited evidence on the topic. Moreover, it is unclear whether the absolute quantity or the proportions of each individual dietary amino acid (i.e. protein quality) that make up total dietary protein would be more relevant for health. The two main stroke subtypes (ischaemic and haemorrhagic) also have different dietary [7] and non-dietary (including genetic) risk factors [8, 9], which highlights the importance of examining stroke subtypes separately. For example, previous analyses in EPIC (the European Prospective Investigation into Cancer and Nutrition) reported inverse associations of ischaemic stroke with consumption of fruit and vegetables, dietary fibre and dairy products, while the risk of haemorrhagic stroke was positively associated with egg consumption [7]. Therefore, any associations with dietary amino acids are likely to vary by stroke type.

The aim of the present study was to assess the associations of both absolute and relative quantities of individual dietary amino acids with risk of stroke subtypes in a prospective study of 356,000 participants from seven European countries. Additionally, we aimed to assess if these associations were independent of blood pressure, the major risk factor for both subtypes of stroke.

## Methods

### Study population

The present analyses involved 356,142 participants recruited from 20 centres between 1992 and 2000 in seven European countries (Denmark, Germany, Italy, the Netherlands, Spain, Sweden, and the UK), who participated in the EPIC study. Details of the study design have been described previously [10, 11]. The inclusion and exclusion criteria for the present study are shown in Supplementary figure 1. All participants provided written informed consent, and the study protocol was approved by the ethical review board of IARC and the institutions where the participants were recruited [10].

### Data collection

At recruitment all participants completed questionnaires on medical history and socio-demographic factors, as well as validated country-specific dietary questionnaires (mostly food frequency questionnaires or diet histories) which asked about diet in the previous year [10]. In addition, a stratified random sample of 8% of participants across all centres also completed a standardised and computerised 24-hour recall, on average ∼1.4 years after recruitment, which was used to calibrate the dietary exposures to reduce between-centre heterogeneity and to correct for measurement error [12, 13]. The calibration method and rationale have been described previously [7, 12, 13].

Based on reported dietary intakes in both the baseline dietary questionnaires and 24-hour recalls, estimates of individual dietary amino acids were derived by matching to the National Nutrient Database for Standard Reference of the United States (developed at the United States Department of Agriculture, or USDA) food composition tables, and appended to the EPIC Nutrient Database. The matching and validation process has been reported in detail [14]. USDA estimates of dietary amino acids were used because the equivalent estimates were not available from all ten national Food Composition Databases that were originally used in EPIC. Altogether, both calibrated and observed estimates were available for the intakes of 19 individual dietary amino acids. Of the 20 standard amino acids, glutamine and asparagine were not available as these two amino acids were affected by deamination reactions during the acid hydrolysis process used to compile the USDA database [15, 16]. Cystine (two cysteine molecules joined by a disulfide bond) [17] was estimated instead of cysteine, while hydroxyproline, which is not a standard amino acid, was also estimated. The sums of branched-chain amino acids (isoleucine, leucine, valine), other essential amino acids (histidine, lysine, methionine, phenylalanine, threonine and tryptophan) and non-essential amino acids (alanine, arginine, aspartic acid, cystine, glutamic acid, glycine, hydroxyproline, proline, serine and tyrosine) were also calculated.

The primary outcomes were ischaemic stroke (ICD 9 433-434 or ICD 10 I63) and haemorrhagic stroke (ICD 9 430-431 or ICD 10 I60-I61). In addition, we also considered intracerebral haemorrhage (ICD 9 431 or ICD 10 I61) and subarachnoid haemorrhage (ICD 9 430 or ICD 10 I60) as separate outcomes. Both non-fatal and fatal incident events were considered, and details of the ascertainment process and validation methods have been previously reported [7, 11]. Details on the data collection for blood pressure (available for most of the cohort) and other covariates have also been reported previously [7, 11].

### Statistical analyses

Cohort characteristics and dietary intakes of amino acids at baseline were summarised as means (SD), median (25^th^, 75^th^ percentile) or absolute numbers (%). Spearman correlation coefficients were estimated between the individual dietary amino acids, and also for dietary amino acids with dietary protein and food sources of protein. Intakes of dietary amino acids were expressed in three different ways, primarily as individual amino acids modelled as a percentage of total protein intake (% of total protein), and secondarily as individual amino acids modelled as grams per day (g/day) and grams per 1000 kcal (g/1000 kcal). The three approaches tested the hypotheses that the relative protein quality or protein makeup, the absolute quantity of amino acids, and the relative quantity of amino acids to overall diet were relevant to risk of stroke. The primary approach of expressing amino acids as a percentage of total protein was selected since this approach was less susceptible to collinearity with other dietary variables [18]. Because the sum of the individual amino acids did not add up to 100% of total protein (mean=86.2%, partly because glutamine and asparagine were missing), we included post hoc analyses expressing the amino acids as a percentage of the sum of all available amino acids. The associations of total dietary protein with stroke risk were also assessed, by expressing protein in g/day and g/1000 kcal.

Using multivariable-adjusted Cox regression, we assessed the hazard ratios (HRs) and 95% confidence intervals (CIs) for sex-specific SD differences in calibrated intakes of each dietary amino acid, and sex-specific SD differences and sex-specific fifths of observed intakes. For each association, a test of trend was estimated by fitting the calibrated or observed intake of each amino acid as a continuous variable when modelling per SD differences, or by fitting the median values of each fifth as a pseudo-continuous variable when modelling differences by fifths of intake. Tests of non-linearity were performed by using a likelihood-ratio test to compare models with the exposure fitted as a continuous variable versus a categorical variable (fifths of intake), based on the observed intake.

The underlying time variable for the Cox regression was age at recruitment to age at diagnosis, death or administrative censoring. All analyses were stratified by sex and EPIC centre, and adjusted for age at recruitment (continuous, Model 1). Model 2 additionally adjusted for total energy intake (continuous, calibrated or observed intake based on USDA estimates). Model 3 additionally adjusted for smoking status and intensity (never, former, current<10, 10-19, 20+ cigarettes/day, unknown), current calibrated or observed alcohol consumption (non-drinker (<0.1), 0.1-4.9, 5.0-14.9, 15-29.9, 30-59.9, 60+ g/day based on USDA estimates), physical activity (inactive, moderately inactive, moderately active, active, unknown), employment status (employed or student, neither employed nor student, unknown) and level of education completed (none or primary, secondary, vocational or university, unknown). Model 4 additionally adjusted for self-reported history of diabetes (yes, no, unknown), prior hypertension (yes, no, unknown) and prior hyperlipidaemia (yes, no, unknown). Model 5 additionally adjusted for body mass index (<22.5, 22.5-24.9, 25.0-27.4, 27.5-29.9, ≥30.0 kg/m2, unknown), and was considered the main model in these analyses. As total energy intake was held constant (i.e. adjusted for) in the regression model, a higher intake of any individual amino acid, which contributes to total energy, would result in a simultaneously lower intake of some other energy-providing foods or nutrients [19]. Therefore, in model 6, we removed the adjustment for total energy intake thus allowing the comparison of the results with (Model 5) and without (Model 6) a theoretical substitution effect of the amino acid of interest for an unspecified energy-providing source. To further assess the influence of major macronutrients and to allow the interpretation of the findings when holding total protein intake constant, we fitted an additional model based on Model 6 (without energy intake), but with the inclusion of intakes of total protein (continuous), total carbohydrates (continuous), saturated fats (continuous) and unsaturated fats (continuous). Likelihood-ratio χ^2^ statistics were estimated at each adjustment level (for models 1-6) by comparing regression models with and without inclusion of the amino acids of interest using a likelihood ratio test [20]. The proportional hazards assumption was assessed using Schoenfeld residuals. The assumption was not met for two covariates in the models for ischaemic stroke (history of diabetes and prior hypertension) and for haemorrhagic stroke (prior hypertension and BMI). However, fitting these covariates as stratifying variables fulfilled the assumption and yielded close to identical estimates as adjusting, so they were retained as adjustment variables.

To investigate whether the observed associations for the amino acids were independent from the other amino acids, we included additional models mutually adjusting for each of the other dietary amino acids, one at a time. We also included post-hoc analyses adjusting the amino acids that remain significant across different models for its major food sources. To assess whether the associations were independent of blood pressure, we estimated the adjusted mean systolic and diastolic blood pressure by fifths of each dietary amino acid in the subset of participants with blood pressure measurements (n=267,642), using the same covariate adjustments as above. Additionally, we repeated the Cox regression analyses in this subset (based on Model 5), with and without further adjustment for systolic or diastolic blood pressure.

As secondary analyses, we repeated the main analyses for the two subtypes of haemorrhagic stroke (intracerebral haemorrhage and subarachnoid haemorrhage). To assess potential heterogeneity of the associations by subgroups of the major confounders and to assess residual confounding, we also examined the results stratified by age at recruitment (<55, 55-64, ≥65 years), sex, BMI (<25, 25-29.9, ≥30 kg/m^2^), smoking status (never, former, current smokers), alcohol drinking (non-drinkers, moderate drinkers <15g/day, heavy drinkers ≥15g/day) and history of diseases (as a dichotomy of no disease history vs history of diabetes, hypertension or hyperlipidaemia). We also examined the associations based on observed intakes stratified by EPIC country and pooled using a meta-analysis approach. Tests of heterogeneity of trend between subgroups were conducted by comparing the risk coefficients in each subgroup using inverse variance weighting, testing for statistical significance with a χ^2^ test on *k*-1 degrees of freedom, where *k* is the number of subgroups. To assess whether the overall results might be influenced by reverse causation, we repeated the analyses for ischaemic and haemorrhagic stroke after excluding the first four years of follow-up. A complete case analysis was also performed to assess the potential influence of missing covariates, which were coded as a missing category in the main analyses.

All tests for statistical significance were two sided. Conventional p-values are reported, but results were interpreted with consideration of multiple testing. To account for multiple testing while allowing for correlation between the exposures, we conducted a principal component analysis of the exposure variables, and determined that the first four principal components explained 99% of the total variation in the exposure data [21, 22]. Consequently, the effective number of independent tests was determined to be four, and the statistical significance level after Bonferroni correction for multiple testing based on this number was defined as 0.05/4=0.0125. All analyses were performed using Stata version 17.0 (StataCorp, TX, USA). All figures were generated using “Jasper makes plots” package [23] version 2-266 in R version 4.2.1.

## Results

### Baseline and dietary characteristics

The baseline characteristics of the study participants are shown in Table 1 and additionally in detail in Supplementary table 1. Intakes of dietary amino acids, total protein and total energy are shown by sex in Supplementary table 2, and by EPIC country in Supplementary table 3. The Spearman correlation coefficients between the individual dietary amino acids are shown in Supplementary figure 2, and for amino acids with dietary protein and food sources of protein in Supplementary figures 3 and 4 respectively.

**Table 1:**
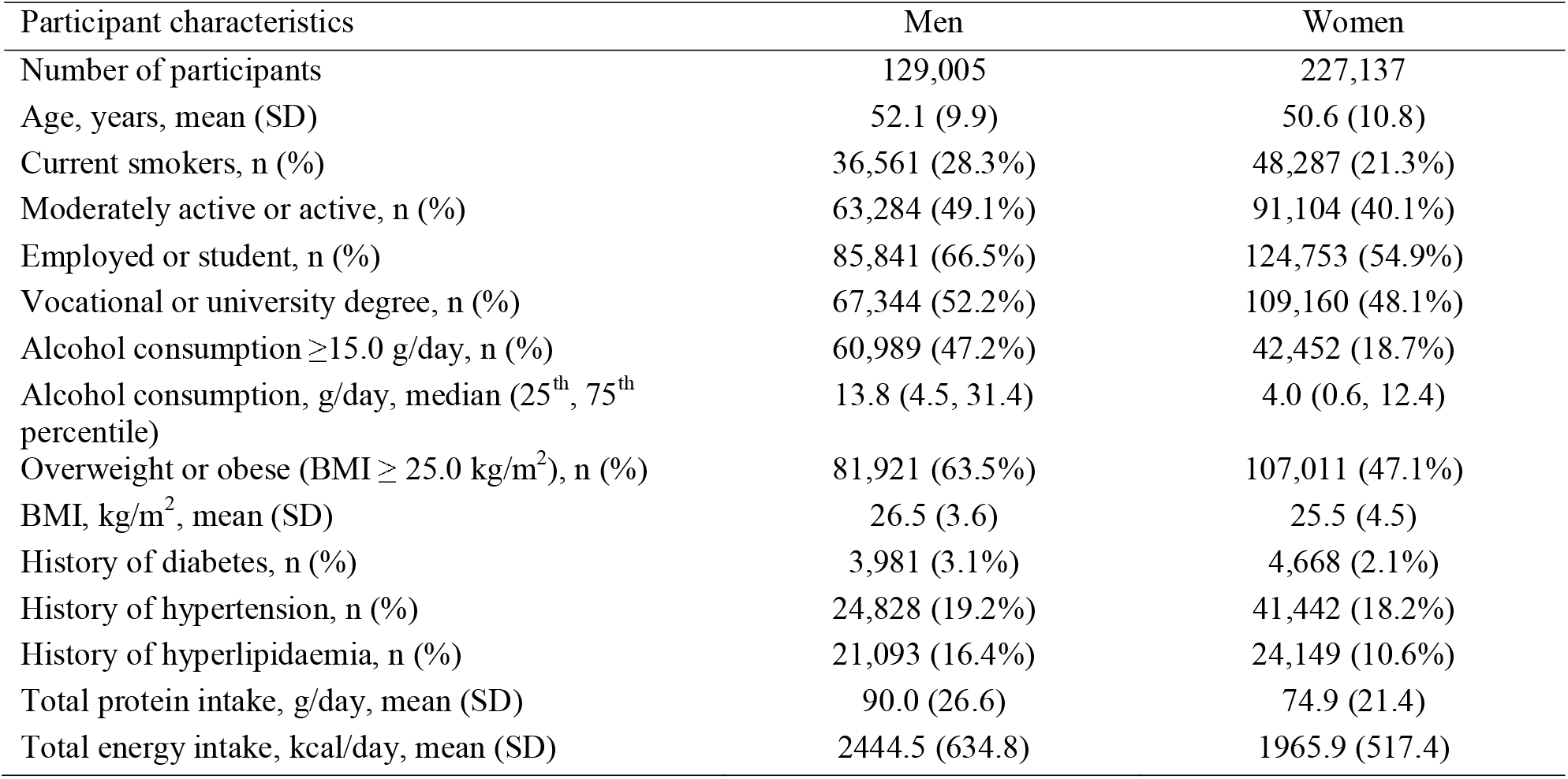
Participant characteristics at recruitment by sex in the EPIC study.

### Associations of dietary amino acids with stroke risk

After a median follow-up of 12.9 years, there were 4,295 cases of ischaemic stroke and 1,375 cases of haemorrhagic stroke among 356,142 participants (4,529,626 person-years in total). In the fully adjusted models, associations between dietary amino acids and risks of ischaemic and haemorrhagic stroke were numerically similar irrespective of whether the amino acids were expressed as a percentage of total protein (Figure 1, Supplementary tables 4-5), in g/day (Supplementary figure 5, Supplementary tables 4-5), g/1000 kcal (Supplementary figure 6, Supplementary tables 4-5), or as a percentage of total amino acids (Supplementary figure 7, Supplementary tables 4-5). Unless otherwise stated, the primary results reported on the associations that remained statistically significant after correcting for multiple testing, or results from categorical analyses where we observed non-linear associations.

**Figure 1.**
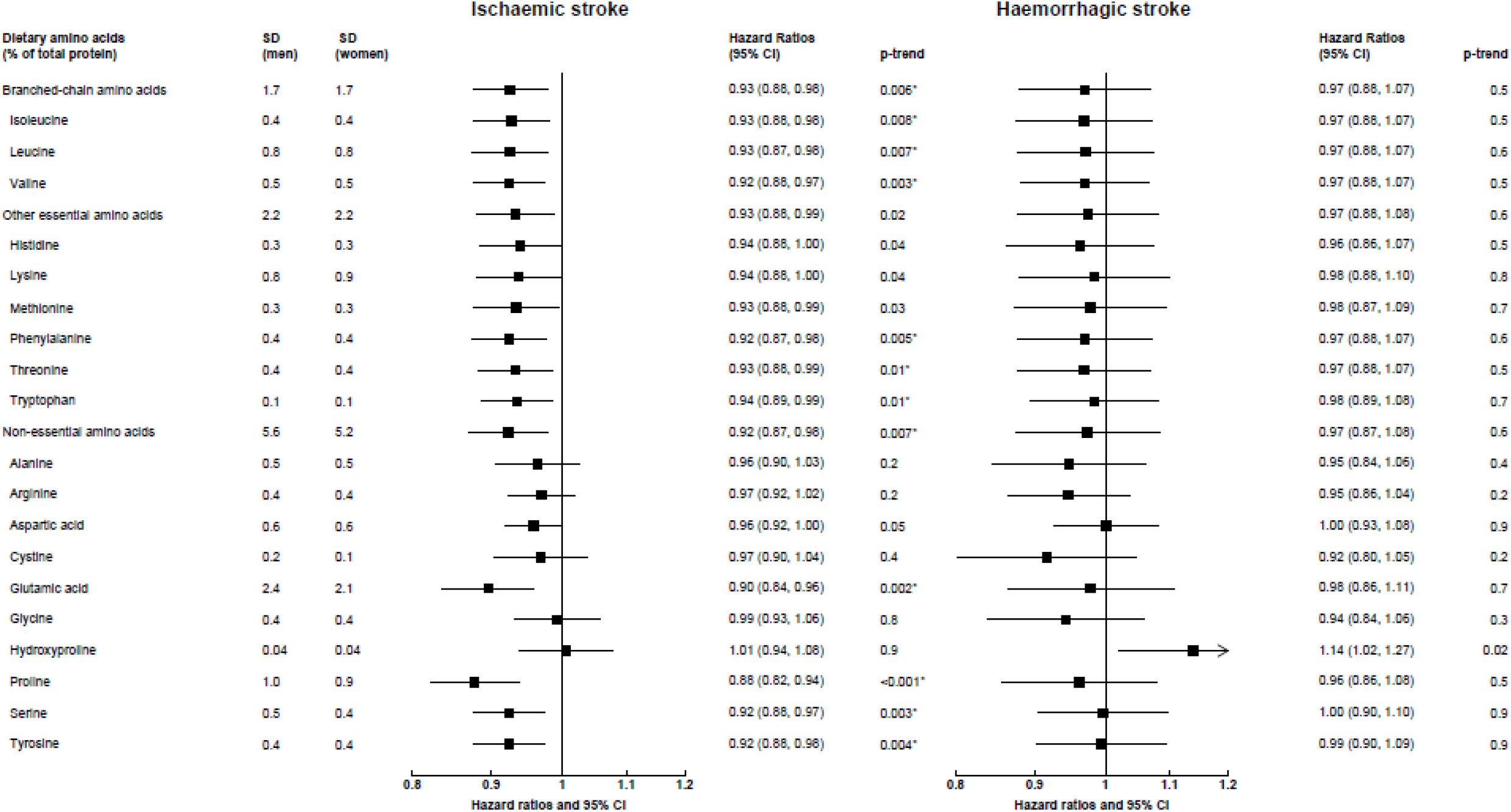
Hazard ratios (95% confidence intervals) for ischaemic (4295 cases) and haemorrhagic stroke (1375 cases) by increments of calibrated intakes of dietary amino acids (as percent of total protein) Hazard ratios modelled per 1 sex-specific SD increment in dietary amino acids, expressed as percent of total protein. The model was stratified by sex and centre, and adjusted for age, calibrated energy intake, smoking, calibrated alcohol consumption, physical activity, employment status, highest level of education completed, history of diabetes, prior hypertension, prio hyperlipidaemia, body mass index. Estimates with an asterisk denote ones which were statistically significant after correcting for multiple testing.

When expressing the amino acids as a percentage of total protein, in the fully adjusted models (Model 5) we found inverse associations of ischaemic stroke for each 1 SD higher calibrated intake of total and individual branched-chain amino acids (HR [95% CI] per SD higher of total: 0.93 [0.88, 0.98], isoleucine: 0.93 [0.88, 0.98], leucine: 0.93 [0.87, 0.98], valine: 0.92 [0.88, 0.97]), phenylalanine (0.92 [0.87, 0.98]), threonine (0.93 [0.88, 0.99]), tryptophan (0.94 [0.89, 0.99]), total non-essential amino acids (0.92 [0.87, 0.98]), glutamic acid (0.90 [0.84, 0.96]), proline (0.88 [0.82, 0.94]), serine (0.92 [0.88, 0.97]) and tyrosine (0.92 [0.88, 0.98]) (Figure 1). Glycine showed a non-linear association with ischaemic stroke in the categorical analysis (HR [95% CI] 1.28 [1.14, 1.43] for top versus bottom fifth, p non-linearity<0.001), but only when expressing glycine as a percentage of estimated intakes of total protein or total amino acids (Supplementary table 4). For haemorrhagic stroke, no statistically significant associations were observed in the continuous analyses after correcting for multiple testing (Figure 1). In categorical analyses, lower risks were observed from the second to top fifth of proline intake compared to bottom fifth (top versus bottom fifth: 0.72 [0.58, 0.90], p-trend across categories=0.006, Supplementary table 5). No statistically significant associations were observed for total protein with either ischaemic or haemorrhagic stroke (Supplementary figures 4-5, Supplementary tables 4-5), or in any analyses examining the two haemorrhagic stroke subtypes as outcomes (Supplementary table 6). Results from the different covariate adjustment models are shown in Supplementary tables 7-9.

### Mutual adjustment of dietary amino acids

Results with mutual adjustment for each of the other amino acids for the ten amino acids that were inversely associated with ischaemic stroke risk, as reported above, are shown in Figure 2 (for the five amino acids that were significantly associated in both the calibrated and observed analyses) and in Supplementary figure 8 (for the five amino acids that were significantly associated in the calibrated analyses only). The inverse associations for proline persisted after adjustment for all other amino acids, but the associations for other amino acids were attenuated upon adjustment for proline. However, the higher risk of ischaemic stroke comparing top to bottom fifth of glycine intake remained with adjustment for proline (1.27 [1.12, 1.43], results not shown in tables). In post-hoc analyses, the inverse associations for proline with ischaemic stroke remained upon mutual adjustment for its different food sources (e.g. 0.89; 0.83, 0.96 after mutual adjustment for cheese, with no significant association for cheese in the same model, results not shown in tables). Based on these findings, the changes in the HRs and likelihood ratio χ^2^ statistics for proline are also depicted graphically in Figure 3, by each level of covariate adjustment.

**Figure 2.**
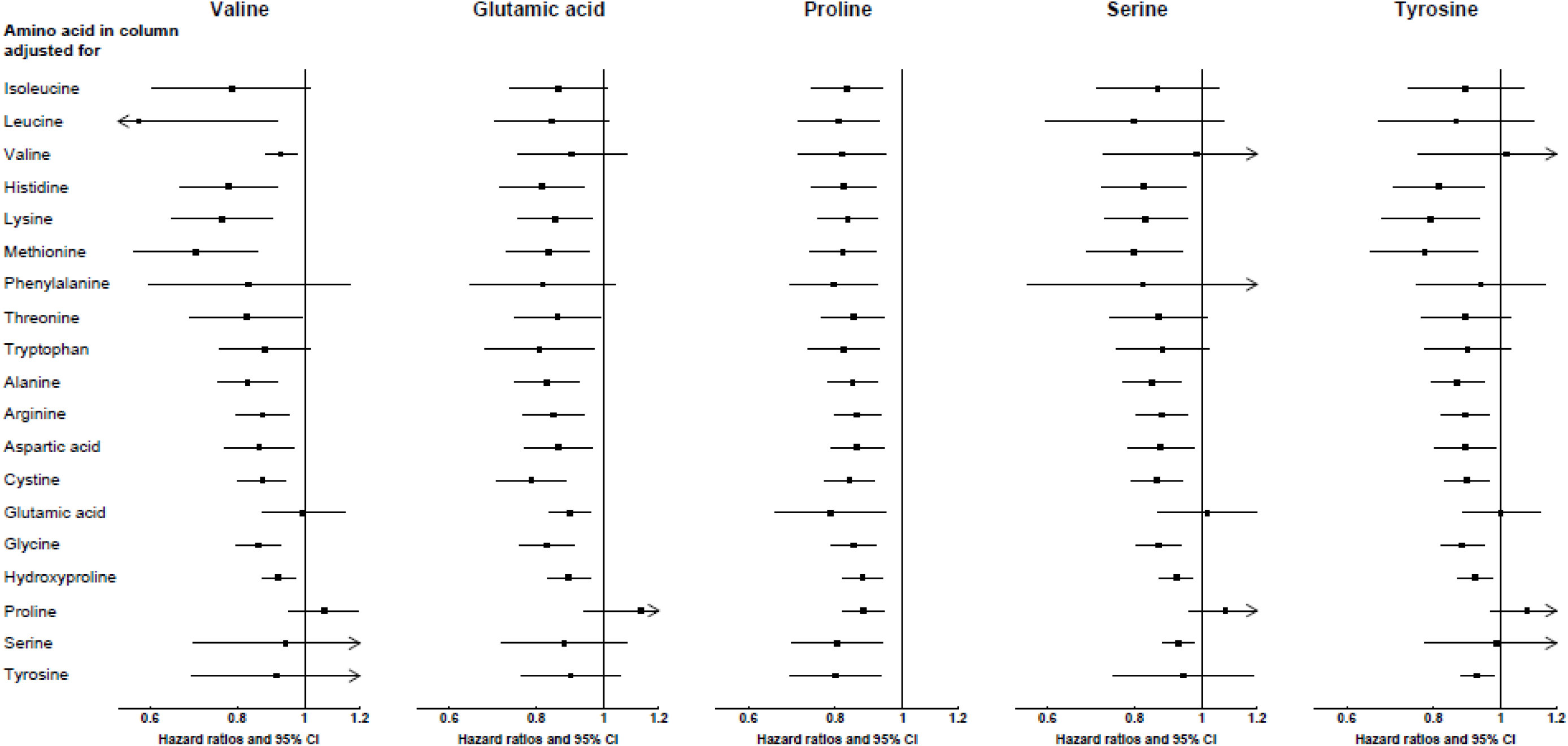
Hazard ratios (95% confidence intervals) for the association between selected dietary amino acids and ischaemic stroke (4295 cases), with mutual adjustment for each of the other amino acids. Results are shown for amino acids which were statistically significantly associated with ischemic stroke in both calibrated and uncalibrated analyses. Hazard ratios modelled per 1 sex-spe SD (as shown in Figure 1) increment in dietary amino acids in column heading, expressed as percent of total protein, with multivariable adjustment plus amino acid on the left panel. If the amino acid in column and in left panel are the same, the result is interpretable as the hazard ratio for the amino acid based on the multivariable adjusted model.

**Figure 3.**
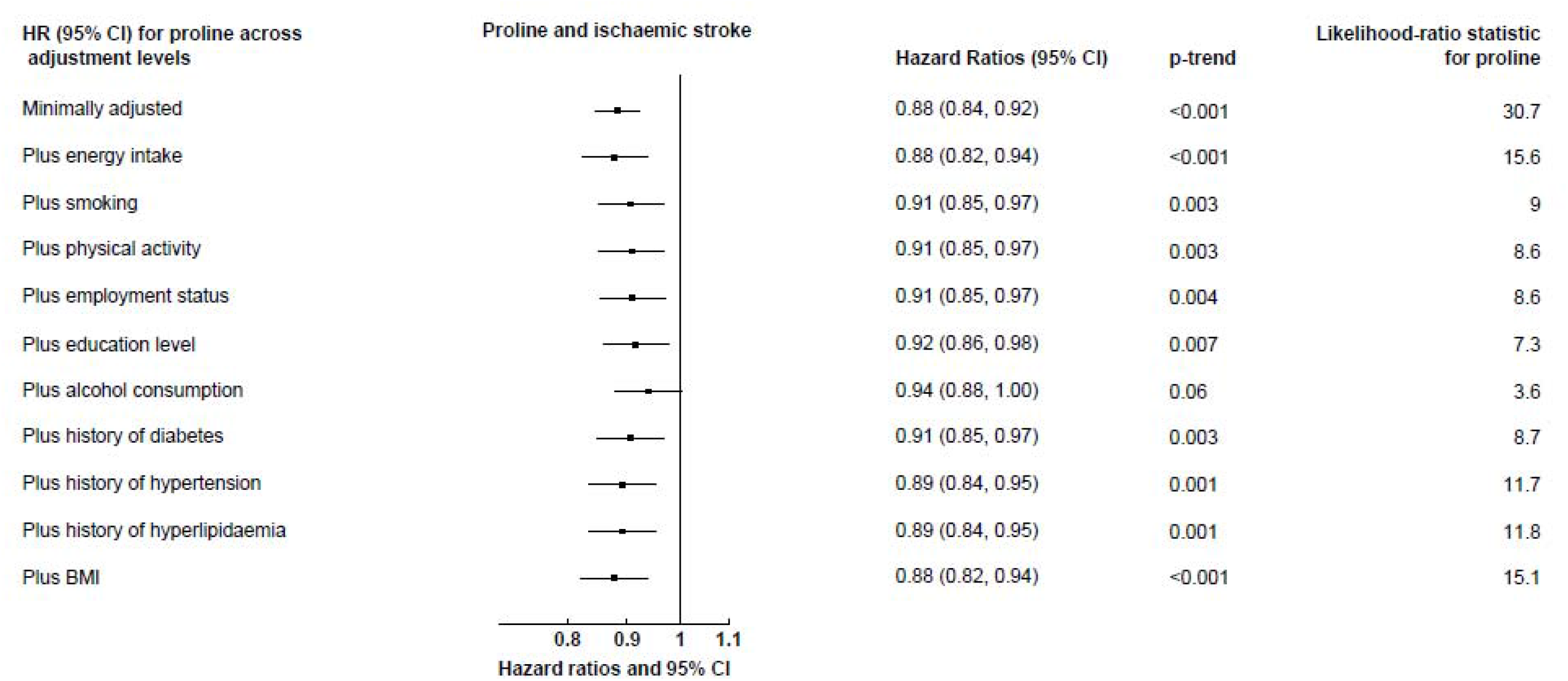
Hazard ratios (95% confidence intervals) for the association between dietary proline and ischaemic stroke (4295 cases), with additional adjustmen each covariate. Hazard ratios modelled per 1 SD increment (1 SD=1.0 in men, 0.9 in women) in calibrated proline intake, expressed as percent of total protein. The minimally adjusted model was stratifie sex and centre, and adjusted for age (continuous). The likelihood-ratio test statistic for proline is the _χ_2 statistic based on a likelihood ratio test comparing models with and without proline the varying levels of covariate adjustment. The changes in the _χ_2 statistic across models could be interpreted as a measure of the extent to which the covariates could account for any associations between proline and ischaemic stroke risk.

### Assessment of role of blood pressure

The adjusted mean levels of systolic and diastolic blood pressure across fifths of dietary amino acids are shown in Supplementary tables 10-11. Across all amino acids, higher intakes were associated with higher blood pressure, though the differences were very small (up to 1 mmHg difference in most cases). As expected from these small differences, the results for both ischaemic and haemorrhagic stroke were almost unchanged after further adjustment for either systolic or diastolic blood pressure (Supplementary table 12).

### Sensitivity analyses

In stratified analyses, the main findings were consistent between subgroups of age, sex, BMI, smoking status, alcohol drinking and disease status for both ischaemic and haemorrhagic stroke (Supplementary tables 13-24). The pooled estimates by country using a meta-analysis approach were similar (Supplementary tables 25-26), and results were consistent after excluding the first four years of follow-up (Supplementary table 27) or based on a complete case analysis (Supplementary table 28).

## Discussion

In this large prospective study, we observed inverse associations of dietary intakes of proline with risk of ischaemic stroke after adjustment of potential confounders, independent of the other amino acids and of total protein intake. The associations were consistent regardless of how the amino acids were modelled (i.e. percentage of protein, g/day and g/1000 kcal), and across all subgroup analyses. Although inverse associations with ischaemic stroke were initially observed for some other amino acids, these might be explained by the high correlations between these amino acids and proline, since the associations were attenuated after adjustment for proline. We further observed evidence of a higher risk of ischaemic stroke in individuals with the highest intakes of dietary glycine, and a lower risk of haemorrhagic stroke in people with higher intakes of dietary proline, though these associations differed depending on the way dietary amino acids were modelled in the analyses.

The present analyses provide important novel evidence on the associations between dietary amino acids and stroke risk, as only two prospective studies have previously published on the topic [5, 6]. The Swedish Mammography Cohort, with 1751 cases of total stroke (1311 ischaemic and 264 haemorrhagic after 10 years), reported an inverse association between dietary cysteine and the risk of total stroke, but no other associations including for proline were observed after mutual adjustment of the amino acids [5]. In contrast to that study, we did not find an association of cystine with either stroke subtype; however, although cystine is expected to be highly correlated with cysteine, their molecular structures are different, so the two sets of results are not directly comparable. The Takayama Study in Japan, with 677 deaths from stroke (393 ischaemic and 153 intracerebral haemorrhage after 16 years), showed that glutamic acid intake was inversely associated with total stroke mortality among women, while glycine intake was positively associated with ischaemic stroke mortality in men with no prior history of hypertension; the study did not report on proline [6]. In our study, we also observed an inverse association of glutamic acid intake with ischaemic stroke, but this association was attenuated when we further adjusted for dietary proline. In accordance with the Takayama Study, we also found a higher risk of ischaemic stroke for people in the highest fifth of glycine intake. The differences in intakes of the amino acids across populations should be considered when comparing the findings. For example, the average intakes of both glutamic acid and glycine when expressed as a percentage of total protein in the EPIC study is closer to the lower end of the intakes reported in the Takayama study. Overall, compared to the two previous studies, the current study is over three- and ten-fold larger in case numbers. We also focused on examining ischaemic and haemorrhagic stroke separately, based on prior evidence of heterogeneity in risk factors by stroke subtypes. Overall, the associations of dietary amino acids with haemorrhagic stroke were less clear. As the number of haemorrhagic stroke cases in the study was comparatively low, further investigations in other populations with higher numbers of haemorrhagic stroke are warranted.

Previous studies have largely attributed the observed associations of dietary amino acids with stroke risk to the influences of the amino acids on blood pressure. The INTERMAP study reported inverse associations of glutamic acid and to a lesser degree proline [18], but positive associations of glycine intake [24], with blood pressure. Our results did not suggest an important role of blood pressure in explaining any of the findings, given that we did not observe any meaningful differences in blood pressure by amino acid intake, nor did we observe any differences in the HRs after further adjusting for blood pressure. However, blood pressure and diet in EPIC were both measured at recruitment, which does not allow the evaluation of a temporal association. Additionally, the use of blood pressure measurements assessed at a single time point as an indicator of long-term blood pressure is also prone to random measurement error, which could result in an underestimation of its role in explaining any associations between amino acid intakes and stroke risk. Furthermore, blood pressure measurements in EPIC were not taken using a standardised protocol across centres [25], but our analyses were stratified by centre, so any influence on the results should be small.

The possible biological mechanisms for the association between proline and stroke risk are not yet well defined. Despite being non-essential amino acids, proline, glycine and hydroxyproline are the major constituents of collagen, which accounts for about one third of proteins in the human body [26]. Collagen has an important role in maintaining the structure and strength of connective tissues, cartilage and blood vessels [26, 27], and studies from animal models have suggested that endogenous synthesis of these amino acids may not be adequate for growth or collagen production [26, 28]. Because proline is the only precursor for hydroxyproline, which in turn is a precursor for glycine [26, 29], dietary intakes of proline may be of particular importance for collagen synthesis.

In contrast to other amino acids, proline contains an imino group in its structure instead of a primary amino group, which results in its distinctive cyclic structure and exclusion from standard amino acid metabolism [30, 31]. For this reason, the family of proline-containing peptides, collectively called glycoprolines, cannot be degraded by most peptidases owing to the presence of proline in their structures, and thus are more likely to enter the bloodstream and exert various biological effects, including possible atheroprotective or neuroprotective roles, regulation of insulin-like growth factor-I (IGF-I) homeostasis and lipid metabolism including lowering of total cholesterol [32]. Glycoprolines may be derived from collagen or IGF-Ia study in rats has shown that oral administration of cyclic glycine-proline, a glycoproline derived from IGF-I, promoted neural plasticity and remodelling in animals with focal ischaemic lesions, suggesting a neuroprotective effect of cyclic glycine-proline against ischaemic stroke or its recovery [33]. In human studies, both cyclic glycine-proline concentrations [34] and proline concentrations [35–37] have been proposed as prognostic markers of stroke recovery or stroke events. However, the directions of the changes observed have been inconsistent. Other studies have also reported low correlations between dietary intake and circulating levels of proline [38]. Therefore, further research into the link between proline intake and the physiological effects of circulating proline, as well as the role of proline in both stroke incidence and prognosis (recovery or mortality) is needed.

Among dietary sources, the results of the present study showed that dairy products, especially cheese, was most highly correlated with proline intake. Therefore, the findings of lower risks of ischaemic stroke in people with higher proline intake in the present study are consistent with previous observations of lower ischaemic stroke risk in people with higher intake of cheese and yogurt in the same cohort [7]. Despite being a major source of saturated fat, higher intakes of dairy products have been inversely associated with cardiovascular disease outcomes in a meta-analysis of prospective studies [39], suggesting that some components in cheese or dairy products might be protective. For example, studies had suggested that the specific saturated fatty acid isomer composition of different foods [40], or the calcium [41] or probiotics content [42] of dairy foods may account for their associations with stroke risk. As the observed association for proline remained upon adjustment for cheese and other food sources, the findings of the present study suggest that the amino acid profiles of dairy products and their associations with CVD warrant further research. Meanwhile, dietary intake of glycine was strongly correlated with red meat intake, which was previously found to be modestly but positively associated with ischaemic stroke risk in EPIC, though this association was attenuated after adjustment for fibre intake [7]. Further population-based and mechanistic studies are needed to replicate the observed associations between dietary amino acids and risks of stroke subtypes, prior to investigating any implications for potential dietary interventions.

The chief strengths of this study include the large sample size with participants recruited from seven European countries, the prospective design and prolonged duration of follow-up. EPIC is also a well-characterised cohort with extensive dietary and lifestyle/health behavioural data, allowing us to examine estimated intakes of 19 dietary amino acids with stroke risk, while adjusting for multiple confounders. The ascertainment of stroke cases in this cohort was either completely or partially validated depending on EPIC country, and previous dietary analyses have found no heterogeneity in the associations by the extent of stroke validation [7]. To adjust for potential bias due to differences between the country-specific dietary questionnaires, we also applied calibration to the dietary data using additional standardised 24-hour recalls to reduce between-centre heterogeneity and to reduce measurement error. The study also had several limitations. Diet was ascertained using a single dietary questionnaire collected at baseline, which meant that we were not able to account for dietary changes during follow-up. We were unable to assess the associations of dietary glutamine, asparagine or cysteine with stroke risk, as estimates of these amino acids were not available from the USDA database. The matching of amino acids data based on a single US database in different countries across Europe may also be a limitation, and the absolute validity of the amino acid estimates assessed using FFQs and 24-hour recalls is not known, although good agreement and thus relative validity has been shown for estimates of total energy and protein intakes between the USDA and country-specific databases in the previous validation study [14]. Because of the large number of tests conducted, false positive findings due to chance are possible, but we focused on the findings that survived correction for multiple testing. We also conducted analyses further adjusting for each one of the other amino acids, which given their high correlations might constitute over-adjustment, but this approach also helped highlight the clear associations with proline. As with any observational study, the present study could not establish causality, and cannot fully exclude residual confounding or reverse causation. However, we assessed the former by comparing the changes in χ^2^ statistics across models, and limited the latter possibility by our sensitivity analysis excluding the first four years of follow-up. Information on other potential mediators such as cholesterol concentrations was only available in a small subset of participants at study baseline, so we were unable to access their potential effects. Lastly, the present study was largely restricted to white European individuals, therefore the findings may not be generalisable to other populations which have different sources of protein intake and dietary amino acid profiles, as well as genetic differences.

## Conclusions

This large study demonstrated an inverse association of both absolute intake and relative proportions of dietary proline with risk of ischaemic stroke, independent of the other amino acids. The suggestive associations of lower risk of haemorrhagic stroke in people with higher proline intake, and higher risk of ischaemic stroke in people with higher proportions of glycine intake, also warrant further investigation. These observations were largely unexplained by differences in blood pressure. Further research is needed from both large-scale population-based studies and mechanistic studies to replicate these findings, in addition to genetic studies and intervention trials to assess the causal relevance of these associations and possible implications for prevention.

## Data Availability

For information on how to submit an application for gaining access to EPIC data and/or biospecimens, please follow the instructions at http://epic.iarc.fr/access/index.php

## Acknowledgments

The authors thank all participants in the EPIC cohort for their invaluable contribution to the study.

## Authors’ contributions

The study was conceived and designed by TYNT and TJK with input from all other authors. TYNT analysed the data and wrote the first draft of the manuscript, with input from TJK, RC, JAS, IH, UN, NGF, FI, RCT, EW, KA, CCD and YTVS. All other authors provided the data and revised the manuscript critically for important intellectual content. TYNT had primary responsibility for final content. All authors read and approved the final manuscript.

## Disclaimer

Where authors are identified as personnel of the International Agency for Research on Cancer / World Health Organization, the authors alone are responsible for the views expressed in this article and they do not necessarily represent the decisions, policy or views of the International Agency for Research on Cancer / World Health Organization.

## Conflicts of interest

The authors have no conflicts of interest to declare.

## Funding

This work was supported by a Nuffield Department of Population Health Fellowship (TYNT), the UK Medical Research Council (MR/M012190/1), Cancer Research UK (C8221/A19170 and 570/A16491), and the Wellcome Trust (Our Planet Our Health, Livestock Environment and People 205212/Z/16/Z). EPIC-CVD has been supported by the European Union Framework 7 (HEALTH-F2-2012-279233), the European Research Council (268834), the UK Medical Research Council (G0800270 and MR/L003120/1), the British Heart Foundation (SP/09/002 and RG/08/014 and RG13/13/30194), and the UK National Institute of Health Research. The establishment of the study subcohort was supported by the EU Sixth Framework Programme (FP6) (grant LSHM_CT_2006_037197 to the InterAct project) and the Medical Research Council Epidemiology Unit (grants MC_UU_12015/1 and MC_UU_12015/5). The coordination of the European Prospective Investigation into Cancer and Nutrition (EPIC) is financially supported by International Agency for Research on Cancer (IARC) and also by the Department of Epidemiology and Biostatistics, School of Public Health, Imperial College London which has additional infrastructure support provided by the NIHR Imperial Biomedical Research Centre (BRC). The national cohorts are supported by: Danish Cancer Society (Denmark); Ligue Contre le Cancer, Institut Gustave Roussy, Mutuelle Générale de l’Education Nationale, Institut National de la Santé et de la Recherche Médicale (INSERM) (France); German Cancer Aid, German Cancer Research Center (DKFZ), German Institute of Human Nutrition Potsdam-Rehbruecke (DIfE), Federal Ministry of Education and Research (BMBF) (Germany); Associazione Italiana per la Ricerca sul Cancro-AIRC-Italy, Compagnia di SanPaolo and National Research Council (Italy); Dutch Ministry of Public Health, Welfare and Sports (VWS), LK Research Funds, Dutch Prevention Funds, Dutch ZON (Zorg Onderzoek Nederland), World Cancer Research Fund (WCRF) (The Netherlands), and Statistics Netherlands is acknowledged for providing the causes of death; Health Research Fund (FIS) - Instituto de Salud Carlos III (ISCIII), Regional Governments of Andalucía, Asturias, Basque Country, Murcia and Navarra, and the Catalan Institute of Oncology - ICO (Spain); Swedish Cancer Society, Swedish Research Council and County Councils of Skåne and Västerbotten and the funds supporting the Northern Sweden Diet Database (Sweden); Cancer Research UK (14136 to EPIC-Norfolk; C8221/A29017 to EPIC-Oxford), Medical Research Council (1000143 to EPIC-Norfolk; MR/M012190/1 to EPIC-Oxford) (United Kingdom). The funders had no role in the design of the study; in the collection, analyses, or interpretation of data; in the writing of the manuscript; or in the decision to publish the results.

## Notes

### Competing Interest Statement

The authors have declared no competing interest.

### Author Declarations

All participants provided written informed consent, and the study protocol was approved by the ethical review board of IARC and the institutions where the participants were recruited

