## Supplementary materials for "Dietary amino acids and risk of stroke subtypes: a prospective analysis of 356,000 participants in seven European countries"

**Supplementary tables**

**Supplementary figures**

[Supplementary figure 3: Spearman correlation coefficients of dietary amino acids (g/day) with **dietary protein from different sources**	49](#_Toc124438230)[_Toc124438231](#_Toc124438231)

Supplementary table 1: Participant characteristics of stroke cases at recruitment by sex in the EPIC study

| Participant characteristics |  | Men | | |  |  | Women | | |
| --- | --- | --- | --- | --- | --- | --- | --- | --- | --- |
|  | All | | Ischaemic stroke cases | Haemorrhagic stroke cases | | All | | Ischaemic stroke cases | Haemorrhagic stroke cases |
| Number of participants | 129,005 | | 2235 | 565 | | 227,137 | | 2060 | 810 |
| Age in years, mean (SD) | 52.1 (9.9) | | 58.7 (7.5) | 57.6 (8.3) | | 50.6 (10.8) | | 60.1 (8.5) | 57.1 (9.6) |
| Smoking status, n (%) |  | |  |  | |  | |  |  |
| Never smoker | 44,097 (34.2%) | | 634 (28.4%) | 164 (29.0%) | | 123,144 (54.2%) | | 997 (48.4%) | 358 (44.2%) |
| Former smoker | 46,936 (36.4%) | | 784 (35.1%) | 225 (39.8%) | | 54,216 (23.9%) | | 441 (21.4%) | 174 (21.5%) |
| Current smoker, <10 or number unknown | 13,518 (10.5%) | | 252 (11.3%) | 52 (9.2%) | | 15,488 (6.8%) | | 144 (7.0%) | 63 (7.8%) |
| Current smoker, 10-19 | 9,704 (7.5%) | | 233 (10.4%) | 55 (9.7%) | | 19,354 (8.5%) | | 283 (13.7%) | 122 (15.1%) |
| Current smoker, ≥20 | 13,339 (10.3%) | | 314 (14.0%) | 67 (11.9%) | | 13,445 (5.9%) | | 181 (8.8%) | 87 (10.7%) |
| Unknown | 1,411 (1.1%) | | 18 (0.8%) | 2 (0.4%) | | 1,490 (0.7%) | | 14 (0.7%) | 6 (0.7%) |
| Cambridge physical activity index, n (%) |  | |  |  | |  | |  |  |
| Inactive | 22,598 (17.5%) | | 537 (24.0%) | 129 (22.8%) | | 53,398 (23.5%) | | 675 (32.8%) | 227 (28.0%) |
| Moderately inactive | 40,102 (31.1%) | | 703 (31.5%) | 196 (34.7%) | | 78,660 (34.6%) | | 703 (34.1%) | 285 (35.2%) |
| Moderately active | 31,220 (24.2%) | | 525 (23.5%) | 123 (21.8%) | | 49,472 (21.8%) | | 380 (18.4%) | 160 (19.8%) |
| Active | 32,064 (24.9%) | | 436 (19.5%) | 108 (19.1%) | | 41,632 (18.3%) | | 273 (13.3%) | 127 (15.7%) |
| Unknown | 3,021 (2.3%) | | 34 (1.5%) | 9 (1.6%) | | 3,975 (1.8%) | | 29 (1.4%) | 11 (1.4%) |
| Employed or student, n (%) |  | |  |  | |  | |  |  |
| Yes | 85,841 (66.5%) | | 1,138 (50.9%) | 306 (54.2%) | | 124,753 (54.9%) | | 781 (37.9%) | 393 (48.5%) |
| No | 27,275 (21.1%) | | 810 (36.2%) | 194 (34.3%) | | 74,742 (32.9%) | | 1,081 (52.5%) | 343 (42.3%) |
| Unknown | 15,889 (12.3%) | | 287 (12.8%) | 65 (11.5%) | | 27,642 (12.2%) | | 198 (9.6%) | 74 (9.1%) |
| Level of education reached, n (%) |  | |  |  | |  | |  |  |
| None or primary | 40,596 (31.5%) | | 1,066 (47.7%) | 226 (40.0%) | | 72,112 (31.7%) | | 954 (46.3%) | 299 (36.9%) |
| Secondary | 17,276 (13.4%) | | 215 (9.6%) | 68 (12.0%) | | 34,849 (15.3%) | | 192 (9.3%) | 117 (14.4%) |
| Vocational or university | 67,344 (52.2%) | | 914 (40.9%) | 247 (43.7%) | | 109,160 (48.1%) | | 817 (39.7%) | 340 (42.0%) |
| Unknown | 3,789 (2.9%) | | 40 (1.8%) | 24 (4.2%) | | 72,112 (31.7%) | | 97 (4.7%) | 54 (6.7%) |
| Alcohol consumption, n (%) |  | |  |  | |  | |  |  |
| Non-drinkers | 7,054 (5.5%) | | 211 (9.4%) | 39 (6.9%) | | 40,872 (15.5%) | | 451 (21.9%) | 141 (17.4%) |
| 0.1-4.9 g/day | 26,875 (20.8%) | | 401 (17.9%) | 112 (19.8%) | | 107,156 (40.6%) | | 741 (36.0%) | 298 (36.8%) |
| 5.0-14.9 g/day | 34,087 (26.4%) | | 570 (25.5%) | 142 (25.1%) | | 72,758 (27.6%) | | 519 (25.2%) | 211 (26.0%) |
| 15.0-29.9 g/day | 27,250 (21.1%) | | 463 (20.7%) | 105 (18.6%) | | 27,313 (10.3%) | | 209 (10.1%) | 91 (11.2%) |
| 30.0-59.9 g/day | 24,137 (18.7%) | | 400 (17.9%) | 112 (19.8%) | | 13,868 (5.3%) | | 125 (6.1%) | 60 (7.4%) |
| ≥60.0 g/day | 9,602 (7.4%) | | 190 (8.5%) | 55 (9.7%) | | 1,946 (0.7%) | | 15 (0.7%) | 9 (1.1%) |
| Alcohol consumption, g/day, median (25^th^, 75^th^ percentile) | 13.8 (4.5, 31.4) | | 13.7 (4.1, 31.7) | 13.7 (4.4, 35.4) | | 4.0 (0.6, 12.4) | | 2.9 (0.2, 11.1) | 4.0 (0.4, 12.9) |
| BMI |  | |  |  | |  | |  |  |
| <22.5 kg/m^2^ | 15,193 (11.8%) | | 209 (9.4%) | 61 (10.8%) | | 72,797 (27.6%) | | 366 (17.8%) | 218 (26.9%) |
| 22.5-24.9 kg/m^2^ | 31,140 (24.1%) | | 450 (20.1%) | 112 (19.8%) | | 68,923 (26.1%) | | 454 (22.0%) | 192 (23.7%) |
| 25.0-27.4 kg/m^2^ | 37,956 (29.4%) | | 641 (28.7%) | 168 (29.7%) | | 51,969 (19.7%) | | 462 (22.4%) | 165 (20.4%) |
| 27.5-29.9 kg/m^2^ | 24,999 (19.4%) | | 517 (23.1%) | 116 (20.5%) | | 31,418 (11.9%) | | 327 (15.9%) | 111 (13.7%) |
| ≥30.0 kg/m^2^ | 18,966 (14.7%) | | 412 (18.4%) | 107 (18.9%) | | 36,447 (13.8%) | | 441 (21.4%) | 119 (14.7%) |
| Unknown | 751 (0.6%) | | 6 (0.3%) | 1 (0.2%) | | 2,359 (0.9%) | | 10 (0.5%) | 5 (0.6%) |
| BMI, kg/m^2^, mean (SD) | 26.5 (3.6) | | 27.0 (3.7) | 27.1 (4.1) | | 25.5 (4.5) | | 26.7 (4.7) | 25.5 (4.3) |
| History of diabetes |  | |  |  | |  | |  |  |
| No | 122,090 (94.6%) | | 1,961 (87.7%) | 509 (90.1%) | | 214,965 (94.6%) | | 1,797 (87.2%) | 746 (92.1%) |
| Yes | 3,981 (3.1%) | | 174 (7.8%) | 24 (4.2%) | | 4,668 (2.1%) | | 149 (7.2%) | 21 (2.6%) |
| Unknown | 2,934 (2.3%) | | 100 (4.5%) | 32 (5.7%) | | 7,504 (3.3%) | | 114 (5.5%) | 43 (5.3%) |
| History of hypertension |  | |  |  | |  | |  |  |
| No | 98,429 (76.3%) | | 1,441 (64.5%) | 366 (64.8%) | | 177,787 (78.3%) | | 1,250 (60.7%) | 541 (66.8%) |
| Yes | 24,828 (19.2%) | | 700 (31.3%) | 168 (29.7%) | | 41,442 (18.2%) | | 761 (36.9%) | 230 (28.4%) |
| Unknown | 5,748 (4.5%) | | 94 (4.2%) | 31 (5.5%) | | 7,908 (3.5%) | | 49 (2.4%) | 39 (4.8%) |
| History of hyperlipidaemia |  | |  |  | |  | |  |  |
| No | 84,665 (65.6%) | | 1,076 (48.1%) | 306 (54.2%) | | 166,136 (73.1%) | | 993 (48.2%) | 492 (60.7%) |
| Yes | 21,093 (16.4%) | | 284 (12.7%) | 76 (13.5%) | | 24,149 (10.6%) | | 260 (12.6%) | 72 (8.9%) |
| Unknown | 23,247 (18.0%) | | 875 (39.1%) | 183 (32.4%) | | 36,852 (16.2%) | | 807 (39.2%) | 246 (30.4%) |

Supplementary table 2: Mean (SD) intakes of dietary amino acids, protein and total energy at recruitment in the EPIC study

|  | Men | | | | | | | | | | | |  | | Women | | | | | | | | | | | | |
| --- | --- | --- | --- | --- | --- | --- | --- | --- | --- | --- | --- | --- | --- | --- | --- | --- | --- | --- | --- | --- | --- | --- | --- | --- | --- | --- | --- |
| Dietary amino acid |  | g/day | |  |  | g/1000kcal | |  |  | | % of total protein | | |  | |  | g/day | |  |  | g/1000kcal | | |  |  | % of total protein | |
|  | observed | | calibrated | | observed | | calibrated | | | observed | | calibrated |  | | observed | | | calibrated | observed | | | calibrated | observed | | | | calibrated |
| Branched-chain amino acids | 13.9 (4.5) | | 13.7 (2.8) | | 5.7 (1.3) | | 5.3 (0.9) | | | 15.5(1.8) | | 15.2 (1.7) |  | | 11.7 (3.8) | | | 10.3 (2.0) | 6.0 (1.4) | | | 5.4 (1.0) | 15.6 (1.8) | | | | 15.2 (1.7) |
| Isoleucine | 3.5 (1.2) | | 3.5 (0.7) | | 1.5 (0.3) | | 1.3 (0.2) | | | 3.9 (0.4) | | 3.8 (0.4) |  | | 3.0 (1.0) | | | 2.6 (0.5) | 1.5 (0.4) | | | 1.4 (0.3) | 3.9 (0.4) | | | | 3.8 (0.4) |
| Leucine | 6.2 (2.0) | | 6.1 (1.2) | | 2.6 (0.6) | | 2.4 (0.4) | | | 6.9 (0.8) | | 6.8 (0.8) |  | | 5.2 (1.7) | | | 4.6 (0.9) | 2.7 (0.6) | | | 2.4 (0.5) | 6.9 (0.8) | | | | 6.8 (0.8) |
| Valine | 4.2 (1.3) | | 4.1 (0.8) | | 1.7 (0.4) | | 1.6 (0.3) | | | 4.7 (0.6) | | 4.6 (0.5) |  | | 3.6 (1.1) | | | 3.1 (0.6) | 1.8 (0.4) | | | 1.6 (0.3) | 4.7 (0.6) | | | | 4.6 (0.5) |
| Other essential amino acids | 17.1 (5.6) | | 16.8 (3.6) | | 7.0 (1.6) | | 6.5 (1.2) | | | 18.9 (2.2) | | 18.7 (2.2) |  | | 14.3 (4.7) | | | 12.5 (2.5) | 7.3 (1.8) | | | 6.6 (1.3) | 19.0 (2.3) | | | | 18.5 (2.2) |
| Histidine | 2.3 (0.7) | | 2.2 (0.5) | | 0.9 (0.2) | | 0.9 (0.2) | | | 2.5 (0.3) | | 2.5 (0.3) |  | | 1.9 (0.6) | | | 1.6 (0.3) | 1.0 (0.2) | | | 0.9 (0.2) | 2.5 (0.3) | | | | 2.4 (0.3) |
| Lysine | 5.5 (1.9) | | 5.4 (1.3) | | 2.3 (0.6) | | 2.1 (0.4) | | | 6.1 (0.9) | | 6.0 (0.8) |  | | 4.6 (1.6) | | | 4.0 (0.9) | 2.4 (0.6) | | | 2.1 (0.5) | 6.1 (0.9) | | | | 5.9 (0.9) |
| Methionine | 1.8 (0.6) | | 1.8 (0.4) | | 0.7 (0.2) | | 0.7 (0.1) | | | 2.0 (0.3) | | 2.0 (0.3) |  | | 1.5 (0.5) | | | 1.3 (0.3) | 0.8 (0.2) | | | 0.7 (0.1) | 2.0 (0.3) | | | | 1.9 (0.3) |
| Phenylalanine | 3.6 (1.1) | | 3.5 (0.7) | | 1.5 (0.3) | | 1.4 (0.2) | | | 4.0 (0.5) | | 3.9 (0.4) |  | | 3.0 (0.9) | | | 2.6 (0.5) | 1.5 (0.3) | | | 1.4 (0.2) | 4.0 (0.4) | | | | 3.9 (0.4) |
| Threonine | 3.1 (1.0) | | 3.0 (0.6) | | 1.3 (0.3) | | 1.2 (0.2) | | | 3.4 (0.4) | | 3.4 (0.4) |  | | 2.6 (0.8) | | | 2.3 (0.4) | 1.3 (0.3) | | | 1.2 (0.2) | 3.4 (0.4) | | | | 3.4 (0.4) |
| Tryptophan | 0.9 (0.3) | | 0.9 (0.2) | | 0.4 (0.1) | | 0.4 (0.1) | | | 1.0 (0.1) | | 1.0 (0.1) |  | | 0.8 (0.2) | | | 0.7 (0.1) | 0.4 (0.1) | | | 0.4 (0.1) | 1.0 (0.1) | | | | 1.0 (0.1) |
| Non-essential amino acids | 46.6 (14.5) | | 46.4 (8.8) | | 19.2(3.9) | | 18.0 (2.8) | | | 51.9 (5.9) | | 51.9 (5.6) |  | | 38.7 (11.9) | | | 34.6 (5.9) | 19.9 (4.1) | | | 18.4 (3.0) | 51.7 (5.7) | | | | 51.6 (5.2) |
| Alanine | 3.7 (1.2) | | 3.6 (0.8) | | 1.5 (0.4) | | 1.4 (0.3) | | | 4.1 (0.5) | | 4.0 (0.5) |  | | 3.0 (1.0) | | | 2.6 (0.5) | 1.5 (0.4) | | | 1.4 (0.3) | 3.9 (0.5) | | | | 3.9 (0.5) |
| Arginine | 4.1 (1.3) | | 4.1 (0.8) | | 1.7 (0.4) | | 1.6 (0.3) | | | 4.5 (0.6) | | 4.5 (0.4) |  | | 3.3 (1.1) | | | 3.0 (0.5) | 1.7 (0.4) | | | 1.6 (0.3) | 4.4 (0.6) | | | | 4.4 (0.4) |
| Aspartic acid | 7.1 (2.2) | | 6.9 (1.3) | | 2.9 (0.6) | | 2.7 (0.4) | | | 7.9 (0.9) | | 7.7 (0.6) |  | | 6.0 (1.8) | | | 5.2 (0.9) | 3.1 (0.7) | | | 2.8 (0.5) | 8.0 (0.9) | | | | 7.8 (0.6) |
| Cystine | 1.0 (0.3) | | 1.0 (0.2) | | 0.4 (0.1) | | 0.4 (0.1) | | | 1.1 (0.2) | | 1.1 (0.2) |  | | 0.8 (0.2) | | | 0.7 (0.1) | 0.4 (0.1) | | | 0.4 (0.1) | 1.1 (0.2) | | | | 1.1 (0.1) |
| Glutamic acid | 15.5 (4.9) | | 15.6 (3.1) | | 6.4 (1.3) | | 6.1 (1.0) | | | 17.3 (2.6) | | 17.5 (2.4) |  | | 12.9 (4.0) | | | 11.8 (2.0) | 6.6 (1.4) | | | 6.2 (1.0) | 17.3 (2.3) | | | | 17.6 (2.1) |
| Glycine | 3.1 (1.1) | | 3.1 (0.7) | | 1.3 (0.3) | | 1.2 (0.2) | | | 3.5 (0.5) | | 3.5 (0.4) |  | | 2.4 (0.8) | | | 2.2 (0.4) | 1.3 (0.3) | | | 1.2 (0.2) | 3.3 (0.5) | | | | 3.3 (0.4) |
| Hydroxyproline | 0.09 (0.07) | | 0.09 (0.04) | | 0.04 (0.03) | | 0.04 (0.01) | | | 0.1 (0.06) | | 0.1 (0.04) |  | | 0.06 (0.05) | | | 0.05 (0.03) | 0.03 (0.02) | | | 0.03 (0.01) | 0.08 (0.06) | | | | 0.08 (0.04) |
| Proline | 5.5 (1.9) | | 5.5 (1.2) | | 2.3 (0.5) | | 2.1 (0.4) | | | 6.2 (1.2) | | 6.2 (1.0) |  | | 4.7 (1.6) | | | 4.2 (0.8) | 2.4 (0.6) | | | 2.2 (0.4) | 6.3 (1.1) | | | | 6.3 (0.9) |
| Serine | 3.7 (1.2) | | 3.7 (0.7) | | 1.5 (0.3) | | 1.4 (0.2) | | | 4.1 (0.5) | | 4.1 (0.5) |  | | 3.1 (1.0) | | | 2.8 (0.5) | 1.6 (0.3) | | | 1.5 (0.3) | 4.2 (0.5) | | | | 4.1 (0.4) |
| Tyrosine | 2.8 (0.9) | | 2.8 (0.6) | | 1.2 (0.3) | | 1.1 (0.2) | | | 3.1 (0.4) | | 3.1 (0.4) |  | | 2.4 (0.8) | | | 2.1 (0.4) | 1.2 (0.3) | | | 1.1 (0.2) | 3.2 (0.4) | | | | 3.1 (0.4) |
| Protein | 90.0 (26.6) | | 89.4(14.2) | | 37.0(6.1) | | 34.8 (3.9) | | | --- | | --- |  | | 74.9 (21.4) | | | 67.0 (9.0) | 38.4 (6.5) | | | 35.5 (4.0) | --- | | | | --- |
| Energy (kcal) | 2445 (635) | | 2577(316) | | --- | | --- | | | --- | | --- |  | | 1966 (517) | | | 1897(236) | --- | | | --- | --- | | | | --- |

Supplementary table 3: Intake of dietary amino acids, protein and total energy (medians and 25^th^, 75^th^ percentile of g or kcal per day) at recruitment by country in the EPIC study

|  | Denmark | Germany | Italy | Netherlands | Spain | Sweden | UK |
| --- | --- | --- | --- | --- | --- | --- | --- |
|  | N=53,617 | N=50,295 | N=44,760 | N=37,351 | N=40,062 | N=51,592 | N=78,465 |
| Branched-chain amino acids (g) | 13.9 (11.3, 16.9) | 10.8 (8.8, 13.3) | 14.2 (11.4, 17.2) | 11.7 (9.7, 14.1) | 13.5 (11.0, 16.4) | 11.9 (9.5, 14.7) | 10.4 (8.2, 12.7) |
| Isoleucine (g) | 3.5 (2.9, 4.3) | 2.7 (2.2, 3.3) | 3.5 (2.9, 4.3) | 2.9 (2.4, 3.5) | 3.5 (2.9, 4.3) | 2.9 (2.3, 3.6) | 2.6 (2.1, 3.2) |
| Leucine (g) | 6.2 (5.0, 7.5) | 4.8 (3.9, 5.9) | 6.4 (5.1, 7.8) | 5.2 (4.3, 6.3) | 5.9 (4.8, 7.2) | 5.3 (4.2, 6.6) | 4.6 (3.6, 5.6) |
| Valine (g) | 4.2 (3.4, 5.1) | 3.3 (2.7, 4.0) | 4.2 (3.4, 5.1) | 3.6 (2.9, 4.3) | 4.0 (3.3, 4.9) | 3.6 (2.9, 4.5) | 3.2 (2.5, 3.9) |
| Other essential amino acids (g) | 16.9 (13.7, 20.6) | 13.1 (10.6, 16.1) | 17.5 (14.1, 21.3) | 14.3 (11.8, 17.1) | 16.9 (13.7, 20.5) | 14.6 (11.5, 18.1) | 12.5 (9.7, 15.5) |
| Histidine (g) | 2.2 (1.8, 2.7) | 1.7 (1.4, 2.1) | 2.3 (1.9, 2.8) | 1.9 (1.6, 2.3) | 2.2 (1.8, 2.7) | 1.9 (1.5, 2.4) | 1.6 (1.2, 2.0) |
| Lysine (g) | 5.4 (4.4, 6.6) | 4.1 (3.2, 5.0) | 5.6 (4.5, 6.9) | 4.7 (3.9, 5.7) | 5.7 (4.6, 6.9) | 4.7 (3.7, 5.9) | 3.9 (2.9, 5.0) |
| Methionine (g) | 1.8 (1.5, 2.2) | 1.4 (1.1, 1.7) | 1.8 (1.5, 2.2) | 1.5 (1.2, 1.8) | 1.7 (1.4, 2.1) | 1.5 (1.2, 1.9) | 1.3 (1.0, 1.6) |
| Phenylalanine (g) | 3.5 (2.9, 4.3) | 2.8 (2.3, 3.5) | 3.6 (2.9, 4.4) | 3.0 (2.4, 3.6) | 3.4 (2.7, 4.1) | 3.0 (2.4, 3.8) | 2.6 (2.1, 3.2) |
| Threonine (g) | 3.1 (2.5, 3.7) | 2.3 (1.9, 2.9) | 3.1 (2.5, 3.8) | 2.5 (2.1, 3.0) | 3.1 (2.5, 3.7) | 2.6 (2.0, 3.2) | 2.3 (1.8, 2.9) |
| Tryptophan (g) | 0.9 (0.7, 1.1) | 0.7 (0.6, 0.9) | 1.0 (0.8, 1.2) | 0.7 (0.6, 0.9) | 0.9 (0.7, 1.1) | 0.8 (0.6, 0.9) | 0.7 (0.6, 0.9) |
| Non-essential amino acids (g) | 46.8 (38.3, 56.5) | 37.1 (30.2, 45.2) | 47.5 (38.5, 57.5) | 38.4 (31.9, 46.0) | 43.3 (35.5, 52.1) | 39.7 (31.7, 48.8) | 34.2 (27.5, 41.5) |
| Alanine (g) | 3.7 (3.0, 4.5) | 2.8 (2.2, 3.4) | 3.6 (2.9, 4.3) | 2.9 (2.4, 3.5) | 3.7 (3.0, 4.5) | 2.9 (2.3, 3.7) | 2.6 (2.0, 3.3) |
| Arginine (g) | 4.0 (3.3, 4.8) | 3.1 (2.5, 3.8) | 4.0 (3.2, 4.8) | 3.4 (2.8, 4.0) | 4.2 (3.4, 5.1) | 3.2 (2.5, 4.0) | 3.0 (2.3, 3.7) |
| Aspartic acid (g) | 7.1 (5.8, 8.5) | 5.2 (4.3, 6.4) | 6.8 (5.5, 8.2) | 6.0 (5.0, 7.1) | 7.3 (6.0, 8.8) | 5.9 (4.7, 7.3) | 5.7 (4.6, 6.9) |
| Cystine (g) | 1.0 (0.8, 1.2) | 0.8 (0.7, 1.0) | 0.9 (0.7, 1.1) | 0.8 (0.6, 0.9) | 0.9 (0.7, 1.1) | 0.8 (0.7, 1.0) | 0.6 (0.5, 0.8) |
| Glutamic acid (g) | 15.6 (12.7, 18.8) | 12.7 (10.4, 15.5) | 16.4 (13.2, 20.0) | 12.5 (10.4, 15.1) | 13.5 (11.1, 16.2) | 13.5 (10.8, 16.6) | 11.2 (9.1, 13.5) |
| Glycine (g) | 3.2 (2.6, 3.9) | 2.4 (1.9, 3.0) | 2.9 (2.3, 3.5) | 2.5 (2.0, 3.0) | 3.0 (2.4, 3.7) | 2.5 (1.9, 3.1) | 2.1 (1.6, 2.7) |
| Hydroxyproline (g) | 0.1 (0.1, 0.1) | 0.1 (0.0, 0.1) | 0.1 (0.0, 0.1) | 0.1 (0.1, 0.2) | 0.0 (0.0, 0.1) | 0.1 (0.0, 0.1) | 0.0 (0.0, 0.0) |
| Proline (g) | 5.5 (4.5, 6.8) | 4.6 (3.7, 5.7) | 6.1 (4.9, 7.5) | 4.7 (3.8, 5.7) | 4.5 (3.6, 5.5) | 5.0 (3.9, 6.2) | 4.0 (3.2, 4.8) |
| Serine (g) | 3.7 (3.0, 4.5) | 3.0 (2.4, 3.6) | 3.8 (3.1, 4.6) | 3.2 (2.6, 3.8) | 3.5 (2.9, 4.2) | 3.2 (2.6, 4.0) | 2.7 (2.2, 3.3) |
| Tyrosine (g) | 2.8 (2.3, 3.5) | 2.2 (1.8, 2.7) | 2.9 (2.3, 3.6) | 2.5 (2.0, 3.0) | 2.6 (2.1, 3.2) | 2.5 (1.9, 3.1) | 2.1 (1.6, 2.6) |
| Protein (g) | 82.0 (67.4, 98.7) | 70.6 (57.7, 86.0) | 85.7 (69.8, 103.0) | 81.1 (68.0, 96.0) | 90.6 (74.1, 110.1) | 73.4 (57.9, 90.8) | 72.0 (59.1, 86.2) |
| Energy (kcal) | 2166 (1788, 2597) | 2039 (1675, 2481) | 2205 (1801, 2652) | 2089 (1756, 2497) | 2110 (1702, 2604) | 1982 (1604, 2435) | 2016 (1664, 2417) |

Supplementary table 4: Hazard ratios (95% confidence intervals)^1^ for **ischaemic stroke** (4295 cases) by increments and fifths of **observed intakes** of dietary amino acids.

| Amino acids | Per SD of observed intakes | |  | Fifths of observed intakes compared to bottom fifth, HR(95% CI) | | | |  |  |
| --- | --- | --- | --- | --- | --- | --- | --- | --- | --- |
|  | HR (95% CI) | p-trend^2^ |  | 2 | 3 | 4 | 5 | p-trend^3^ | p for non-linearity^4^ |
| Branched-chain amino acids |  |  |  |  |  |  |  |  |  |
| Percent of total protein | 0.97 (0.95, 0.99) | 0.02 |  | 0.97 (0.88, 1.08) | 0.96 (0.86, 1.06) | 1.00 (0.90, 1.12) | 0.89 (0.79, 0.99) | 0.12 | 0.53 |
| g/day | 0.94 (0.89, 0.99) | 0.02 |  | 0.98 (0.88, 1.08) | 0.91 (0.82, 1.02) | 0.95 (0.84, 1.07) | 0.92 (0.80, 1.07) | 0.34 | 1.00 |
| g/1000kcal | 0.98 (0.97, 1.00) | 0.02 |  | 0.98 (0.88, 1.08) | 0.91 (0.82, 1.01) | 0.96 (0.86, 1.06) | 0.89 (0.80, 1.00) | 0.04 | 0.92 |
| Percent of total amino acids | 0.99 (0.99, 1.00) | 0.02 |  | 1.00 (0.91, 1.10) | 0.95 (0.86, 1.05) | 0.94 (0.85, 1.04) | 0.91 (0.82, 1.01) | 0.04 | 1.00 |
| Isoleucine |  |  |  |  |  |  |  |  |  |
| Percent of total protein | 0.98 (0.96, 1.00) | 0.02 |  | 1.03 (0.93, 1.14) | 0.93 (0.84, 1.03) | 0.98 (0.88, 1.09) | 0.93 (0.83, 1.04) | 0.17 | 0.94 |
| g/day | 0.94 (0.90, 0.99) | 0.02 |  | 0.94 (0.85, 1.04) | 0.92 (0.83, 1.03) | 0.91 (0.81, 1.03) | 0.92 (0.80, 1.06) | 0.32 | 1.00 |
| g/1000kcal | 0.98 (0.97, 1.00) | 0.02 |  | 0.99 (0.89, 1.09) | 0.92 (0.83, 1.02) | 0.95 (0.86, 1.06) | 0.91 (0.81, 1.02) | 0.08 | 1.00 |
| Percent of total amino acids | 0.99 (0.98, 1.00) | 0.02 |  | 0.93 (0.85, 1.03) | 0.93 (0.85, 1.03) | 0.98 (0.88, 1.09) | 0.92 (0.80, 1.05) | 0.35 | 1.00 |
| Leucine |  |  |  |  |  |  |  |  |  |
| Percent of total protein | 0.97 (0.95, 1.00) | 0.02 |  | 0.94 (0.84, 1.04) | 1.00 (0.90, 1.11) | 0.98 (0.87, 1.09) | 0.90 (0.80, 1.02) | 0.23 | 0.90 |
| g/day | 0.94 (0.90, 0.99) | 0.02 |  | 0.95 (0.86, 1.06) | 0.93 (0.83, 1.04) | 0.93 (0.82, 1.04) | 0.92 (0.80, 1.06) | 0.29 | 1.00 |
| g/1000kcal | 0.98 (0.97, 1.00) | 0.02 |  | 0.96 (0.87, 1.07) | 0.91 (0.82, 1.01) | 0.95 (0.85, 1.05) | 0.89 (0.80, 0.99) | 0.04 | 0.93 |
| Percent of total amino acids | 0.99 (0.99, 1.00) | 0.02 |  | 0.89 (0.81, 0.99) | 0.94 (0.85, 1.03) | 0.89 (0.80, 0.98) | 0.88 (0.79, 0.97) | 0.02 | 0.34 |
| Valine |  |  |  |  |  |  |  |  |  |
| Percent of total protein | 0.97 (0.95, 0.99) | 0.009* |  | 0.97 (0.88, 1.08) | 0.97 (0.87, 1.07) | 0.97 (0.87, 1.09) | 0.92 (0.82, 1.03) | 0.19 | 1.00 |
| g/day | 0.94 (0.89, 0.98) | 0.009 |  | 0.97 (0.87, 1.07) | 0.90 (0.81, 1.01) | 0.95 (0.85, 1.07) | 0.89 (0.78, 1.03) | 0.16 | 1.00 |
| g/1000kcal | 0.98 (0.96, 0.99) | 0.01 |  | 0.96 (0.86, 1.06) | 0.91 (0.82, 1.01) | 0.92 (0.83, 1.03) | 0.87 (0.78, 0.97) | 0.01 | 0.76 |
| Percent of total amino acids | 0.99 (0.98, 1.00) | 0.009 |  | 0.98 (0.89, 1.07) | 0.90 (0.81, 0.99) | 0.88 (0.79, 0.97) | 0.87 (0.78, 0.96) | 0.001 | 0.14 |
| Other essential amino acids |  |  |  |  |  |  |  |  |  |
| Percent of total protein | 0.98 (0.96, 1.00) | 0.05 |  | 1.01 (0.91, 1.13) | 0.96 (0.86, 1.07) | 0.95 (0.85, 1.06) | 0.98 (0.87, 1.10) | 0.49 | 1.00 |
| g/day | 0.95 (0.90, 1.00) | 0.05 |  | 0.96 (0.86, 1.06) | 0.92 (0.82, 1.03) | 0.93 (0.83, 1.05) | 0.90 (0.78, 1.03) | 0.17 | 1.00 |
| g/1000kcal | 0.98 (0.97, 1.00) | 0.05 |  | 0.96 (0.87, 1.07) | 0.88 (0.79, 0.98) | 0.94 (0.84, 1.04) | 0.90 (0.81, 1.01) | 0.09 | 0.30 |
| Percent of total amino acids | 0.99 (0.98, 1.00) | 0.05 |  | 1.01 (0.91, 1.13) | 1.03 (0.92, 1.16) | 1.02 (0.90, 1.14) | 1.09 (0.96, 1.23) | 0.20 | 1.00 |
| Histidine |  |  |  |  |  |  |  |  |  |
| Percent of total protein | 0.98 (0.96, 1.00) | 0.11 |  | 0.98 (0.88, 1.09) | 0.95 (0.85, 1.06) | 0.96 (0.85, 1.07) | 1.00 (0.89, 1.12) | 0.96 | 1.00 |
| g/day | 0.96 (0.91, 1.01) | 0.11 |  | 0.94 (0.85, 1.05) | 0.95 (0.85, 1.06) | 0.92 (0.81, 1.04) | 0.92 (0.80, 1.06) | 0.31 | 1.00 |
| g/1000kcal | 0.99 (0.97, 1.00) | 0.11 |  | 0.98 (0.88, 1.09) | 0.94 (0.84, 1.05) | 0.98 (0.88, 1.09) | 0.93 (0.83, 1.04) | 0.27 | 1.00 |
| Percent of total amino acids | 0.99 (0.98, 1.00) | 0.11 |  | 0.97 (0.87, 1.09) | 1.01 (0.90, 1.12) | 1.11 (1.00, 1.24) | 1.12 (1.00, 1.25) | 0.004 | 0.02 |
| Lysine |  |  |  |  |  |  |  |  |  |
| Percent of total protein | 0.98 (0.96, 1.00) | 0.08 |  | 1.03 (0.92, 1.15) | 0.98 (0.87, 1.10) | 0.92 (0.82, 1.03) | 0.99 (0.88, 1.11) | 0.39 | 0.42 |
| g/day | 0.96 (0.91, 1.01) | 0.08 |  | 0.99 (0.89, 1.10) | 0.93 (0.84, 1.05) | 0.95 (0.84, 1.07) | 0.93 (0.81, 1.07) | 0.29 | 1.00 |
| g/1000kcal | 0.99 (0.97, 1.00) | 0.08 |  | 0.98 (0.88, 1.09) | 0.92 (0.82, 1.03) | 0.95 (0.85, 1.06) | 0.93 (0.83, 1.04) | 0.19 | 0.96 |
| Percent of total amino acids | 0.98 (0.97, 1.00) | 0.08 |  | 1.00 (0.89, 1.12) | 1.01 (0.90, 1.13) | 1.02 (0.91, 1.15) | 1.04 (0.92, 1.18) | 0.48 | 1.00 |
| Methionine |  |  |  |  |  |  |  |  |  |
| Percent of total protein | 0.98 (0.95, 1.00) | 0.07 |  | 1.05 (0.94, 1.17) | 1.01 (0.90, 1.13) | 0.97 (0.86, 1.09) | 1.02 (0.90, 1.15) | 0.80 | 1.00 |
| g/day | 0.96 (0.91, 1.00) | 0.07 |  | 0.95 (0.86, 1.06) | 0.92 (0.82, 1.03) | 0.94 (0.83, 1.06) | 0.89 (0.77, 1.03) | 0.16 | 1.00 |
| g/1000kcal | 0.98 (0.97, 1.00) | 0.07 |  | 0.96 (0.86, 1.07) | 0.91 (0.82, 1.01) | 0.95 (0.85, 1.05) | 0.90 (0.81, 1.01) | 0.09 | 0.72 |
| Percent of total amino acids | 0.99 (0.97, 1.00) | 0.07 |  | 0.96 (0.85, 1.07) | 1.02 (0.91, 1.14) | 0.92 (0.82, 1.04) | 1.03 (0.91, 1.15) | 0.71 | 0.23 |
| Phenylalanine |  |  |  |  |  |  |  |  |  |
| Percent of total protein | 0.97 (0.95, 0.99) | 0.02 |  | 0.98 (0.89, 1.09) | 1.03 (0.93, 1.15) | 1.01 (0.90, 1.12) | 0.90 (0.79, 1.02) | 0.30 | 0.66 |
| g/day | 0.94 (0.89, 0.99) | 0.02 |  | 0.98 (0.89, 1.08) | 0.91 (0.81, 1.01) | 0.95 (0.84, 1.07) | 0.90 (0.78, 1.04) | 0.17 | 1.00 |
| g/1000kcal | 0.98 (0.97, 1.00) | 0.02 |  | 0.91 (0.82, 1.01) | 0.94 (0.85, 1.05) | 0.89 (0.80, 0.98) | 0.89 (0.80, 0.99) | 0.03 | 0.72 |
| Percent of total amino acids | 0.99 (0.98, 1.00) | 0.02 |  | 1.03 (0.94, 1.12) | 0.96 (0.87, 1.05) | 0.87 (0.79, 0.97) | 0.93 (0.83, 1.05) | 0.03 | 0.14 |
| Threonine |  |  |  |  |  |  |  |  |  |
| Percent of total protein | 0.98 (0.96, 1.00) | 0.04 |  | 0.99 (0.89, 1.10) | 0.99 (0.89, 1.10) | 0.91 (0.81, 1.02) | 1.01 (0.90, 1.13) | 0.60 | 0.64 |
| g/day | 0.95 (0.90, 1.00) | 0.04 |  | 0.93 (0.84, 1.03) | 0.93 (0.83, 1.04) | 0.91 (0.80, 1.02) | 0.89 (0.77, 1.03) | 0.15 | 1.00 |
| g/1000kcal | 0.98 (0.97, 1.00) | 0.04 |  | 0.96 (0.86, 1.06) | 0.92 (0.83, 1.02) | 0.93 (0.83, 1.03) | 0.89 (0.80, 0.99) | 0.04 | 0.85 |
| Percent of total amino acids | 0.99 (0.98, 1.00) | 0.04 |  | 1.01 (0.91, 1.11) | 0.95 (0.86, 1.06) | 1.09 (0.98, 1.21) | 1.02 (0.90, 1.16) | 0.37 | 0.28 |
| Tryptophan |  |  |  |  |  |  |  |  |  |
| Percent of total protein | 0.97 (0.95, 1.00) | 0.03 |  | 0.87 (0.79, 0.96) | 0.97 (0.88, 1.08) | 0.96 (0.86, 1.07) | 0.95 (0.85, 1.07) | 0.79 | 0.28 |
| g/day | 0.94 (0.89, 0.99) | 0.03 |  | 0.91 (0.82, 1.01) | 0.88 (0.79, 0.98) | 0.93 (0.82, 1.04) | 0.87 (0.75, 1.00) | 0.14 | 0.58 |
| g/1000kcal | 0.98 (0.97, 1.00) | 0.03 |  | 0.90 (0.82, 1.00) | 0.90 (0.82, 1.00) | 0.95 (0.86, 1.05) | 0.90 (0.81, 1.00) | 0.21 | 0.71 |
| Percent of total amino acids | 0.98 (0.97, 1.00) | 0.03 |  | 1.03 (0.94, 1.12) | 1.05 (0.96, 1.15) | 1.01 (0.91, 1.12) | 0.86 (0.76, 0.99) | 0.21 | 0.18 |
| Non-essential amino acids |  |  |  |  |  |  |  |  |  |
| Percent of total protein | 0.97 (0.95, 1.00) | 0.03 |  | 0.94 (0.84, 1.04) | 1.08 (0.96, 1.20) | 1.01 (0.90, 1.14) | 0.92 (0.81, 1.05) | 0.68 | 0.04 |
| g/day | 0.94 (0.89, 0.99) | 0.03 |  | 0.91 (0.82, 1.01) | 0.89 (0.79, 0.99) | 0.87 (0.77, 0.98) | 0.84 (0.73, 0.98) | 0.04 | 0.69 |
| g/1000kcal | 0.98 (0.97, 1.00) | 0.03 |  | 0.89 (0.81, 0.99) | 0.89 (0.80, 0.98) | 0.93 (0.84, 1.03) | 0.86 (0.77, 0.96) | 0.04 | 0.22 |
| Percent of total amino acids | 0.99 (0.99, 1.00) | 0.03 |  | 0.99 (0.90, 1.09) | 1.02 (0.92, 1.12) | 1.03 (0.93, 1.14) | 1.03 (0.91, 1.15) | 0.51 | 1.00 |
| Alanine |  |  |  |  |  |  |  |  |  |
| Percent of total protein | 0.99 (0.97, 1.02) | 0.52 |  | 1.12 (1.01, 1.25) | 1.06 (0.95, 1.18) | 1.11 (0.99, 1.24) | 1.18 (1.04, 1.33) | 0.02 | 0.03 |
| g/day | 0.98 (0.94, 1.03) | 0.53 |  | 0.92 (0.83, 1.02) | 0.98 (0.87, 1.09) | 0.95 (0.84, 1.07) | 0.94 (0.81, 1.09) | 0.62 | 0.45 |
| g/1000kcal | 0.99 (0.98, 1.01) | 0.52 |  | 1.05 (0.94, 1.16) | 1.00 (0.90, 1.12) | 1.00 (0.90, 1.12) | 1.02 (0.91, 1.14) | 0.95 | 0.85 |
| Percent of total amino acids | 0.99 (0.98, 1.01) | 0.52 |  | 0.97 (0.88, 1.08) | 0.99 (0.89, 1.10) | 1.06 (0.95, 1.18) | 1.22 (1.09, 1.36) | <0.001 | <0.001 |
| Arginine |  |  |  |  |  |  |  |  |  |
| Percent of total protein | 0.99 (0.97, 1.02) | 0.57 |  | 1.01 (0.92, 1.12) | 0.99 (0.89, 1.09) | 1.06 (0.95, 1.17) | 1.11 (1.00, 1.23) | 0.04 | 0.09 |
| g/day | 0.99 (0.94, 1.04) | 0.58 |  | 0.93 (0.84, 1.03) | 0.95 (0.85, 1.06) | 0.94 (0.83, 1.07) | 0.97 (0.84, 1.12) | 0.95 | 0.50 |
| g/1000kcal | 1.00 (0.98, 1.01) | 0.57 |  | 1.03 (0.93, 1.14) | 1.02 (0.92, 1.13) | 0.98 (0.88, 1.09) | 1.05 (0.94, 1.18) | 0.53 | 0.51 |
| Percent of total amino acids | 0.99 (0.97, 1.02) | 0.57 |  | 1.05 (0.95, 1.15) | 1.09 (0.99, 1.21) | 1.10 (0.99, 1.22) | 1.22 (1.09, 1.36) | <0.001 | 0.005 |
| Aspartic acid |  |  |  |  |  |  |  |  |  |
| Percent of total protein | 0.98 (0.96, 1.01) | 0.13 |  | 1.01 (0.91, 1.12) | 0.96 (0.86, 1.07) | 1.01 (0.91, 1.13) | 1.03 (0.92, 1.15) | 0.56 | 0.98 |
| g/day | 0.96 (0.92, 1.01) | 0.15 |  | 0.99 (0.90, 1.10) | 0.93 (0.83, 1.04) | 0.97 (0.86, 1.09) | 0.97 (0.84, 1.13) | 0.80 | 0.97 |
| g/1000kcal | 0.99 (0.97, 1.00) | 0.13 |  | 0.99 (0.89, 1.10) | 0.95 (0.85, 1.05) | 0.92 (0.83, 1.03) | 0.95 (0.85, 1.06) | 0.25 | 0.88 |
| Percent of total amino acids | 0.98 (0.96, 1.01) | 0.13 |  | 1.04 (0.94, 1.15) | 1.03 (0.93, 1.14) | 1.10 (0.98, 1.23) | 1.18 (1.04, 1.34) | 0.01 | 0.09 |
| Cystine |  |  |  |  |  |  |  |  |  |
| Percent of total protein | 0.99 (0.96, 1.03) | 0.64 |  | 1.09 (0.98, 1.21) | 1.13 (1.01, 1.26) | 1.13 (1.01, 1.27) | 1.07 (0.95, 1.22) | 0.33 | 0.10 |
| g/day | 0.99 (0.94, 1.04) | 0.63 |  | 0.91 (0.82, 1.01) | 0.92 (0.82, 1.03) | 0.95 (0.84, 1.07) | 0.91 (0.78, 1.05) | 0.42 | 0.31 |
| g/1000kcal | 1.00 (0.98, 1.01) | 0.65 |  | 1.02 (0.92, 1.14) | 0.99 (0.89, 1.11) | 0.97 (0.87, 1.09) | 1.01 (0.90, 1.13) | 0.84 | 0.82 |
| Percent of total amino acids | 0.99 (0.97, 1.02) | 0.65 |  | 1.04 (0.95, 1.15) | 1.04 (0.94, 1.15) | 1.04 (0.94, 1.16) | 1.16 (1.04, 1.30) | 0.01 | 0.05 |
| Glutamic acid |  |  |  |  |  |  |  |  |  |
| Percent of total protein | 0.96 (0.93, 0.99) | 0.007* |  | 1.03 (0.93, 1.14) | 1.07 (0.96, 1.20) | 0.98 (0.87, 1.11) | 0.92 (0.80, 1.05) | 0.19 | 0.73 |
| g/day | 0.93 (0.88, 0.98) | 0.007 |  | 0.94 (0.85, 1.03) | 0.89 (0.80, 0.99) | 0.89 (0.79, 1.01) | 0.82 (0.71, 0.95) | 0.01 | 0.98 |
| g/1000kcal | 0.98 (0.96, 0.99) | 0.007 |  | 0.94 (0.85, 1.04) | 0.89 (0.80, 0.98) | 0.92 (0.83, 1.02) | 0.88 (0.79, 0.98) | 0.03 | 1.00 |
| Percent of total amino acids | 0.98 (0.96, 0.99) | 0.007 |  | 0.92 (0.83, 1.01) | 0.87 (0.78, 0.96) | 0.89 (0.80, 1.00) | 0.88 (0.78, 1.00) | 0.04 | 0.80 |
| Glycine |  |  |  |  |  |  |  |  |  |
| Percent of total protein | 1.01 (0.98, 1.03) | 0.70 |  | 1.10 (0.99, 1.23) | 1.13 (1.01, 1.27) | 1.12 (1.00, 1.26) | 1.28 (1.14, 1.43) | <0.001 | <0.001 |
| g/day | 1.01 (0.96, 1.06) | 0.71 |  | 0.92 (0.83, 1.02) | 0.97 (0.86, 1.08) | 1.00 (0.88, 1.13) | 0.99 (0.85, 1.14) | 0.67 | 0.32 |
| g/1000kcal | 1.00 (0.99, 1.02) | 0.69 |  | 1.08 (0.97, 1.20) | 1.04 (0.94, 1.16) | 1.03 (0.92, 1.15) | 1.08 (0.96, 1.21) | 0.38 | 0.40 |
| Percent of total amino acids | 1.00 (0.98, 1.03) | 0.70 |  | 0.98 (0.88, 1.09) | 1.03 (0.93, 1.14) | 1.14 (1.02, 1.26) | 1.23 (1.11, 1.37) | <0.001 | <0.001 |
| Hydroxyproline |  |  |  |  |  |  |  |  |  |
| Percent of total protein | 1.02 (0.99, 1.06) | 0.24 |  | 1.05 (0.94, 1.17) | 1.03 (0.92, 1.15) | 1.08 (0.96, 1.21) | 1.15 (1.02, 1.29) | 0.01 | 0.14 |
| g/day | 1.02 (0.98, 1.06) | 0.29 |  | 0.94 (0.84, 1.05) | 1.03 (0.92, 1.15) | 1.08 (0.96, 1.21) | 1.04 (0.93, 1.18) | 0.18 | 0.09 |
| g/1000kcal | 1.01 (0.99, 1.03) | 0.23 |  | 1.00 (0.90, 1.12) | 1.06 (0.95, 1.18) | 1.10 (0.98, 1.23) | 1.07 (0.95, 1.20) | 0.17 | 0.45 |
| Percent of total amino acids | 1.03 (0.98, 1.07) | 0.23 |  | 1.04 (0.94, 1.16) | 1.05 (0.94, 1.18) | 1.03 (0.92, 1.16) | 1.21 (1.07, 1.35) | <0.001 | 0.003 |
| Proline |  |  |  |  |  |  |  |  |  |
| Percent of total protein | 0.94 (0.91, 0.97) | <0.001* |  | 0.95 (0.85, 1.05) | 0.90 (0.80, 1.00) | 0.86 (0.77, 0.97) | 0.83 (0.73, 0.94) | 0.001 | 1.00 |
| g/day | 0.91 (0.86, 0.95) | <0.001 |  | 0.93 (0.84, 1.03) | 0.84 (0.75, 0.93) | 0.84 (0.75, 0.94) | 0.77 (0.67, 0.88) | <0.001 | 0.58 |
| g/1000kcal | 0.97 (0.96, 0.99) | <0.001 |  | 0.93 (0.84, 1.02) | 0.87 (0.79, 0.97) | 0.88 (0.79, 0.98) | 0.82 (0.74, 0.92) | <0.001 | 1.00 |
| Percent of total amino acids | 0.95 (0.93, 0.98) | <0.001 |  | 0.88 (0.80, 0.96) | 0.82 (0.75, 0.91) | 0.82 (0.73, 0.91) | 0.79 (0.70, 0.88) | <0.001 | 0.07 |
| Serine |  |  |  |  |  |  |  |  |  |
| Percent of total protein | 0.97 (0.95, 0.99) | 0.009* |  | 0.95 (0.85, 1.05) | 0.93 (0.84, 1.04) | 0.95 (0.85, 1.06) | 0.85 (0.76, 0.96) | 0.02 | 0.66 |
| g/day | 0.93 (0.89, 0.98) | 0.008 |  | 0.97 (0.88, 1.08) | 0.93 (0.84, 1.04) | 0.92 (0.81, 1.03) | 0.93 (0.81, 1.08) | 0.35 | 1.00 |
| g/1000kcal | 0.98 (0.96, 0.99) | 0.01 |  | 0.93 (0.84, 1.03) | 0.89 (0.81, 0.99) | 0.92 (0.83, 1.02) | 0.85 (0.76, 0.94) | 0.01 | 0.39 |
| Percent of total amino acids | 0.99 (0.98, 1.00) | 0.009 |  | 0.93 (0.85, 1.03) | 0.96 (0.87, 1.06) | 0.90 (0.81, 0.99) | 0.90 (0.81, 1.00) | 0.04 | 1.00 |
| Tyrosine |  |  |  |  |  |  |  |  |  |
| Percent of total protein | 0.97 (0.94, 0.99) | 0.006* |  | 0.94 (0.84, 1.04) | 0.91 (0.82, 1.01) | 0.85 (0.76, 0.95) | 0.86 (0.77, 0.96) | 0.004 | 0.60 |
| g/day | 0.93 (0.89, 0.98) | 0.006 |  | 0.95 (0.86, 1.05) | 0.91 (0.82, 1.01) | 0.91 (0.81, 1.02) | 0.85 (0.74, 0.97) | 0.02 | 1.00 |
| g/1000kcal | 0.98 (0.96, 0.99) | 0.007 |  | 0.97 (0.88, 1.08) | 0.89 (0.80, 0.99) | 0.90 (0.81, 1.00) | 0.88 (0.79, 0.98) | 0.01 | 0.75 |
| Percent of total amino acids | 0.98 (0.97, 0.99) | 0.006 |  | 0.87 (0.79, 0.96) | 0.90 (0.81, 1.00) | 0.83 (0.75, 0.92) | 0.84 (0.76, 0.93) | 0.002 | 0.06 |
| Total protein |  |  |  |  |  |  |  |  |  |
| Per SD of g/day | 0.95 (0.89, 1.00) | 0.07 |  | 0.94 (0.85, 1.05) | 0.91 (0.81, 1.02) | 0.88 (0.78, 1.01) | 0.95 (0.81, 1.12) | 0.75 | 0.39 |
| Per SD of g/1000kcal | 0.99 (0.97, 1.00) | 0.07 |  | 0.98 (0.88, 1.08) | 0.96 (0.87, 1.06) | 0.95 (0.86, 1.06) | 0.91 (0.82, 1.02) | 0.09 | 1.00 |

1. Based on multivariable model stratified by sex and centre, and adjusted for age (continuous), observed energy intake (continuous), smoking (never, former, current<10, 10-19, 20+ cigarettes/day, unknown), observed alcohol consumption (non-drinkers (<0.1), 0.1-4.9, 5.0-14.9, 15-29.9, 30-59.9, 60+ g/day), physical activity (inactive, moderately inactive, moderately active, active, unknown), employment status (employed or student, neither employed nor student, unknown), highest level of education completed (none or primary, secondary, vocational or university, unknown), history of diabetes (yes, no, unknown), prior hypertension (yes, no , unknown), prior hyperlipidaemia (yes, no, unknown), body mass index (<22.5, 22.5-24.9, 25.0-27.4, 27.5-29.9, ≥30.0 kg/m^2^, unknown).
2. Linear trends estimated by including the exposures as continuous variables in the Cox regression. Estimates with an asterisk denotes ones which were statistically significant after correcting for multiple testing.
3. Linear trends estimated by replacing the values with the median value in each fifth of intake and fitting as a pseudo-continuous variable in the Cox regression.
4. P-value from test of heterogeneity comparing models with the exposure expressed as a true continuous variable versus a categorical variable.

Supplementary table 5: Hazard ratios (95% confidence intervals)^1^ for **haemorrhagic stroke** (1375 cases) by increments and fifths of **observed intakes** of dietary amino acids.

| Amino acids | Per SD of observed intakes | |  | Fifths of observed intakes compared to bottom fifth, HR(95% CI) | | | |  |  |
| --- | --- | --- | --- | --- | --- | --- | --- | --- | --- |
|  | HR (95% CI) | p-trend^2^ |  | 2 | 3 | 4 | 5 | p-trend^3^ | p for non-linearity^4^ |
| Branched-chain amino acids |  |  |  |  |  |  |  |  |  |
| Percent of total protein | 0.98 (0.95, 1.02) | 0.38 |  | 0.88 (0.74, 1.05) | 0.89 (0.74, 1.07) | 0.89 (0.74, 1.08) | 0.86 (0.71, 1.06) | 0.19 | 0.59 |
| g/day | 0.97 (0.89, 1.06) | 0.48 |  | 1.01 (0.84, 1.21) | 0.99 (0.81, 1.19) | 0.82 (0.66, 1.01) | 0.98 (0.76, 1.25) | 0.58 | 0.07 |
| g/1000kcal | 0.99 (0.96, 1.01) | 0.34 |  | 0.89 (0.75, 1.07) | 0.86 (0.72, 1.03) | 0.77 (0.64, 0.92) | 0.93 (0.78, 1.12) | 0.37 | 0.03 |
| Percent of total amino acids | 1.00 (0.98, 1.01) | 0.40 |  | 0.87 (0.73, 1.04) | 1.03 (0.86, 1.22) | 0.97 (0.81, 1.15) | 0.87 (0.72, 1.04) | 0.36 | 0.11 |
| Isoleucine |  |  |  |  |  |  |  |  |  |
| Percent of total protein | 0.98 (0.95, 1.02) | 0.38 |  | 0.92 (0.77, 1.10) | 0.80 (0.67, 0.97) | 0.98 (0.81, 1.18) | 0.91 (0.74, 1.11) | 0.43 | 0.09 |
| g/day | 0.97 (0.89, 1.06) | 0.50 |  | 1.01 (0.85, 1.21) | 0.93 (0.76, 1.12) | 0.81 (0.66, 1.01) | 0.92 (0.72, 1.18) | 0.33 | 0.10 |
| g/1000kcal | 0.99 (0.96, 1.01) | 0.35 |  | 0.92 (0.77, 1.10) | 0.85 (0.71, 1.02) | 0.76 (0.63, 0.91) | 0.94 (0.78, 1.13) | 0.33 | 0.02 |
| Percent of total amino acids | 0.99 (0.98, 1.01) | 0.48 |  | 1.01 (0.84, 1.21) | 1.03 (0.86, 1.24) | 1.08 (0.89, 1.30) | 1.00 (0.80, 1.25) | 0.78 | 0.93 |
| Leucine |  |  |  |  |  |  |  |  |  |
| Percent of total protein | 0.98 (0.95, 1.02) | 0.41 |  | 0.82 (0.68, 0.98) | 0.88 (0.73, 1.06) | 0.86 (0.71, 1.05) | 0.84 (0.68, 1.03) | 0.14 | 0.21 |
| g/day | 0.97 (0.89, 1.06) | 0.51 |  | 0.98 (0.82, 1.18) | 1.00 (0.83, 1.21) | 0.80 (0.65, 1.00) | 0.98 (0.77, 1.25) | 0.65 | 0.04 |
| g/1000kcal | 0.99 (0.96, 1.02) | 0.38 |  | 0.88 (0.74, 1.05) | 0.86 (0.72, 1.02) | 0.77 (0.64, 0.93) | 0.93 (0.77, 1.12) | 0.39 | 0.04 |
| Percent of total amino acids | 1.00 (0.99, 1.01) | 0.42 |  | 0.94 (0.79, 1.12) | 0.96 (0.81, 1.15) | 1.02 (0.86, 1.22) | 0.98 (0.82, 1.17) | 0.91 | 0.94 |
| Valine |  |  |  |  |  |  |  |  |  |
| Percent of total protein | 0.98 (0.94, 1.02) | 0.33 |  | 0.83 (0.69, 0.99) | 0.89 (0.74, 1.06) | 0.82 (0.68, 0.99) | 0.84 (0.69, 1.02) | 0.08 | 0.19 |
| g/day | 0.97 (0.89, 1.05) | 0.41 |  | 0.97 (0.81, 1.16) | 0.97 (0.80, 1.17) | 0.81 (0.66, 1.00) | 0.91 (0.71, 1.16) | 0.29 | 0.17 |
| g/1000kcal | 0.99 (0.96, 1.01) | 0.29 |  | 0.85 (0.72, 1.02) | 0.85 (0.72, 1.02) | 0.75 (0.62, 0.90) | 0.91 (0.76, 1.09) | 0.31 | 0.02 |
| Percent of total amino acids | 0.99 (0.98, 1.01) | 0.33 |  | 1.02 (0.86, 1.21) | 1.03 (0.87, 1.23) | 1.05 (0.88, 1.25) | 0.88 (0.73, 1.06) | 0.25 | 0.24 |
| Other essential amino acids |  |  |  |  |  |  |  |  |  |
| Percent of total protein | 0.99 (0.95, 1.03) | 0.52 |  | 0.92 (0.76, 1.11) | 0.90 (0.75, 1.09) | 1.02 (0.84, 1.23) | 0.86 (0.70, 1.05) | 0.32 | 0.15 |
| g/day | 0.98 (0.90, 1.07) | 0.66 |  | 1.03 (0.86, 1.24) | 1.02 (0.84, 1.24) | 0.86 (0.70, 1.07) | 1.04 (0.81, 1.32) | 0.94 | 0.10 |
| g/1000kcal | 0.99 (0.96, 1.02) | 0.49 |  | 0.98 (0.82, 1.17) | 0.88 (0.74, 1.06) | 0.75 (0.62, 0.90) | 1.02 (0.85, 1.23) | 0.71 | 0.001 |
| Percent of total amino acids | 1.00 (0.98, 1.01) | 0.57 |  | 1.07 (0.88, 1.30) | 1.13 (0.93, 1.37) | 1.07 (0.88, 1.31) | 1.14 (0.92, 1.41) | 0.27 | 0.64 |
| Histidine |  |  |  |  |  |  |  |  |  |
| Percent of total protein | 0.98 (0.95, 1.02) | 0.43 |  | 0.93 (0.77, 1.12) | 0.87 (0.72, 1.05) | 0.95 (0.78, 1.15) | 0.90 (0.73, 1.10) | 0.37 | 0.62 |
| g/day | 0.98 (0.90, 1.06) | 0.58 |  | 1.03 (0.86, 1.23) | 0.99 (0.81, 1.20) | 0.86 (0.70, 1.07) | 0.99 (0.78, 1.27) | 0.73 | 0.21 |
| g/1000kcal | 0.99 (0.96, 1.02) | 0.40 |  | 1.04 (0.87, 1.24) | 0.92 (0.77, 1.11) | 0.83 (0.68, 1.00) | 1.01 (0.83, 1.22) | 0.59 | 0.04 |
| Percent of total amino acids | 0.99 (0.97, 1.01) | 0.43 |  | 0.94 (0.79, 1.12) | 0.89 (0.74, 1.06) | 0.95 (0.79, 1.14) | 0.98 (0.81, 1.19) | 0.97 | 0.65 |
| Lysine |  |  |  |  |  |  |  |  |  |
| Percent of total protein | 0.99 (0.95, 1.03) | 0.68 |  | 0.93 (0.76, 1.13) | 0.88 (0.73, 1.08) | 0.96 (0.79, 1.17) | 0.90 (0.73, 1.10) | 0.44 | 0.57 |
| g/day | 0.99 (0.91, 1.08) | 0.83 |  | 1.15 (0.96, 1.38) | 0.98 (0.80, 1.20) | 0.96 (0.77, 1.18) | 1.06 (0.83, 1.35) | 0.92 | 0.11 |
| g/1000kcal | 0.99 (0.97, 1.02) | 0.66 |  | 1.00 (0.83, 1.20) | 0.86 (0.72, 1.04) | 0.77 (0.63, 0.93) | 1.04 (0.86, 1.26) | 0.91 | 0.001 |
| Percent of total amino acids | 0.99 (0.96, 1.03) | 0.72 |  | 1.12 (0.92, 1.37) | 1.04 (0.85, 1.27) | 1.15 (0.93, 1.41) | 1.14 (0.92, 1.42) | 0.25 | 0.40 |
| Methionine |  |  |  |  |  |  |  |  |  |
| Percent of total protein | 0.99 (0.95, 1.03) | 0.58 |  | 0.80 (0.66, 0.97) | 0.85 (0.70, 1.02) | 0.91 (0.75, 1.10) | 0.85 (0.69, 1.05) | 0.29 | 0.12 |
| g/day | 0.98 (0.91, 1.07) | 0.72 |  | 1.05 (0.87, 1.26) | 0.91 (0.75, 1.11) | 0.88 (0.71, 1.09) | 0.96 (0.75, 1.22) | 0.54 | 0.23 |
| g/1000kcal | 0.99 (0.96, 1.02) | 0.56 |  | 0.93 (0.78, 1.12) | 0.90 (0.75, 1.08) | 0.75 (0.62, 0.91) | 1.00 (0.83, 1.20) | 0.73 | 0.003 |
| Percent of total amino acids | 0.99 (0.97, 1.02) | 0.58 |  | 1.04 (0.86, 1.27) | 1.09 (0.89, 1.34) | 1.18 (0.96, 1.44) | 1.06 (0.86, 1.30) | 0.40 | 0.36 |
| Phenylalanine |  |  |  |  |  |  |  |  |  |
| Percent of total protein | 0.98 (0.94, 1.02) | 0.41 |  | 0.77 (0.64, 0.91) | 0.94 (0.79, 1.12) | 0.84 (0.69, 1.02) | 0.84 (0.67, 1.03) | 0.16 | 0.02 |
| g/day | 0.97 (0.89, 1.06) | 0.51 |  | 1.00 (0.83, 1.19) | 0.97 (0.81, 1.18) | 0.79 (0.64, 0.98) | 0.94 (0.73, 1.21) | 0.42 | 0.04 |
| g/1000kcal | 0.99 (0.96, 1.01) | 0.36 |  | 0.84 (0.71, 1.00) | 0.93 (0.78, 1.10) | 0.76 (0.63, 0.91) | 0.95 (0.79, 1.14) | 0.52 | 0.01 |
| Percent of total amino acids | 1.00 (0.98, 1.01) | 0.41 |  | 0.87 (0.75, 1.02) | 1.01 (0.86, 1.19) | 0.94 (0.79, 1.13) | 0.86 (0.70, 1.05) | 0.30 | 0.19 |
| Threonine |  |  |  |  |  |  |  |  |  |
| Percent of total protein | 0.99 (0.95, 1.02) | 0.47 |  | 0.95 (0.79, 1.14) | 0.83 (0.69, 1.00) | 0.95 (0.79, 1.15) | 0.90 (0.74, 1.10) | 0.35 | 0.26 |
| g/day | 0.98 (0.90, 1.07) | 0.62 |  | 1.03 (0.86, 1.24) | 0.97 (0.80, 1.18) | 0.85 (0.68, 1.05) | 1.00 (0.78, 1.28) | 0.74 | 0.12 |
| g/1000kcal | 0.99 (0.96, 1.02) | 0.45 |  | 0.98 (0.83, 1.17) | 0.81 (0.68, 0.98) | 0.73 (0.61, 0.88) | 0.98 (0.82, 1.18) | 0.41 | <0.001 |
| Percent of total amino acids | 0.99 (0.98, 1.01) | 0.54 |  | 0.90 (0.75, 1.08) | 0.96 (0.80, 1.15) | 1.04 (0.86, 1.25) | 0.91 (0.73, 1.14) | 0.86 | 0.31 |
| Tryptophan |  |  |  |  |  |  |  |  |  |
| Percent of total protein | 0.98 (0.94, 1.02) | 0.41 |  | 0.98 (0.82, 1.17) | 0.99 (0.82, 1.19) | 1.03 (0.85, 1.25) | 0.96 (0.78, 1.18) | 0.89 | 1.00 |
| g/day | 0.97 (0.89, 1.06) | 0.52 |  | 0.89 (0.75, 1.07) | 0.92 (0.76, 1.11) | 0.80 (0.65, 0.98) | 0.90 (0.70, 1.16) | 0.45 | 0.15 |
| g/1000kcal | 0.99 (0.96, 1.02) | 0.38 |  | 0.81 (0.68, 0.97) | 0.85 (0.71, 1.01) | 0.82 (0.69, 0.98) | 0.88 (0.73, 1.06) | 0.28 | 0.10 |
| Percent of total amino acids | 0.99 (0.96, 1.01) | 0.36 |  | 1.12 (0.96, 1.31) | 1.13 (0.95, 1.34) | 1.06 (0.87, 1.30) | 1.02 (0.82, 1.28) | 0.28 | 0.51 |
| Non-essential amino acids |  |  |  |  |  |  |  |  |  |
| Percent of total protein | 0.99 (0.95, 1.03) | 0.48 |  | 0.79 (0.66, 0.94) | 0.83 (0.69, 1.01) | 0.87 (0.71, 1.06) | 0.84 (0.67, 1.05) | 0.13 | 0.07 |
| g/day | 0.98 (0.89, 1.07) | 0.59 |  | 0.96 (0.80, 1.15) | 0.97 (0.80, 1.18) | 0.82 (0.66, 1.02) | 0.98 (0.76, 1.26) | 0.80 | 0.13 |
| g/1000kcal | 0.99 (0.96, 1.02) | 0.43 |  | 0.90 (0.75, 1.07) | 0.93 (0.78, 1.11) | 0.80 (0.66, 0.96) | 0.96 (0.80, 1.16) | 0.54 | 0.08 |
| Percent of total amino acids | 1.00 (0.99, 1.01) | 0.50 |  | 1.18 (1.00, 1.39) | 1.01 (0.85, 1.21) | 1.00 (0.82, 1.21) | 1.12 (0.91, 1.37) | 0.79 | 0.09 |
| Alanine |  |  |  |  |  |  |  |  |  |
| Percent of total protein | 0.98 (0.94, 1.03) | 0.48 |  | 0.97 (0.81, 1.17) | 0.88 (0.73, 1.07) | 0.98 (0.80, 1.18) | 0.97 (0.78, 1.19) | 0.73 | 0.65 |
| g/day | 0.98 (0.90, 1.07) | 0.63 |  | 0.95 (0.79, 1.14) | 0.88 (0.72, 1.07) | 0.87 (0.70, 1.07) | 0.89 (0.69, 1.13) | 0.38 | 0.55 |
| g/1000kcal | 0.99 (0.96, 1.02) | 0.45 |  | 1.00 (0.84, 1.19) | 0.85 (0.71, 1.02) | 0.71 (0.58, 0.86) | 0.99 (0.82, 1.20) | 0.43 | <0.001 |
| Percent of total amino acids | 0.99 (0.96, 1.02) | 0.53 |  | 1.01 (0.84, 1.21) | 1.08 (0.90, 1.29) | 0.99 (0.82, 1.20) | 0.98 (0.81, 1.19) | 0.78 | 0.80 |
| Arginine |  |  |  |  |  |  |  |  |  |
| Percent of total protein | 0.99 (0.95, 1.03) | 0.50 |  | 0.95 (0.80, 1.13) | 0.87 (0.73, 1.04) | 0.99 (0.83, 1.18) | 0.89 (0.74, 1.07) | 0.33 | 0.34 |
| g/day | 0.98 (0.90, 1.07) | 0.66 |  | 0.88 (0.73, 1.05) | 0.84 (0.69, 1.02) | 0.83 (0.67, 1.02) | 0.78 (0.61, 0.99) | 0.07 | 0.24 |
| g/1000kcal | 0.99 (0.96, 1.02) | 0.49 |  | 0.97 (0.81, 1.15) | 0.89 (0.75, 1.07) | 0.76 (0.63, 0.92) | 1.01 (0.84, 1.22) | 0.71 | 0.003 |
| Percent of total amino acids | 0.99 (0.95, 1.03) | 0.53 |  | 1.06 (0.89, 1.25) | 1.07 (0.90, 1.27) | 0.96 (0.80, 1.15) | 1.02 (0.84, 1.23) | 0.82 | 0.65 |
| Aspartic acid |  |  |  |  |  |  |  |  |  |
| Percent of total protein | 1.00 (0.96, 1.04) | 0.97 |  | 1.00 (0.84, 1.20) | 0.82 (0.68, 0.99) | 0.99 (0.82, 1.19) | 1.00 (0.82, 1.21) | 0.99 | 0.05 |
| g/day | 1.01 (0.93, 1.10) | 0.78 |  | 0.93 (0.77, 1.12) | 0.95 (0.78, 1.15) | 0.87 (0.70, 1.07) | 1.00 (0.78, 1.29) | 0.86 | 0.27 |
| g/1000kcal | 1.00 (0.97, 1.03) | 1.00 |  | 0.92 (0.77, 1.11) | 0.88 (0.74, 1.06) | 0.80 (0.66, 0.96) | 0.98 (0.82, 1.19) | 0.76 | 0.03 |
| Percent of total amino acids | 1.00 (0.96, 1.04) | 0.94 |  | 1.10 (0.92, 1.33) | 1.12 (0.93, 1.36) | 1.15 (0.94, 1.40) | 1.14 (0.91, 1.43) | 0.27 | 0.57 |
| Cystine |  |  |  |  |  |  |  |  |  |
| Percent of total protein | 0.96 (0.90, 1.02) | 0.21 |  | 0.91 (0.76, 1.08) | 0.91 (0.76, 1.10) | 0.89 (0.72, 1.08) | 0.79 (0.63, 0.99) | 0.06 | 0.43 |
| g/day | 0.96 (0.87, 1.04) | 0.31 |  | 0.83 (0.69, 0.99) | 0.85 (0.71, 1.03) | 0.77 (0.62, 0.95) | 0.82 (0.64, 1.06) | 0.25 | 0.13 |
| g/1000kcal | 0.98 (0.95, 1.01) | 0.19 |  | 0.94 (0.79, 1.11) | 0.80 (0.67, 0.97) | 0.78 (0.64, 0.95) | 0.91 (0.75, 1.11) | 0.22 | 0.04 |
| Percent of total amino acids | 0.97 (0.92, 1.02) | 0.20 |  | 1.04 (0.88, 1.23) | 1.00 (0.84, 1.18) | 0.93 (0.77, 1.11) | 0.87 (0.71, 1.06) | 0.10 | 0.53 |
| Glutamic acid |  |  |  |  |  |  |  |  |  |
| Percent of total protein | 0.98 (0.93, 1.03) | 0.48 |  | 0.82 (0.69, 0.97) | 0.90 (0.74, 1.08) | 0.86 (0.69, 1.06) | 0.81 (0.65, 1.02) | 0.12 | 0.13 |
| g/day | 0.97 (0.89, 1.07) | 0.55 |  | 0.92 (0.77, 1.10) | 1.01 (0.83, 1.22) | 0.89 (0.71, 1.10) | 0.99 (0.76, 1.27) | 0.98 | 0.36 |
| g/1000kcal | 0.99 (0.96, 1.02) | 0.41 |  | 0.95 (0.80, 1.13) | 0.98 (0.82, 1.17) | 0.82 (0.68, 0.99) | 0.99 (0.82, 1.20) | 0.61 | 0.10 |
| Percent of total amino acids | 0.99 (0.96, 1.02) | 0.50 |  | 0.98 (0.83, 1.16) | 1.07 (0.90, 1.27) | 0.96 (0.79, 1.16) | 0.98 (0.79, 1.21) | 0.82 | 0.67 |
| Glycine |  |  |  |  |  |  |  |  |  |
| Percent of total protein | 0.98 (0.93, 1.03) | 0.42 |  | 1.06 (0.89, 1.27) | 0.98 (0.82, 1.19) | 0.95 (0.78, 1.16) | 0.97 (0.79, 1.18) | 0.50 | 0.80 |
| g/day | 0.98 (0.90, 1.06) | 0.58 |  | 0.83 (0.69, 1.00) | 0.92 (0.76, 1.12) | 0.85 (0.69, 1.05) | 0.88 (0.69, 1.13) | 0.60 | 0.24 |
| g/1000kcal | 0.99 (0.96, 1.02) | 0.38 |  | 1.03 (0.86, 1.23) | 0.86 (0.71, 1.04) | 0.87 (0.72, 1.05) | 0.94 (0.77, 1.14) | 0.30 | 0.14 |
| Percent of total amino acids | 0.98 (0.94, 1.03) | 0.45 |  | 1.07 (0.90, 1.28) | 1.04 (0.87, 1.24) | 0.97 (0.81, 1.16) | 0.97 (0.81, 1.17) | 0.49 | 0.69 |
| Hydroxyproline |  |  |  |  |  |  |  |  |  |
| Percent of total protein | 1.06 (1.00, 1.14) | 0.06 |  | 1.07 (0.88, 1.31) | 1.21 (0.99, 1.48) | 1.16 (0.94, 1.43) | 1.29 (1.04, 1.60) | 0.02 | 0.29 |
| g/day | 1.07 (1.01, 1.13) | 0.03 |  | 1.01 (0.83, 1.23) | 1.18 (0.97, 1.44) | 1.07 (0.87, 1.31) | 1.21 (0.98, 1.51) | 0.08 | 0.57 |
| g/1000kcal | 1.03 (1.00, 1.05) | 0.07 |  | 1.13 (0.93, 1.38) | 1.20 (0.98, 1.47) | 1.21 (0.98, 1.49) | 1.22 (0.99, 1.51) | 0.12 | 0.78 |
| Percent of total amino acids | 1.08 (1.00, 1.16) | 0.07 |  | 1.13 (0.93, 1.38) | 1.27 (1.04, 1.56) | 1.21 (0.98, 1.49) | 1.30 (1.05, 1.62) | 0.03 | 0.23 |
| Proline |  |  |  |  |  |  |  |  |  |
| Percent of total protein | 0.96 (0.91, 1.02) | 0.17 |  | 0.82 (0.69, 0.97) | 0.79 (0.66, 0.95) | 0.79 (0.64, 0.96) | 0.72 (0.58, 0.90) | 0.006 | 0.05 |
| g/day | 0.94 (0.86, 1.03) | 0.20 |  | 0.82 (0.69, 0.98) | 0.90 (0.75, 1.08) | 0.77 (0.63, 0.94) | 0.75 (0.59, 0.96) | 0.03 | 0.07 |
| g/1000kcal | 0.98 (0.95, 1.01) | 0.15 |  | 1.00 (0.84, 1.19) | 0.99 (0.83, 1.19) | 0.88 (0.73, 1.06) | 0.93 (0.77, 1.13) | 0.27 | 0.78 |
| Percent of total amino acids | 0.97 (0.93, 1.01) | 0.17 |  | 0.89 (0.75, 1.05) | 0.91 (0.76, 1.08) | 0.94 (0.78, 1.13) | 0.74 (0.60, 0.91) | 0.02 | 0.06 |
| Serine |  |  |  |  |  |  |  |  |  |
| Percent of total protein | 0.99 (0.95, 1.03) | 0.72 |  | 0.85 (0.71, 1.01) | 0.90 (0.75, 1.08) | 0.86 (0.70, 1.04) | 0.94 (0.76, 1.15) | 0.51 | 0.23 |
| g/day | 0.99 (0.91, 1.08) | 0.83 |  | 1.00 (0.84, 1.20) | 1.00 (0.83, 1.21) | 0.84 (0.68, 1.04) | 0.98 (0.76, 1.26) | 0.64 | 0.12 |
| g/1000kcal | 0.99 (0.97, 1.02) | 0.66 |  | 0.82 (0.69, 0.98) | 0.90 (0.75, 1.07) | 0.81 (0.67, 0.97) | 0.94 (0.78, 1.12) | 0.67 | 0.05 |
| Percent of total amino acids | 1.00 (0.98, 1.01) | 0.71 |  | 1.11 (0.94, 1.32) | 1.06 (0.89, 1.27) | 1.18 (0.99, 1.40) | 1.15 (0.96, 1.38) | 0.11 | 0.26 |
| Tyrosine |  |  |  |  |  |  |  |  |  |
| Percent of total protein | 0.99 (0.95, 1.03) | 0.57 |  | 0.83 (0.69, 0.99) | 0.87 (0.72, 1.04) | 0.83 (0.69, 1.01) | 0.88 (0.72, 1.07) | 0.29 | 0.19 |
| g/day | 0.98 (0.90, 1.07) | 0.67 |  | 1.00 (0.83, 1.20) | 1.06 (0.88, 1.28) | 0.88 (0.71, 1.08) | 0.94 (0.74, 1.20) | 0.39 | 0.19 |
| g/1000kcal | 0.99 (0.96, 1.02) | 0.53 |  | 0.84 (0.70, 1.00) | 0.83 (0.69, 0.99) | 0.82 (0.68, 0.98) | 0.91 (0.76, 1.09) | 0.56 | 0.08 |
| Percent of total amino acids | 0.99 (0.97, 1.02) | 0.57 |  | 0.95 (0.80, 1.14) | 0.94 (0.78, 1.12) | 1.00 (0.84, 1.20) | 0.99 (0.82, 1.18) | 0.91 | 0.89 |
| Total protein |  |  |  |  |  |  |  |  |  |
| Per SD of g/day | 1.00 (0.90, 1.11) | 0.99 |  | 0.97 (0.81, 1.17) | 1.00 (0.82, 1.22) | 0.93 (0.74, 1.17) | 1.00 (0.76, 1.32) | 0.97 | 0.80 |
| Per SD of g/1000kcal | 0.99 (0.97, 1.02) | 0.70 |  | 0.91 (0.77, 1.09) | 0.83 (0.70, 1.00) | 0.89 (0.74, 1.06) | 0.98 (0.82, 1.18) | 0.89 | 0.13 |

1. Based on multivariable model stratified by sex and centre, and adjusted for age (continuous), observed energy intake (continuous), smoking (never, former, current<10, 10-19, 20+ cigarettes/day, unknown), observed alcohol consumption (non-drinkers (<0.1), 0.1-4.9, 5.0-14.9, 15-29.9, 30-59.9, 60+ g/day), physical activity (inactive, moderately inactive, moderately active, active, unknown), employment status (employed or student, neither employed nor student, unknown), highest level of education completed (none or primary, secondary, vocational or university, unknown), history of diabetes (yes, no, unknown), prior hypertension (yes, no , unknown), prior hyperlipidaemia (yes, no, unknown), body mass index (<22.5, 22.5-24.9, 25.0-27.4, 27.5-29.9, ≥30.0 kg/m^2^, unknown).
2. Linear trends estimated by including the exposures as continuous variables in the Cox regression.
3. Linear trends estimated by replacing the values with the median value in each fifth of intake and fitting as a pseudo-continuous variable in the Cox regression.
4. P-value from test of heterogeneity comparing models with the exposure expressed as a true continuous variable versus a categorical variable.

Supplementary table 6: Hazard ratios (95% confidence intervals)^1^ for **intracerebral** (915 cases) and **subarachnoid haemorrhagic stroke** (460 cases) by increments of calibrated intakes of dietary amino acids.

| Amino acids (percent of total protein) | Intracerebral haemorrhage | |  | Subarachnoid haemorrhage | |
| --- | --- | --- | --- | --- | --- |
|  | HR (95% CI) | p-trend^2^ |  | HR (95% CI) | p-trend^2^ |
| Branched-chain amino acids | 0.92 (0.81, 1.04) | 0.17 |  | 1.08 (0.91, 1.28) | 0.39 |
| Isoleucine | 0.92 (0.81, 1.04) | 0.17 |  | 1.08 (0.90, 1.28) | 0.41 |
| Leucine | 0.92 (0.81, 1.04) | 0.18 |  | 1.08 (0.91, 1.29) | 0.37 |
| Valine | 0.92 (0.82, 1.03) | 0.17 |  | 1.07 (0.91, 1.26) | 0.42 |
| Other essential amino acids | 0.92 (0.81, 1.05) | 0.20 |  | 1.09 (0.91, 1.31) | 0.35 |
| Histidine | 0.90 (0.79, 1.03) | 0.13 |  | 1.10 (0.91, 1.32) | 0.34 |
| Lysine | 0.92 (0.81, 1.06) | 0.26 |  | 1.11 (0.92, 1.35) | 0.29 |
| Methionine | 0.92 (0.80, 1.06) | 0.24 |  | 1.10 (0.91, 1.34) | 0.34 |
| Phenylalanine | 0.92 (0.81, 1.04) | 0.19 |  | 1.08 (0.90, 1.28) | 0.41 |
| Threonine | 0.92 (0.82, 1.04) | 0.20 |  | 1.07 (0.90, 1.27) | 0.47 |
| Tryptophan | 0.94 (0.83, 1.05) | 0.26 |  | 1.09 (0.92, 1.28) | 0.34 |
| Non-essential amino acids | 0.92 (0.80, 1.05) | 0.20 |  | 1.09 (0.91, 1.32) | 0.35 |
| Alanine | 0.91 (0.79, 1.04) | 0.17 |  | 1.03 (0.84, 1.25) | 0.79 |
| Arginine | 0.91 (0.82, 1.02) | 0.11 |  | 1.01 (0.86, 1.18) | 0.91 |
| Aspartic acid | 0.96 (0.87, 1.05) | 0.37 |  | 1.09 (0.95, 1.25) | 0.21 |
| Cystine | 0.89 (0.76, 1.05) | 0.17 |  | 0.96 (0.76, 1.22) | 0.74 |
| Glutamic acid | 0.91 (0.78, 1.06) | 0.23 |  | 1.12 (0.91, 1.39) | 0.29 |
| Glycine | 0.91 (0.79, 1.06) | 0.22 |  | 0.99 (0.81, 1.21) | 0.94 |
| Hydroxyproline | 1.09 (0.94, 1.26) | 0.24 |  | 1.22 (1.03, 1.45) | 0.02 |
| Proline | 0.90 (0.78, 1.04) | 0.15 |  | 1.08 (0.89, 1.32) | 0.42 |
| Serine | 0.95 (0.85, 1.07) | 0.39 |  | 1.09 (0.93, 1.29) | 0.29 |
| Tyrosine | 0.94 (0.83, 1.05) | 0.27 |  | 1.12 (0.95, 1.32) | 0.19 |
| Total protein (g/1000 kcal) | 0.96 (0.90, 1.02) | 0.20 |  | 1.04 (0.96, 1.14) | 0.34 |

1. Hazard ratios modelled per 1 sex-specific SD increment in dietary amino acids, expressed as percent of total protein. Based on multivariable model stratified by sex and centre, and adjusted for age (continuous), calibrated energy intake (continuous), smoking (never, former, current<10, 10-19, 20+ cigarettes/day, unknown), calibrated alcohol consumption (non-drinkers (<0.1), 0.1-4.9, 5.0-14.9, 15-29.9, 30-59.9, 60+ g/day), physical activity (inactive, moderately inactive, moderately active, active, unknown), employment status (employed or student, neither employed nor student, unknown), highest level of education completed (none or primary, secondary, vocational or university, unknown), history of diabetes (yes, no, unknown), prior hypertension (yes, no , unknown), prior hyperlipidaemia (yes, no, unknown), body mass index (<22.5, 22.5-24.9, 25.0-27.4, 27.5-29.9, ≥30.0 kg/m^2^, unknown).

Supplementary table 7: Hazard ratios (95% confidence intervals) for **ischaemic stroke** (4295 cases) by increments of **calibrated** intakes with **different levels of adjustment**

|  | HR (95% CIs) for per SD of calibrated intakes | | | | | |
| --- | --- | --- | --- | --- | --- | --- |
| Amino acids (percent of total protein) | Model 1^1^ | Model 2^2^ | Model 3^3^ | Model 4^4^ | Model 5^5^ | Model 6^6^ |
| Branched-chain amino acids | 0.93 (0.90, 0.97) | 0.96 (0.91, 1.02) | 1.01 (0.95, 1.06) | 0.95 (0.90, 1.00) | 0.93 (0.88, 0.98) | 0.95 (0.91, 0.98) |
| Likelihood-ratio test statistic^7^ | 14.3 | 1.9 | 0.04 | 3.6 | 7.7 | 8.6 |
| Isoleucine | 0.94 (0.90, 0.97) | 0.97 (0.92, 1.02) | 1.01 (0.96, 1.07) | 0.95 (0.90, 1.01) | 0.93 (0.88, 0.98) | 0.95 (0.91, 0.98) |
| Likelihood-ratio test statistic | 12.8 | 1.1 | 0.1 | 3.0 | 7.0 | 8.1 |
| Leucine | 0.93 (0.90, 0.97) | 0.97 (0.91, 1.02) | 1.01 (0.95, 1.07) | 0.95 (0.90, 1.00) | 0.93 (0.87, 0.98) | 0.95 (0.91, 0.98) |
| Likelihood-ratio test statistic | 13.8 | 1.6 | 0.1 | 3.3 | 7.3 | 8.3 |
| Valine | 0.93 (0.90, 0.96) | 0.96 (0.91, 1.01) | 1.00 (0.95, 1.05) | 0.95 (0.90, 1.00) | 0.92 (0.88, 0.97) | 0.95 (0.92, 0.98) |
| Likelihood-ratio test statistic | 16.3 | 3.1 | <0.01 | 4.5 | 8.7 | 9.3 |
| Other essential amino acids | 0.94 (0.90, 0.98) | 0.99 (0.93, 1.04) | 1.02 (0.97, 1.08) | 0.96 (0.91, 1.02) | 0.93 (0.88, 0.99) | 0.95 (0.91, 0.99) |
| Likelihood-ratio test statistic | 9.8 | 0.2 | 0.7 | 1.9 | 5.5 | 7.1 |
| Histidine | 0.94 (0.91, 0.98) | 1.00 (0.94, 1.06) | 1.04 (0.98, 1.10) | 0.97 (0.91, 1.03) | 0.94 (0.88, 1.00) | 0.95 (0.91, 0.99) |
| Likelihood-ratio test statistic | 7.6 | <0.01 | 1.5 | 1.1 | 4.2 | 6.1 |
| Lysine | 0.95 (0.90, 0.99) | 1.01 (0.95, 1.07) | 1.04 (0.98, 1.10) | 0.97 (0.91, 1.03) | 0.94 (0.88, 1.00) | 0.95 (0.90, 0.99) |
| Likelihood-ratio test statistic | 6.2 | 0.04 | 1.5 | 1.1 | 4.2 | 6.3 |
| Methionine | 0.94 (0.90, 0.98) | 0.99 (0.94, 1.06) | 1.03 (0.97, 1.10) | 0.96 (0.91, 1.03) | 0.93 (0.88, 0.99) | 0.94 (0.90, 0.99) |
| Likelihood-ratio test statistic | 8.0 | 0.0 | 1.1 | 1.3 | 4.6 | 6.5 |
| Phenylalanine | 0.93 (0.90, 0.96) | 0.95 (0.90, 1.00) | 1.00 (0.95, 1.06) | 0.95 (0.90, 1.00) | 0.92 (0.87, 0.98) | 0.95 (0.92, 0.98) |
| Likelihood-ratio test statistic | 17.1 | 3.3 | <0.01 | 4.1 | 8.0 | 8.5 |
| Threonine | 0.94 (0.91, 0.98) | 0.98 (0.93, 1.04) | 1.02 (0.97, 1.08) | 0.96 (0.91, 1.01) | 0.93 (0.88, 0.99) | 0.95 (0.92, 0.99) |
| Likelihood-ratio test statistic | 10.7 | 0.4 | 0.5 | 2.3 | 6.2 | 7.6 |
| Tryptophan | 0.94 (0.91, 0.97) | 0.97 (0.92, 1.02) | 1.01 (0.96, 1.06) | 0.96 (0.91, 1.01) | 0.94 (0.89, 0.99) | 0.96 (0.92, 0.99) |
| Likelihood-ratio test statistic | 14.8 | 1.9 | 0.1 | 3.0 | 6.2 | 7.4 |
| Non-essential amino acids | 0.93 (0.90, 0.96) | 0.96 (0.91, 1.02) | 1.01 (0.96, 1.07) | 0.95 (0.89, 1.00) | 0.92 (0.87, 0.98) | 0.95 (0.92, 0.98) |
| Likelihood-ratio test statistic | 15.3 | 2.1 | 0.2 | 3.4 | 7.3 | 8.0 |
| Alanine | 0.96 (0.92, 1.00) | 1.04 (0.98, 1.11) | 1.07 (1.01, 1.14) | 1.00 (0.94, 1.06) | 0.96 (0.90, 1.03) | 0.96 (0.92, 1.00) |
| Likelihood-ratio test statistic | 3.0 | 1.9 | 5.4 | 0.01 | 1.4 | 3.5 |
| Arginine | 0.96 (0.93, 0.99) | 1.02 (0.97, 1.06) | 1.05 (1.00, 1.10) | 0.99 (0.95, 1.04) | 0.97 (0.92, 1.02) | 0.97 (0.94, 1.00) |
| Likelihood-ratio test statistic | 5.6 | 0.4 | 4.0 | 0.1 | 1.5 | 3.7 |
| Aspartic acid | 0.96 (0.93, 0.99) | 1.00 (0.96, 1.04) | 1.03 (0.99, 1.07) | 0.98 (0.94, 1.02) | 0.96 (0.92, 1.00) | 0.97 (0.94, 0.99) |
| Likelihood-ratio test statistic | 8.4 | <0.01 | 1.8 | 1.0 | 3.9 | 5.7 |
| Cystine | 0.93 (0.89, 0.98) | 1.00 (0.93, 1.07) | 1.07 (1.00, 1.15) | 0.99 (0.92, 1.06) | 0.97 (0.90, 1.04) | 0.96 (0.92, 1.01) |
| Likelihood-ratio test statistic | 9.2 | 0.01 | 3.5 | 0.1 | 0.8 | 2.9 |
| Glutamic acid | 0.90 (0.86, 0.94) | 0.91 (0.85, 0.97) | 0.98 (0.92, 1.05) | 0.92 (0.86, 0.98) | 0.90 (0.84, 0.96) | 0.94 (0.90, 0.98) |
| Likelihood-ratio test statistic | 22.9 | 7.9 | 0.4 | 6.6 | 10.1 | 9.7 |
| Glycine | 0.98 (0.94, 1.03) | 1.09 (1.02, 1.15) | 1.10 (1.04, 1.17) | 1.02 (0.96, 1.09) | 0.99 (0.93, 1.06) | 0.97 (0.93, 1.02) |
| Likelihood-ratio test statistic | 0.7 | 6.9 | 10.1 | 0.6 | <0.01 | 1.5 |
| Hydroxyproline | 1.10 (1.03, 1.17) | 1.13 (1.06, 1.20) | 1.09 (1.03, 1.17) | 1.04 (0.98, 1.11) | 1.01 (0.94, 1.08) | 0.99 (0.93, 1.06) |
| Likelihood-ratio test statistic | 8.3 | 12.8 | 7.2 | 1.5 | 0.03 | 0.03 |
| Proline | 0.88 (0.84, 0.92) | 0.88 (0.82, 0.94) | 0.94 (0.88, 1.00) | 0.89 (0.84, 0.95) | 0.88 (0.82, 0.94) | 0.92 (0.88, 0.96) |
| Likelihood-ratio test statistic | 30.7 | 15.6 | 3.6 | 11.8 | 15.1 | 13.7 |
| Serine | 0.93 (0.90, 0.96) | 0.95 (0.90, 1.00) | 1.00 (0.95, 1.05) | 0.94 (0.90, 1.00) | 0.92 (0.88, 0.97) | 0.95 (0.92, 0.98) |
| Likelihood-ratio test statistic | 17.7 | 3.8 | 0.03 | 4.6 | 8.6 | 9.0 |
| Tyrosine | 0.93 (0.90, 0.96) | 0.96 (0.91, 1.01) | 0.99 (0.94, 1.05) | 0.95 (0.90, 1.00) | 0.92 (0.88, 0.98) | 0.94 (0.91, 0.98) |
| Likelihood-ratio test statistic | 15.5 | 2.8 | <0.01 | 4.3 | 8.3 | 9.3 |

1. Model 1 was stratified by sex and centre, and adjusted for age (continuous).
2. Model 2 was model 1 plus calibrated energy intake (continuous)
3. Model 3 was model 2 plus smoking (never, former, current<10, 10-19, 20+ cigarettes/day, unknown), calibrated alcohol consumption (non-drinkers (<0.1), 0.1-4.9, 5.0-14.9, 15-29.9, 30-59.9, 60+ g/day), physical activity (inactive, moderately inactive, moderately active, active, unknown), employment status (employed or student, neither employed nor student, unknown), highest level of education completed (none or primary, secondary, vocational or university, unknown)
4. Model 4 was model 3 plus history of diabetes (yes, no, unknown), prior hypertension (yes, no , unknown), prior hyperlipidaemia (yes, no, unknown)
5. Model 5 was model 4 body mass index (<22.5, 22.5-24.9, 25.0-27.4, 27.5-29.9, ≥30.0 kg/m^2^, unknown)
6. Model 6 was model 5 without calibrated energy intake
7. The χ^2^ statistic from a likelihood ratio test comparing models with and without the amino acid of interest, at the varying levels of covariate adjustment. The changes in the χ^2^ statistic across models could be interpreted as a measure of the extent to which the covariates could account for any associations between dietary amino acids and stroke risk.

Supplementary table 8: Hazard ratios (95% confidence intervals) for **haemorrhagic stroke** (1375 cases) by increments of **calibrated** intakes with **different levels of adjustment**

|  | HR (95% CIs) for per SD of calibrated intakes | | | | | |
| --- | --- | --- | --- | --- | --- | --- |
| Amino acids (percent of total protein) | Model 1^1^ | Model 2^2^ | Model 3^3^ | Model 4^4^ | Model 5^5^ | Model 6^6^ |
| Branched-chain amino acids | 0.98 (0.92, 1.05) | 0.95 (0.86, 1.05) | 0.99 (0.90, 1.08) | 0.96 (0.88, 1.06) | 0.97 (0.88, 1.07) | 1.00 (0.94, 1.07) |
| Likelihood-ratio test statistic^7^ | 0.2 | 1.1 | 0.09 | 0.6 | 0.4 | 0.02 |
| Isoleucine | 0.99 (0.92, 1.05) | 0.95 (0.86, 1.05) | 0.98 (0.89, 1.09) | 0.96 (0.87, 1.06) | 0.97 (0.88, 1.07) | 1.00 (0.94, 1.07) |
| Likelihood-ratio test statistic | 0.2 | 1.0 | 0.09 | 0.6 | 0.4 | 0.02 |
| Leucine | 0.99 (0.92, 1.05) | 0.95 (0.86, 1.05) | 0.99 (0.89, 1.09) | 0.96 (0.87, 1.06) | 0.97 (0.88, 1.07) | 1.01 (0.94, 1.07) |
| Likelihood-ratio test statistic | 0.2 | 1.0 | 0.07 | 0.5 | 0.3 | <0.01 |
| Valine | 0.98 (0.92, 1.05) | 0.95 (0.87, 1.04) | 0.98 (0.90, 1.08) | 0.96 (0.88, 1.06) | 0.97 (0.88, 1.07) | 1.00 (0.94, 1.07) |
| Likelihood-ratio test statistic | 0.3 | 1.3 | 0.12 | 0.6 | 0.4 | 0.02 |
| Other essential amino acids | 0.99 (0.92, 1.06) | 0.96 (0.87, 1.06) | 0.99 (0.90, 1.10) | 0.97 (0.87, 1.07) | 0.97 (0.88, 1.08) | 1.01 (0.94, 1.08) |
| Likelihood-ratio test statistic | 0.09 | 0.6 | 0.024 | 0.4 | 0.2 | <0.01 |
| Histidine | 0.99 (0.92, 1.06) | 0.95 (0.86, 1.06) | 0.98 (0.89, 1.09) | 0.96 (0.86, 1.06) | 0.96 (0.86, 1.07) | 1.00 (0.93, 1.08) |
| Likelihood-ratio test statistic | 0.1 | 0.8 | 0.10 | 0.7 | 0.5 | <0.01 |
| Lysine | 1.00 (0.92, 1.08) | 0.98 (0.88, 1.09) | 1.00 (0.90, 1.12) | 0.97 (0.87, 1.09) | 0.98 (0.88, 1.10) | 1.01 (0.93, 1.10) |
| Likelihood-ratio test statistic | <0.01 | 0.2 | <0.01 | 0.2 | 0.09 | 0.1 |
| Methionine | 0.99 (0.92, 1.07) | 0.97 (0.87, 1.08) | 1.00 (0.89, 1.11) | 0.97 (0.87, 1.08) | 0.98 (0.87, 1.09) | 1.01 (0.93, 1.09) |
| Likelihood-ratio test statistic | 0.04 | 0.4 | <0.01 | 0.3 | 0.2 | 0.1 |
| Phenylalanine | 0.98 (0.92, 1.05) | 0.94 (0.86, 1.04) | 0.98 (0.89, 1.09) | 0.96 (0.87, 1.06) | 0.97 (0.88, 1.07) | 1.01 (0.95, 1.07) |
| Likelihood-ratio test statistic | 0.3 | 1.4 | 0.10 | 0.5 | 0.4 | <0.01 |
| Threonine | 0.99 (0.92, 1.05) | 0.96 (0.87, 1.05) | 0.99 (0.90, 1.09) | 0.96 (0.87, 1.06) | 0.97 (0.88, 1.07) | 1.00 (0.94, 1.07) |
| Likelihood-ratio test statistic | 0.2 | 0.9 | 0.08 | 0.6 | 0.4 | 0.02 |
| Tryptophan | 0.99 (0.93, 1.05) | 0.96 (0.87, 1.05) | 1.00 (0.91, 1.09) | 0.98 (0.89, 1.07) | 0.98 (0.89, 1.08) | 1.01 (0.95, 1.07) |
| Likelihood-ratio test statistic | 0.1 | 0.8 | <0.01 | 0.2 | 0.1 | 0.1 |
| Non-essential amino acids | 0.99 (0.92, 1.05) | 0.95 (0.85, 1.05) | 0.99 (0.89, 1.10) | 0.97 (0.87, 1.07) | 0.97 (0.87, 1.08) | 1.01 (0.95, 1.08) |
| Likelihood-ratio test statistic | 0.2 | 1.1 | 0.03 | 0.4 | 0.2 | 0.1 |
| Alanine | 0.98 (0.91, 1.06) | 0.95 (0.85, 1.06) | 0.97 (0.87, 1.09) | 0.94 (0.84, 1.05) | 0.95 (0.84, 1.06) | 1.00 (0.92, 1.08) |
| Likelihood-ratio test statistic | 0.2 | 0.9 | 0.2 | 1.1 | 0.9 | 0.0 |
| Arginine | 0.98 (0.92, 1.04) | 0.94 (0.86, 1.02) | 0.96 (0.88, 1.05) | 0.94 (0.86, 1.03) | 0.95 (0.86, 1.04) | 0.99 (0.93, 1.05) |
| Likelihood-ratio test statistic | 0.6 | 2.1 | 0.7 | 1.7 | 1.4 | 0.1 |
| Aspartic acid | 1.00 (0.95, 1.05) | 0.99 (0.92, 1.06) | 1.01 (0.94, 1.09) | 0.99 (0.92, 1.07) | 1.00 (0.93, 1.08) | 1.01 (0.97, 1.07) |
| Likelihood-ratio test statistic | <0.01 | 0.1 | 0.13 | 0.03 | <0.01 | 0.3 |
| Cystine | 0.96 (0.88, 1.04) | 0.88 (0.77, 1.00) | 0.94 (0.82, 1.07) | 0.91 (0.80, 1.04) | 0.92 (0.80, 1.05) | 0.99 (0.91, 1.08) |
| Likelihood-ratio test statistic | 1.0 | 3.9 | 0.97 | 1.9 | 1.7 | 0.1 |
| Glutamic acid | 0.98 (0.91, 1.06) | 0.93 (0.83, 1.05) | 0.99 (0.88, 1.12) | 0.97 (0.86, 1.10) | 0.98 (0.86, 1.11) | 1.01 (0.94, 1.09) |
| Likelihood-ratio test statistic | 0.2 | 1.4 | 0.01 | 0.2 | 0.1 | 0.1 |
| Glycine | 0.99 (0.91, 1.07) | 0.95 (0.85, 1.07) | 0.97 (0.87, 1.08) | 0.94 (0.84, 1.05) | 0.94 (0.84, 1.06) | 0.99 (0.92, 1.08) |
| Likelihood-ratio test statistic | 0.1 | 0.7 | 0.30 | 1.2 | 1.0 | 0.03 |
| Hydroxyproline | 1.17 (1.06, 1.30) | 1.18 (1.06, 1.31) | 1.15 (1.03, 1.28) | 1.12 (1.00, 1.25) | 1.14 (1.02, 1.27) | 1.14 (1.02, 1.27) |
| Likelihood-ratio test statistic | 8.6 | 8.5 | 6.0 | 4.0 | 5.0 | 5.5 |
| Proline | 0.97 (0.90, 1.05) | 0.92 (0.82, 1.03) | 0.97 (0.87, 1.09) | 0.96 (0.85, 1.07) | 0.96 (0.86, 1.08) | 1.00 (0.93, 1.08) |
| Likelihood-ratio test statistic | 0.6 | 2.1 | 0.2 | 0.6 | 0.4 | 0.01 |
| Serine | 0.99 (0.93, 1.06) | 0.97 (0.88, 1.06) | 1.01 (0.92, 1.11) | 0.99 (0.90, 1.09) | 1.00 (0.90, 1.10) | 1.02 (0.96, 1.08) |
| Likelihood-ratio test statistic | 0.04 | 0.5 | 0.03 | 0.05 | 0.01 | 0.3 |
| Tyrosine | 0.99 (0.93, 1.06) | 0.97 (0.89, 1.07) | 1.01 (0.92, 1.10) | 0.99 (0.90, 1.08) | 0.99 (0.90, 1.09) | 1.01 (0.95, 1.08) |
| Likelihood-ratio test statistic | 0.03 | 0.3 | 0.01 | 0.08 | 0.02 | 0.2 |

1. Model 1 was stratified by sex and centre, and adjusted for age (continuous).
2. Model 2 was model 1 plus calibrated energy intake (continuous)
3. Model 3 was model 2 plus smoking (never, former, current<10, 10-19, 20+ cigarettes/day, unknown), calibrated alcohol consumption (non-drinkers (<0.1), 0.1-4.9, 5.0-14.9, 15-29.9, 30-59.9, 60+ g/day), physical activity (inactive, moderately inactive, moderately active, active, unknown), employment status (employed or student, neither employed nor student, unknown), highest level of education completed (none or primary, secondary, vocational or university, unknown)
4. Model 4 was model 3 plus history of diabetes (yes, no, unknown), prior hypertension (yes, no , unknown), prior hyperlipidaemia (yes, no, unknown)
5. Model 5 was model 4 body mass index (<22.5, 22.5-24.9, 25.0-27.4, 27.5-29.9, ≥30.0 kg/m^2^, unknown)
6. Model 6 was model 5 without calibrated energy intake
7. The χ^2^ statistic from a likelihood ratio test comparing models with and without the amino acid of interest, at the varying levels of covariate adjustment. The changes in the χ^2^ statistic across models could be interpreted as a measure of the extent to which the covariates could account for any associations between dietary amino acids and stroke risk

Supplementary table 9: Hazard ratios (95% confidence intervals)^1^ for **ischaemic** (3603 cases) and **haemorrhagic** stroke (1146 cases) by increments of calibrated intakes of dietary amino acids, adjusting for major macronutrients.

| Amino acids (percent of total protein) | Ischaemic stroke | |  | Haemorrhagic stroke | |
| --- | --- | --- | --- | --- | --- |
|  | HR (95% CI) | p-trend^2^ |  | HR (95% CI) | p-trend^2^ |
| Branched-chain amino acids | 0.88 (0.79, 0.99) | 0.03 |  | 0.99 (0.81, 1.22) | 0.94 |
| Isoleucine | 0.89 (0.79, 1.00) | 0.05 |  | 0.98 (0.79, 1.21) | 0.86 |
| Leucine | 0.89 (0.79, 1.00) | 0.04 |  | 1.00 (0.81, 1.24) | 1.00 |
| Valine | 0.88 (0.80, 0.98) | 0.02 |  | 0.99 (0.82, 1.20) | 0.94 |
| Other essential amino acids | 0.91 (0.80, 1.03) | 0.15 |  | 1.01 (0.80, 1.27) | 0.93 |
| Histidine | 0.94 (0.83, 1.07) | 0.38 |  | 0.96 (0.76, 1.21) | 0.72 |
| Lysine | 0.94 (0.82, 1.07) | 0.36 |  | 1.06 (0.84, 1.35) | 0.61 |
| Methionine | 0.93 (0.82, 1.06) | 0.29 |  | 1.04 (0.82, 1.30) | 0.77 |
| Phenylalanine | 0.89 (0.80, 0.99) | 0.04 |  | 0.99 (0.81, 1.21) | 0.92 |
| Threonine | 0.90 (0.80, 1.01) | 0.08 |  | 0.96 (0.78, 1.19) | 0.73 |
| Tryptophan | 0.92 (0.84, 1.02) | 0.11 |  | 1.03 (0.86, 1.23) | 0.75 |
| Non-essential amino acids | 0.89 (0.80, 1.00) | 0.06 |  | 1.00 (0.80, 1.24) | 0.97 |
| Alanine | 1.04 (0.92, 1.17) | 0.57 |  | 0.87 (0.70, 1.08) | 0.20 |
| Arginine | 1.01 (0.92, 1.10) | 0.87 |  | 0.85 (0.73, 1.00) | 0.05 |
| Aspartic acid | 0.97 (0.90, 1.05) | 0.51 |  | 1.04 (0.90, 1.21) | 0.56 |
| Cystine | 1.03 (0.94, 1.14) | 0.51 |  | 0.85 (0.70, 1.03) | 0.09 |
| Glutamic acid | 0.88 (0.79, 0.98) | 0.02 |  | 1.03 (0.84, 1.26) | 0.76 |
| Glycine | 1.08 (0.98, 1.20) | 0.12 |  | 0.86 (0.71, 1.04) | 0.12 |
| Hydroxyproline | 1.02 (0.95, 1.10) | 0.61 |  | 1.16 (1.03, 1.31) | 0.01 |
| Proline | 0.85 (0.78, 0.94) | 0.001 |  | 1.01 (0.85, 1.20) | 0.90 |
| Serine | 0.90 (0.81, 0.99) | 0.03 |  | 1.09 (0.91, 1.30) | 0.37 |
| Tyrosine | 0.89 (0.80, 0.98) | 0.02 |  | 1.10 (0.91, 1.33) | 0.32 |

1. Hazard ratios modelled per 1 sex-specific SD increment in dietary amino acids, expressed as percent of total protein. Based on multivariable model stratified by sex and centre, and adjusted for age (continuous), smoking (never, former, current<10, 10-19, 20+ cigarettes/day, unknown), calibrated alcohol consumption (non-drinkers (<0.1), 0.1-4.9, 5.0-14.9, 15-29.9, 30-59.9, 60+ g/day), physical activity (inactive, moderately inactive, moderately active, active, unknown), employment status (employed or student, neither employed nor student, unknown), highest level of education completed (none or primary, secondary, vocational or university, unknown), history of diabetes (yes, no, unknown), prior hypertension (yes, no , unknown), prior hyperlipidaemia (yes, no, unknown), body mass index (<22.5, 22.5-24.9, 25.0-27.4, 27.5-29.9, ≥30.0 kg/m^2^, unknown), calibrated intakes of total protein (continuous), carbohydrates (continuous), saturated fats (continuous) and unsaturated fats (continuous).

Supplementary table 10: Adjusted means (95% confidence intervals) of **systolic blood pressure** by observed fifths of dietary amino acids (n=267,642)

| Amino acids (percent of total protein) | Q1 | Q2 | Q3 | Q4 | Q5 | p-trend |
| --- | --- | --- | --- | --- | --- | --- |
| Branched-chain amino acids |  |  |  |  |  |  |
| Model 1 | 131.2 (131.0, 131.3) | 131.5 (131.3, 131.6) | 132.3 (132.1, 132.4) | 132.6 (132.5, 132.7) | 132.5 (132.3, 132.6) | <0.001 |
| Model 2 | 131.5 (131.3, 131.7) | 131.6 (131.4, 131.8) | 132.2 (132.1, 132.3) | 132.4 (132.3, 132.6) | 132.3 (132.2, 132.5) | <0.001 |
| Model 3 | 131.4 (131.3, 131.6) | 131.5 (131.3, 131.6) | 132.1 (131.9, 132.2) | 132.5 (132.3, 132.6) | 132.6 (132.4, 132.7) | <0.001 |
| Model 4 | 131.8 (131.6, 132.0) | 131.5 (131.3, 131.6) | 132.0 (131.8, 132.1) | 132.4 (132.2, 132.5) | 132.5 (132.4, 132.7) | <0.001 |
| Isoleucine |  |  |  |  |  |  |
| Model 1 | 131.0 (130.8, 131.2) | 131.4 (131.3, 131.6) | 132.1 (131.9, 132.2) | 132.7 (132.6, 132.9) | 132.7 (132.6, 132.9) | <0.001 |
| Model 2 | 131.4 (131.2, 131.5) | 131.6 (131.4, 131.7) | 132.0 (131.9, 132.2) | 132.5 (132.4, 132.7) | 132.6 (132.5, 132.8) | <0.001 |
| Model 3 | 131.2 (131.1, 131.4) | 131.4 (131.3, 131.6) | 132.0 (131.8, 132.1) | 132.6 (132.4, 132.7) | 132.8 (132.7, 132.9) | <0.001 |
| Model 4 | 131.6 (131.5, 131.8) | 131.4 (131.3, 131.5) | 131.9 (131.7, 132.0) | 132.5 (132.3, 132.6) | 132.7 (132.6, 132.9) | <0.001 |
| Leucine |  |  |  |  |  |  |
| Model 1 | 131.1 (130.9, 131.3) | 131.4 (131.2, 131.6) | 132.2 (132.1, 132.4) | 132.6 (132.5, 132.8) | 132.5 (132.4, 132.7) | <0.001 |
| Model 2 | 131.5 (131.3, 131.6) | 131.5 (131.4, 131.7) | 132.2 (132.0, 132.3) | 132.5 (132.3, 132.6) | 132.4 (132.3, 132.5) | <0.001 |
| Model 3 | 131.4 (131.3, 131.6) | 131.4 (131.3, 131.6) | 132.1 (131.9, 132.2) | 132.5 (132.4, 132.6) | 132.6 (132.4, 132.7) | <0.001 |
| Model 4 | 131.8 (131.6, 131.9) | 131.4 (131.3, 131.6) | 131.9 (131.8, 132.1) | 132.4 (132.2, 132.5) | 132.6 (132.4, 132.7) | <0.001 |
| Valine |  |  |  |  |  |  |
| Model 1 | 131.3 (131.1, 131.5) | 131.7 (131.6, 131.9) | 132.4 (132.2, 132.5) | 132.6 (132.4, 132.7) | 132.1 (132.0, 132.2) | <0.001 |
| Model 2 | 131.6 (131.5, 131.8) | 131.8 (131.7, 132.0) | 132.3 (132.1, 132.4) | 132.4 (132.3, 132.6) | 132.0 (131.9, 132.2) | <0.001 |
| Model 3 | 131.5 (131.4, 131.7) | 131.7 (131.5, 131.8) | 132.2 (132.0, 132.3) | 132.4 (132.3, 132.6) | 132.3 (132.2, 132.4) | <0.001 |
| Model 4 | 131.8 (131.7, 132.0) | 131.7 (131.5, 131.8) | 132.0 (131.9, 132.2) | 132.3 (132.2, 132.5) | 132.3 (132.2, 132.4) | <0.001 |
| Other essential amino acids |  |  |  |  |  |  |
| Model 1 | 130.6 (130.5, 130.8) | 131.2 (131.1, 131.4) | 131.9 (131.8, 132.1) | 132.6 (132.5, 132.8) | 133.2 (133.1, 133.4) | <0.001 |
| Model 2 | 131.0 (130.9, 131.2) | 131.4 (131.2, 131.5) | 131.9 (131.7, 132.0) | 132.5 (132.3, 132.6) | 133.0 (132.9, 133.2) | <0.001 |
| Model 3 | 131.1 (130.9, 131.2) | 131.3 (131.2, 131.5) | 131.8 (131.7, 132.0) | 132.5 (132.4, 132.7) | 133.0 (132.9, 133.2) | <0.001 |
| Model 4 | 131.5 (131.4, 131.7) | 131.3 (131.2, 131.5) | 131.8 (131.6, 131.9) | 132.5 (132.3, 132.6) | 132.9 (132.8, 133.0) | <0.001 |
| Histidine |  |  |  |  |  |  |
| Model 1 | 130.6 (130.4, 130.8) | 131.2 (131.1, 131.4) | 131.9 (131.8, 132.0) | 132.7 (132.6, 132.9) | 133.1 (133.0, 133.3) | <0.001 |
| Model 2 | 131.0 (130.8, 131.1) | 131.4 (131.2, 131.5) | 131.9 (131.7, 132.0) | 132.6 (132.5, 132.7) | 132.9 (132.8, 133.1) | <0.001 |
| Model 3 | 131.1 (130.9, 131.3) | 131.4 (131.2, 131.5) | 131.8 (131.7, 132.0) | 132.7 (132.5, 132.8) | 132.8 (132.7, 133.0) | <0.001 |
| Model 4 | 131.6 (131.5, 131.8) | 131.4 (131.2, 131.5) | 131.7 (131.6, 131.9) | 132.5 (132.4, 132.7) | 132.7 (132.6, 132.8) | <0.001 |
| Lysine |  |  |  |  |  |  |
| Model 1 | 130.2 (130.0, 130.3) | 131.2 (131.0, 131.3) | 131.9 (131.8, 132.1) | 132.8 (132.7, 133.0) | 133.5 (133.4, 133.7) | <0.001 |
| Model 2 | 130.5 (130.3, 130.7) | 131.3 (131.2, 131.5) | 131.9 (131.8, 132.0) | 132.7 (132.6, 132.8) | 133.3 (133.2, 133.5) | <0.001 |
| Model 3 | 130.6 (130.4, 130.7) | 131.3 (131.2, 131.4) | 132.0 (131.8, 132.1) | 132.7 (132.6, 132.9) | 133.2 (133.1, 133.4) | <0.001 |
| Model 4 | 131.1 (130.9, 131.3) | 131.4 (131.2, 131.5) | 131.9 (131.8, 132.0) | 132.6 (132.5, 132.7) | 133.0 (132.8, 133.1) | <0.001 |
| Methionine |  |  |  |  |  |  |
| Model 1 | 130.4 (130.2, 130.6) | 131.1 (131.0, 131.3) | 131.8 (131.6, 131.9) | 132.7 (132.6, 132.8) | 133.5 (133.4, 133.6) | <0.001 |
| Model 2 | 130.9 (130.7, 131.0) | 131.3 (131.1, 131.4) | 131.7 (131.5, 131.8) | 132.5 (132.4, 132.6) | 133.3 (133.2, 133.5) | <0.001 |
| Model 3 | 131.0 (130.8, 131.1) | 131.3 (131.1, 131.4) | 131.7 (131.6, 131.8) | 132.5 (132.4, 132.7) | 133.3 (133.1, 133.4) | <0.001 |
| Model 4 | 131.5 (131.3, 131.7) | 131.3 (131.1, 131.4) | 131.6 (131.5, 131.8) | 132.4 (132.3, 132.6) | 133.1 (132.9, 133.2) | <0.001 |
| Phenylalanine |  |  |  |  |  |  |
| Model 1 | 131.5 (131.3, 131.7) | 131.8 (131.7, 132.0) | 132.4 (132.2, 132.5) | 132.6 (132.4, 132.7) | 131.9 (131.8, 132.1) | <0.001 |
| Model 2 | 131.9 (131.7, 132.0) | 132.0 (131.8, 132.1) | 132.3 (132.2, 132.5) | 132.4 (132.3, 132.5) | 131.8 (131.7, 131.9) | <0.001 |
| Model 3 | 131.8 (131.7, 132.0) | 131.9 (131.7, 132.0) | 132.1 (132.0, 132.3) | 132.3 (132.1, 132.4) | 132.1 (132.0, 132.2) | <0.001 |
| Model 4 | 132.1 (131.9, 132.3) | 131.8 (131.7, 132.0) | 132.0 (131.9, 132.1) | 132.1 (132.0, 132.3) | 132.2 (132.1, 132.3) | 0.003 |
| Threonine |  |  |  |  |  |  |
| Model 1 | 130.7 (130.5, 130.9) | 131.2 (131.1, 131.4) | 131.9 (131.8, 132.1) | 132.7 (132.5, 132.8) | 133.3 (133.2, 133.4) | <0.001 |
| Model 2 | 131.1 (131.0, 131.3) | 131.4 (131.2, 131.5) | 131.9 (131.7, 132.0) | 132.5 (132.3, 132.6) | 133.1 (133.0, 133.3) | <0.001 |
| Model 3 | 131.0 (130.9, 131.2) | 131.3 (131.1, 131.4) | 131.8 (131.7, 132.0) | 132.5 (132.4, 132.7) | 133.2 (133.1, 133.4) | <0.001 |
| Model 4 | 131.4 (131.3, 131.6) | 131.3 (131.1, 131.4) | 131.8 (131.6, 131.9) | 132.4 (132.3, 132.6) | 133.1 (132.9, 133.2) | <0.001 |
| Tryptophan |  |  |  |  |  |  |
| Model 1 | 131.2 (131.1, 131.4) | 132.1 (131.9, 132.2) | 132.2 (132.0, 132.3) | 132.2 (132.0, 132.3) | 132.5 (132.4, 132.7) | <0.001 |
| Model 2 | 131.7 (131.5, 131.9) | 132.1 (131.9, 132.2) | 132.1 (131.9, 132.2) | 132.1 (132.0, 132.3) | 132.3 (132.1, 132.4) | <0.001 |
| Model 3 | 131.4 (131.3, 131.6) | 132.0 (131.8, 132.1) | 132.1 (132.0, 132.2) | 132.2 (132.1, 132.4) | 132.4 (132.3, 132.6) | <0.001 |
| Model 4 | 131.5 (131.4, 131.7) | 131.9 (131.8, 132.1) | 132.0 (131.9, 132.2) | 132.2 (132.0, 132.3) | 132.5 (132.4, 132.7) | <0.001 |
| Non-essential amino acids |  |  |  |  |  |  |
| Model 1 | 131.1 (130.9, 131.3) | 131.5 (131.4, 131.7) | 132.2 (132.0, 132.3) | 132.7 (132.6, 132.8) | 132.4 (132.3, 132.5) | <0.001 |
| Model 2 | 131.6 (131.4, 131.8) | 131.8 (131.6, 131.9) | 132.1 (132.0, 132.3) | 132.4 (132.3, 132.6) | 132.2 (132.1, 132.3) | <0.001 |
| Model 3 | 131.7 (131.5, 131.8) | 131.7 (131.6, 131.9) | 131.9 (131.8, 132.0) | 132.3 (132.2, 132.5) | 132.5 (132.3, 132.6) | <0.001 |
| Model 4 | 131.9 (131.8, 132.1) | 131.7 (131.5, 131.8) | 131.8 (131.7, 131.9) | 132.2 (132.1, 132.3) | 132.5 (132.4, 132.6) | <0.001 |
| Alanine |  |  |  |  |  |  |
| Model 1 | 130.2 (130.0, 130.4) | 131.0 (130.9, 131.2) | 131.9 (131.7, 132.0) | 132.8 (132.7, 133.0) | 133.8 (133.7, 134.0) | <0.001 |
| Model 2 | 130.7 (130.5, 130.9) | 131.2 (131.1, 131.4) | 131.8 (131.7, 132.0) | 132.6 (132.5, 132.8) | 133.6 (133.4, 133.7) | <0.001 |
| Model 3 | 130.8 (130.7, 131.0) | 131.2 (131.1, 131.3) | 131.8 (131.6, 131.9) | 132.6 (132.5, 132.7) | 133.5 (133.4, 133.7) | <0.001 |
| Model 4 | 131.3 (131.2, 131.5) | 131.3 (131.1, 131.4) | 131.7 (131.6, 131.9) | 132.5 (132.3, 132.6) | 133.3 (133.1, 133.4) | <0.001 |
| Arginine |  |  |  |  |  |  |
| Model 1 | 130.8 (130.7, 131.0) | 131.5 (131.4, 131.7) | 132.0 (131.9, 132.2) | 132.7 (132.6, 132.9) | 133.1 (133.0, 133.3) | <0.001 |
| Model 2 | 131.2 (131.0, 131.3) | 131.7 (131.5, 131.8) | 132.0 (131.9, 132.1) | 132.6 (132.4, 132.7) | 132.9 (132.7, 133.0) | <0.001 |
| Model 3 | 131.3 (131.2, 131.5) | 131.7 (131.5, 131.8) | 131.9 (131.7, 132.0) | 132.5 (132.3, 132.6) | 132.9 (132.8, 133.1) | <0.001 |
| Model 4 | 131.7 (131.5, 131.8) | 131.7 (131.6, 131.9) | 131.8 (131.7, 132.0) | 132.3 (132.2, 132.5) | 132.7 (132.6, 132.8) | <0.001 |
| Aspartic acid |  |  |  |  |  |  |
| Model 1 | 130.7 (130.5, 130.8) | 131.6 (131.4, 131.7) | 132.4 (132.3, 132.6) | 132.7 (132.6, 132.9) | 132.8 (132.6, 132.9) | <0.001 |
| Model 2 | 131.0 (130.8, 131.1) | 131.7 (131.5, 131.8) | 132.3 (132.1, 132.4) | 132.6 (132.4, 132.7) | 132.7 (132.6, 132.9) | <0.001 |
| Model 3 | 130.8 (130.6, 130.9) | 131.5 (131.3, 131.6) | 132.2 (132.1, 132.3) | 132.7 (132.5, 132.8) | 133.1 (132.9, 133.2) | <0.001 |
| Model 4 | 131.1 (131.0, 131.3) | 131.5 (131.3, 131.6) | 132.1 (132.0, 132.3) | 132.6 (132.5, 132.7) | 132.9 (132.8, 133.1) | <0.001 |
| Cystine |  |  |  |  |  |  |
| Model 1 | 131.5 (131.3, 131.6) | 132.2 (132.1, 132.4) | 133.1 (133.0, 133.3) | 132.3 (132.2, 132.4) | 131.3 (131.1, 131.4) | <0.001 |
| Model 2 | 132.0 (131.8, 132.1) | 132.4 (132.3, 132.6) | 132.9 (132.8, 133.1) | 132.1 (131.9, 132.2) | 131.1 (131.0, 131.3) | <0.001 |
| Model 3 | 132.2 (132.0, 132.4) | 132.4 (132.3, 132.6) | 132.7 (132.6, 132.9) | 131.9 (131.8, 132.0) | 131.3 (131.1, 131.4) | <0.001 |
| Model 4 | 132.5 (132.3, 132.6) | 132.3 (132.2, 132.5) | 132.6 (132.5, 132.8) | 131.8 (131.7, 132.0) | 131.3 (131.2, 131.5) | <0.001 |
| Glutamic acid |  |  |  |  |  |  |
| Model 1 | 131.5 (131.4, 131.7) | 131.8 (131.6, 132.0) | 132.7 (132.5, 132.8) | 132.5 (132.4, 132.6) | 131.7 (131.6, 131.8) | <0.001 |
| Model 2 | 132.0 (131.8, 132.2) | 132.1 (131.9, 132.2) | 132.6 (132.4, 132.7) | 132.3 (132.2, 132.4) | 131.5 (131.4, 131.6) | <0.001 |
| Model 3 | 132.1 (131.9, 132.2) | 132.1 (131.9, 132.2) | 132.3 (132.2, 132.5) | 132.2 (132.0, 132.3) | 131.8 (131.6, 131.9) | <0.001 |
| Model 4 | 132.2 (132.0, 132.4) | 132.0 (131.8, 132.1) | 132.2 (132.0, 132.3) | 132.1 (132.0, 132.2) | 132.0 (131.9, 132.1) | 0.08 |
| Glycine |  |  |  |  |  |  |
| Model 1 | 130.3 (130.1, 130.4) | 131.1 (131.0, 131.3) | 131.8 (131.7, 132.0) | 132.7 (132.5, 132.8) | 133.8 (133.7, 134.0) | <0.001 |
| Model 2 | 130.8 (130.6, 130.9) | 131.4 (131.2, 131.5) | 131.8 (131.7, 132.0) | 132.5 (132.3, 132.6) | 133.5 (133.3, 133.6) | <0.001 |
| Model 3 | 131.1 (130.9, 131.2) | 131.4 (131.3, 131.6) | 131.8 (131.6, 131.9) | 132.3 (132.2, 132.5) | 133.3 (133.2, 133.5) | <0.001 |
| Model 4 | 131.6 (131.4, 131.7) | 131.5 (131.4, 131.7) | 131.7 (131.6, 131.9) | 132.2 (132.1, 132.3) | 133.1 (132.9, 133.2) | <0.001 |
| Hydroxyproline |  |  |  |  |  |  |
| Model 1 | 130.3 (130.1, 130.4) | 131.4 (131.3, 131.6) | 132.1 (131.9, 132.2) | 132.6 (132.5, 132.8) | 132.9 (132.8, 133.1) | <0.001 |
| Model 2 | 130.4 (130.2, 130.6) | 131.4 (131.2, 131.5) | 132.0 (131.8, 132.1) | 132.6 (132.4, 132.7) | 133.0 (132.9, 133.2) | <0.001 |
| Model 3 | 131.0 (130.8, 131.2) | 131.5 (131.4, 131.7) | 132.0 (131.9, 132.2) | 132.5 (132.3, 132.6) | 132.7 (132.6, 132.8) | <0.001 |
| Model 4 | 131.7 (131.5, 131.9) | 131.7 (131.6, 131.8) | 132.0 (131.9, 132.2) | 132.3 (132.2, 132.5) | 132.3 (132.2, 132.5) | <0.001 |
| Proline |  |  |  |  |  |  |
| Model 1 | 131.7 (131.5, 131.8) | 132.1 (131.9, 132.2) | 132.4 (132.3, 132.6) | 132.4 (132.3, 132.5) | 131.7 (131.6, 131.9) | <0.001 |
| Model 2 | 132.0 (131.8, 132.2) | 132.3 (132.1, 132.4) | 132.4 (132.2, 132.5) | 132.2 (132.1, 132.4) | 131.5 (131.4, 131.7) | <0.001 |
| Model 3 | 132.1 (132.0, 132.3) | 132.2 (132.1, 132.4) | 132.2 (132.1, 132.4) | 132.1 (132.0, 132.3) | 131.7 (131.6, 131.9) | <0.001 |
| Model 4 | 132.2 (132.0, 132.4) | 132.1 (132.0, 132.3) | 132.1 (132.0, 132.2) | 132.1 (132.0, 132.2) | 131.9 (131.8, 132.0) | 0.06 |
| Serine |  |  |  |  |  |  |
| Model 1 | 131.4 (131.3, 131.6) | 131.8 (131.7, 132.0) | 132.3 (132.2, 132.5) | 132.6 (132.4, 132.7) | 131.9 (131.8, 132.1) | <0.001 |
| Model 2 | 131.8 (131.6, 132.0) | 132.0 (131.8, 132.2) | 132.3 (132.1, 132.4) | 132.4 (132.2, 132.5) | 131.9 (131.7, 132.0) | <0.001 |
| Model 3 | 131.8 (131.6, 132.0) | 131.9 (131.8, 132.1) | 132.1 (131.9, 132.2) | 132.3 (132.2, 132.5) | 132.1 (132.0, 132.2) | <0.001 |
| Model 4 | 132.1 (131.9, 132.2) | 131.9 (131.7, 132.0) | 131.9 (131.8, 132.1) | 132.2 (132.1, 132.3) | 132.2 (132.1, 132.3) | 0.003 |
| Tyrosine |  |  |  |  |  |  |
| Model 1 | 131.2 (131.0, 131.3) | 131.7 (131.5, 131.8) | 132.1 (131.9, 132.2) | 132.5 (132.4, 132.7) | 132.5 (132.3, 132.6) | <0.001 |
| Model 2 | 131.4 (131.2, 131.6) | 131.8 (131.6, 131.9) | 132.0 (131.9, 132.2) | 132.4 (132.3, 132.6) | 132.4 (132.2, 132.5) | <0.001 |
| Model 3 | 131.4 (131.2, 131.5) | 131.7 (131.5, 131.8) | 132.0 (131.9, 132.2) | 132.4 (132.3, 132.6) | 132.5 (132.3, 132.6) | <0.001 |
| Model 4 | 131.7 (131.6, 131.9) | 131.7 (131.5, 131.8) | 131.9 (131.8, 132.0) | 132.3 (132.2, 132.4) | 132.5 (132.3, 132.6) | <0.001 |
| Total protein (g/1000 kcal) |  |  |  |  |  |  |
| Model 1 | 131.2 (131.0, 131.3) | 131.5 (131.4, 131.7) | 132.1 (132.0, 132.3) | 132.5 (132.3, 132.6) | 133.1 (132.9, 133.3) | <0.001 |
| Model 2 | 130.7 (130.5, 130.8) | 131.4 (131.2, 131.5) | 132.2 (132.0, 132.3) | 132.7 (132.5, 132.8) | 133.5 (133.3, 133.6) | <0.001 |
| Model 3 | 131.2 (131.0, 131.3) | 131.5 (131.4, 131.7) | 132.2 (132.0, 132.3) | 132.5 (132.4, 132.7) | 133.0 (132.9, 133.2) | <0.001 |
| Model 4 | 131.6 (131.5, 131.8) | 131.8 (131.6, 131.9) | 132.2 (132.1, 132.3) | 132.3 (132.2, 132.5) | 132.4 (132.3, 132.6) | <0.001 |

Model 1 was adjusted for age (continuous), sex, centre
Model 2 was model 1 plus smoking (never, former, current<10, 10-19, 20+ cigarettes/day, unknown), current alcohol consumption (non-drinkers, 0.1-4.9, 5.0-14.9, 15-29.9, 30-59.9, 60+ g/day), physical activity (inactive, moderately inactive, moderately active, active, unknown), employment status (employed or student, neither employed nor student, unknown), highest level of education completed (none or primary, secondary, vocational or university, unknown),calibrated energy intake (continuous)
Model 3 was model 2 plus history of diabetes (yes, no, unknown), prior hypertension (yes, no , unknown), prior hyperlipidaemia (yes, no, unknown)
Model 4 was model 3 plus body mass index (<22.5, 22.5-24.9, 25.0-27.4, 27.5-29.9, ≥30.0 kg/m^2^, unknown)

Supplementary table 11: Adjusted means (95% confidence intervals) of **diastolic blood pressure** by observed fifths of dietary amino acids (n=267,642)

| Amino acids (percent of total protein) | Q1 | Q2 | Q3 | Q4 | Q5 | p-trend |
| --- | --- | --- | --- | --- | --- | --- |
| Branched-chain amino acids |  |  |  |  |  |  |
| Model 1 | 80.4 (80.3, 80.5) | 81.5 (81.4, 81.6) | 82.0 (81.9, 82.1) | 81.7 (81.6, 81.8) | 81.4 (81.4, 81.5) | <0.001 |
| Model 2 | 80.7 (80.6, 80.8) | 81.6 (81.5, 81.7) | 81.9 (81.8, 82.0) | 81.6 (81.5, 81.7) | 81.4 (81.3, 81.4) | <0.001 |
| Model 3 | 80.7 (80.6, 80.8) | 81.5 (81.4, 81.6) | 81.8 (81.8, 81.9) | 81.6 (81.5, 81.7) | 81.5 (81.4, 81.6) | <0.001 |
| Model 4 | 80.9 (80.9, 81.0) | 81.5 (81.4, 81.6) | 81.8 (81.7, 81.8) | 81.5 (81.5, 81.6) | 81.5 (81.4, 81.6) | <0.001 |
| Isoleucine |  |  |  |  |  |  |
| Model 1 | 80.5 (80.4, 80.6) | 81.6 (81.5, 81.7) | 81.8 (81.7, 81.9) | 81.7 (81.6, 81.8) | 81.5 (81.5, 81.6) | <0.001 |
| Model 2 | 80.8 (80.7, 80.9) | 81.7 (81.6, 81.8) | 81.8 (81.7, 81.8) | 81.6 (81.5, 81.7) | 81.4 (81.4, 81.5) | <0.001 |
| Model 3 | 80.7 (80.6, 80.8) | 81.5 (81.5, 81.6) | 81.7 (81.6, 81.8) | 81.6 (81.6, 81.7) | 81.6 (81.5, 81.7) | <0.001 |
| Model 4 | 81.0 (80.9, 81.1) | 81.5 (81.5, 81.6) | 81.6 (81.6, 81.7) | 81.5 (81.5, 81.6) | 81.5 (81.5, 81.6) | <0.001 |
| Leucine |  |  |  |  |  |  |
| Model 1 | 80.3 (80.2, 80.4) | 81.4 (81.4, 81.5) | 82.0 (81.9, 82.1) | 81.7 (81.7, 81.8) | 81.5 (81.4, 81.6) | <0.001 |
| Model 2 | 80.6 (80.5, 80.7) | 81.5 (81.5, 81.6) | 81.9 (81.8, 82.0) | 81.6 (81.5, 81.7) | 81.4 (81.3, 81.5) | <0.001 |
| Model 3 | 80.6 (80.5, 80.7) | 81.5 (81.4, 81.5) | 81.9 (81.8, 81.9) | 81.6 (81.6, 81.7) | 81.5 (81.5, 81.6) | <0.001 |
| Model 4 | 80.9 (80.8, 81.0) | 81.5 (81.4, 81.5) | 81.8 (81.7, 81.8) | 81.5 (81.5, 81.6) | 81.5 (81.5, 81.6) | <0.001 |
| Valine |  |  |  |  |  |  |
| Model 1 | 80.6 (80.5, 80.7) | 81.6 (81.5, 81.7) | 82.0 (81.9, 82.1) | 81.8 (81.7, 81.9) | 81.2 (81.2, 81.3) | <0.001 |
| Model 2 | 80.8 (80.7, 80.9) | 81.6 (81.5, 81.7) | 81.9 (81.8, 82.0) | 81.7 (81.6, 81.7) | 81.2 (81.1, 81.3) | <0.001 |
| Model 3 | 80.8 (80.7, 80.9) | 81.5 (81.4, 81.6) | 81.8 (81.8, 81.9) | 81.7 (81.6, 81.8) | 81.4 (81.3, 81.4) | <0.001 |
| Model 4 | 81.0 (80.9, 81.1) | 81.5 (81.4, 81.6) | 81.8 (81.7, 81.8) | 81.6 (81.5, 81.7) | 81.4 (81.3, 81.5) | <0.001 |
| Other essential amino acids |  |  |  |  |  |  |
| Model 1 | 80.1 (80.0, 80.2) | 81.3 (81.3, 81.4) | 81.7 (81.6, 81.8) | 81.7 (81.6, 81.8) | 82.0 (81.9, 82.1) | <0.001 |
| Model 2 | 80.4 (80.3, 80.5) | 81.5 (81.4, 81.5) | 81.7 (81.6, 81.7) | 81.6 (81.5, 81.7) | 81.9 (81.8, 82.0) | <0.001 |
| Model 3 | 80.4 (80.3, 80.5) | 81.4 (81.3, 81.5) | 81.6 (81.5, 81.7) | 81.6 (81.6, 81.7) | 81.9 (81.8, 82.0) | <0.001 |
| Model 4 | 80.8 (80.7, 80.9) | 81.4 (81.3, 81.5) | 81.6 (81.5, 81.6) | 81.6 (81.5, 81.6) | 81.8 (81.7, 81.9) | <0.001 |
| Histidine |  |  |  |  |  |  |
| Model 1 | 79.9 (79.8, 80.0) | 81.3 (81.2, 81.4) | 81.7 (81.6, 81.8) | 81.7 (81.6, 81.8) | 82.2 (82.1, 82.2) | <0.001 |
| Model 2 | 80.2 (80.1, 80.3) | 81.4 (81.3, 81.5) | 81.7 (81.6, 81.7) | 81.6 (81.5, 81.7) | 82.0 (81.9, 82.1) | <0.001 |
| Model 3 | 80.3 (80.2, 80.4) | 81.4 (81.3, 81.5) | 81.6 (81.5, 81.7) | 81.6 (81.5, 81.7) | 82.0 (81.9, 82.0) | <0.001 |
| Model 4 | 80.7 (80.6, 80.8) | 81.4 (81.3, 81.5) | 81.5 (81.5, 81.6) | 81.5 (81.5, 81.6) | 81.9 (81.8, 81.9) | <0.001 |
| Lysine |  |  |  |  |  |  |
| Model 1 | 80.3 (80.2, 80.4) | 81.2 (81.1, 81.3) | 81.4 (81.3, 81.5) | 81.7 (81.7, 81.8) | 82.3 (82.2, 82.4) | <0.001 |
| Model 2 | 80.5 (80.4, 80.6) | 81.4 (81.3, 81.4) | 81.4 (81.3, 81.4) | 81.6 (81.6, 81.7) | 82.2 (82.1, 82.2) | <0.001 |
| Model 3 | 80.5 (80.4, 80.6) | 81.3 (81.2, 81.4) | 81.4 (81.3, 81.5) | 81.7 (81.6, 81.8) | 82.1 (82.0, 82.2) | <0.001 |
| Model 4 | 80.9 (80.8, 81.0) | 81.3 (81.3, 81.4) | 81.4 (81.3, 81.4) | 81.6 (81.5, 81.7) | 81.9 (81.9, 82.0) | <0.001 |
| Methionine |  |  |  |  |  |  |
| Model 1 | 79.7 (79.6, 79.8) | 81.1 (81.1, 81.2) | 81.6 (81.5, 81.7) | 82.0 (81.9, 82.0) | 82.2 (82.1, 82.3) | <0.001 |
| Model 2 | 80.0 (79.9, 80.1) | 81.3 (81.2, 81.4) | 81.6 (81.5, 81.7) | 81.8 (81.7, 81.9) | 82.1 (82.0, 82.2) | <0.001 |
| Model 3 | 80.1 (80.0, 80.2) | 81.2 (81.1, 81.3) | 81.5 (81.5, 81.6) | 81.8 (81.8, 81.9) | 82.1 (82.0, 82.2) | <0.001 |
| Model 4 | 80.5 (80.4, 80.6) | 81.3 (81.2, 81.3) | 81.5 (81.4, 81.6) | 81.8 (81.7, 81.8) | 81.9 (81.9, 82.0) | <0.001 |
| Phenylalanine |  |  |  |  |  |  |
| Model 1 | 80.4 (80.3, 80.5) | 81.4 (81.3, 81.5) | 82.2 (82.1, 82.3) | 82.0 (81.9, 82.1) | 81.1 (81.1, 81.2) | <0.001 |
| Model 2 | 80.7 (80.6, 80.8) | 81.5 (81.4, 81.6) | 82.1 (82.0, 82.2) | 81.9 (81.8, 82.0) | 81.1 (81.0, 81.1) | <0.001 |
| Model 3 | 80.7 (80.6, 80.8) | 81.5 (81.4, 81.5) | 82.0 (81.9, 82.1) | 81.8 (81.7, 81.9) | 81.2 (81.2, 81.3) | <0.001 |
| Model 4 | 80.9 (80.8, 81.0) | 81.4 (81.3, 81.5) | 81.9 (81.8, 82.0) | 81.7 (81.6, 81.8) | 81.3 (81.2, 81.4) | <0.001 |
| Threonine |  |  |  |  |  |  |
| Model 1 | 80.2 (80.1, 80.3) | 81.5 (81.4, 81.5) | 81.7 (81.6, 81.8) | 81.7 (81.6, 81.8) | 82.0 (81.9, 82.0) | <0.001 |
| Model 2 | 80.6 (80.5, 80.6) | 81.5 (81.4, 81.6) | 81.7 (81.6, 81.8) | 81.6 (81.5, 81.7) | 81.8 (81.7, 81.9) | <0.001 |
| Model 3 | 80.5 (80.4, 80.6) | 81.4 (81.4, 81.5) | 81.6 (81.6, 81.7) | 81.6 (81.5, 81.7) | 81.9 (81.8, 82.0) | <0.001 |
| Model 4 | 80.8 (80.7, 80.9) | 81.5 (81.4, 81.5) | 81.6 (81.5, 81.7) | 81.5 (81.5, 81.6) | 81.8 (81.7, 81.9) | <0.001 |
| Tryptophan |  |  |  |  |  |  |
| Model 1 | 80.2 (80.1, 80.3) | 81.6 (81.5, 81.7) | 82.0 (81.9, 82.0) | 81.7 (81.6, 81.8) | 81.7 (81.6, 81.8) | <0.001 |
| Model 2 | 80.6 (80.5, 80.7) | 81.6 (81.5, 81.7) | 81.9 (81.8, 82.0) | 81.6 (81.6, 81.7) | 81.5 (81.4, 81.6) | <0.001 |
| Model 3 | 80.5 (80.4, 80.6) | 81.6 (81.5, 81.7) | 81.8 (81.8, 81.9) | 81.7 (81.6, 81.8) | 81.6 (81.5, 81.7) | <0.001 |
| Model 4 | 80.6 (80.5, 80.7) | 81.5 (81.4, 81.6) | 81.8 (81.7, 81.9) | 81.6 (81.6, 81.7) | 81.7 (81.6, 81.8) | <0.001 |
| Non-essential amino acids |  |  |  |  |  |  |
| Model 1 | 80.1 (79.9, 80.2) | 81.2 (81.1, 81.3) | 82.3 (82.2, 82.4) | 82.2 (82.1, 82.3) | 81.2 (81.2, 81.3) | <0.001 |
| Model 2 | 80.4 (80.3, 80.5) | 81.4 (81.3, 81.5) | 82.2 (82.1, 82.3) | 82.0 (81.9, 82.1) | 81.1 (81.0, 81.2) | <0.001 |
| Model 3 | 80.5 (80.4, 80.6) | 81.3 (81.2, 81.4) | 82.1 (82.0, 82.2) | 81.9 (81.8, 82.0) | 81.3 (81.2, 81.3) | <0.001 |
| Model 4 | 80.7 (80.6, 80.8) | 81.3 (81.2, 81.4) | 82.0 (81.9, 82.1) | 81.8 (81.8, 81.9) | 81.3 (81.2, 81.4) | <0.001 |
| Alanine |  |  |  |  |  |  |
| Model 1 | 79.7 (79.6, 79.8) | 81.2 (81.1, 81.3) | 81.7 (81.7, 81.8) | 82.1 (82.0, 82.2) | 82.2 (82.1, 82.3) | <0.001 |
| Model 2 | 80.1 (80.0, 80.2) | 81.3 (81.3, 81.4) | 81.7 (81.6, 81.8) | 81.9 (81.9, 82.0) | 81.9 (81.9, 82.0) | <0.001 |
| Model 3 | 80.2 (80.1, 80.3) | 81.3 (81.2, 81.4) | 81.7 (81.6, 81.7) | 81.9 (81.9, 82.0) | 82.0 (81.9, 82.1) | <0.001 |
| Model 4 | 80.5 (80.4, 80.6) | 81.4 (81.3, 81.4) | 81.6 (81.5, 81.7) | 81.8 (81.8, 81.9) | 81.8 (81.7, 81.9) | <0.001 |
| Arginine |  |  |  |  |  |  |
| Model 1 | 80.3 (80.2, 80.4) | 81.3 (81.3, 81.4) | 81.7 (81.7, 81.8) | 82.0 (81.9, 82.1) | 81.9 (81.8, 82.0) | <0.001 |
| Model 2 | 80.5 (80.4, 80.6) | 81.4 (81.4, 81.5) | 81.7 (81.6, 81.8) | 81.9 (81.8, 82.0) | 81.7 (81.6, 81.8) | <0.001 |
| Model 3 | 80.6 (80.5, 80.7) | 81.4 (81.3, 81.5) | 81.6 (81.6, 81.7) | 81.8 (81.8, 81.9) | 81.7 (81.6, 81.8) | <0.001 |
| Model 4 | 80.9 (80.8, 81.0) | 81.5 (81.4, 81.6) | 81.6 (81.5, 81.7) | 81.7 (81.7, 81.8) | 81.6 (81.5, 81.7) | <0.001 |
| Aspartic acid |  |  |  |  |  |  |
| Model 1 | 80.9 (80.9, 81.0) | 81.6 (81.5, 81.7) | 81.8 (81.7, 81.9) | 81.6 (81.5, 81.7) | 81.4 (81.3, 81.5) | <0.001 |
| Model 2 | 81.2 (81.1, 81.3) | 81.7 (81.6, 81.7) | 81.7 (81.7, 81.8) | 81.5 (81.4, 81.5) | 81.3 (81.2, 81.4) | <0.001 |
| Model 3 | 81.0 (81.0, 81.1) | 81.5 (81.4, 81.6) | 81.7 (81.6, 81.8) | 81.6 (81.5, 81.6) | 81.5 (81.4, 81.6) | <0.001 |
| Model 4 | 81.3 (81.2, 81.4) | 81.5 (81.4, 81.6) | 81.6 (81.6, 81.7) | 81.5 (81.4, 81.6) | 81.4 (81.3, 81.5) | <0.001 |
| Cystine |  |  |  |  |  |  |
| Model 1 | 80.0 (79.9, 80.1) | 81.2 (81.1, 81.3) | 82.1 (82.0, 82.2) | 82.2 (82.1, 82.3) | 81.5 (81.4, 81.6) | <0.001 |
| Model 2 | 80.4 (80.3, 80.5) | 81.3 (81.2, 81.4) | 82.0 (81.9, 82.1) | 82.0 (81.9, 82.1) | 81.4 (81.3, 81.5) | <0.001 |
| Model 3 | 80.6 (80.5, 80.7) | 81.4 (81.3, 81.5) | 81.9 (81.8, 82.0) | 81.9 (81.8, 82.0) | 81.4 (81.3, 81.5) | <0.001 |
| Model 4 | 80.8 (80.7, 80.9) | 81.3 (81.2, 81.4) | 81.8 (81.7, 81.9) | 81.8 (81.8, 81.9) | 81.4 (81.4, 81.5) | <0.001 |
| Glutamic acid |  |  |  |  |  |  |
| Model 1 | 80.2 (80.1, 80.3) | 81.1 (81.0, 81.2) | 82.3 (82.2, 82.4) | 82.3 (82.2, 82.3) | 81.1 (81.0, 81.2) | <0.001 |
| Model 2 | 80.5 (80.4, 80.6) | 81.3 (81.2, 81.4) | 82.2 (82.1, 82.3) | 82.1 (82.0, 82.2) | 81.0 (80.9, 81.1) | <0.001 |
| Model 3 | 80.7 (80.6, 80.8) | 81.3 (81.2, 81.4) | 82.0 (81.9, 82.1) | 82.0 (81.9, 82.1) | 81.1 (81.1, 81.2) | <0.001 |
| Model 4 | 80.7 (80.6, 80.8) | 81.3 (81.2, 81.3) | 81.9 (81.8, 82.0) | 81.9 (81.8, 82.0) | 81.3 (81.2, 81.4) | <0.001 |
| Glycine |  |  |  |  |  |  |
| Model 1 | 79.7 (79.6, 79.8) | 81.0 (80.9, 81.1) | 81.7 (81.6, 81.8) | 82.1 (82.0, 82.2) | 82.3 (82.2, 82.4) | <0.001 |
| Model 2 | 80.1 (80.0, 80.2) | 81.2 (81.1, 81.3) | 81.7 (81.6, 81.8) | 82.0 (81.9, 82.1) | 82.0 (82.0, 82.1) | <0.001 |
| Model 3 | 80.3 (80.2, 80.4) | 81.2 (81.1, 81.3) | 81.6 (81.5, 81.7) | 81.9 (81.8, 82.0) | 82.0 (81.9, 82.1) | <0.001 |
| Model 4 | 80.7 (80.6, 80.7) | 81.3 (81.2, 81.4) | 81.6 (81.5, 81.7) | 81.8 (81.7, 81.9) | 81.8 (81.7, 81.9) | <0.001 |
| Hydroxyproline |  |  |  |  |  |  |
| Model 1 | 80.0 (79.9, 80.1) | 81.5 (81.4, 81.6) | 81.7 (81.6, 81.8) | 81.8 (81.7, 81.9) | 81.7 (81.6, 81.8) | <0.001 |
| Model 2 | 80.1 (80.0, 80.2) | 81.4 (81.3, 81.5) | 81.6 (81.5, 81.7) | 81.8 (81.7, 81.9) | 81.8 (81.7, 81.9) | <0.001 |
| Model 3 | 80.4 (80.3, 80.5) | 81.5 (81.4, 81.6) | 81.6 (81.6, 81.7) | 81.7 (81.6, 81.8) | 81.6 (81.6, 81.7) | <0.001 |
| Model 4 | 80.9 (80.8, 81.0) | 81.6 (81.5, 81.7) | 81.6 (81.6, 81.7) | 81.6 (81.6, 81.7) | 81.4 (81.3, 81.5) | <0.001 |
| Proline |  |  |  |  |  |  |
| Model 1 | 80.5 (80.4, 80.6) | 81.3 (81.2, 81.4) | 81.9 (81.8, 82.0) | 82.0 (81.9, 82.1) | 81.3 (81.2, 81.4) | <0.001 |
| Model 2 | 80.8 (80.7, 80.9) | 81.4 (81.3, 81.5) | 81.9 (81.8, 82.0) | 81.9 (81.8, 81.9) | 81.2 (81.1, 81.3) | <0.001 |
| Model 3 | 80.9 (80.8, 81.0) | 81.4 (81.3, 81.5) | 81.8 (81.7, 81.9) | 81.8 (81.7, 81.9) | 81.3 (81.2, 81.4) | <0.001 |
| Model 4 | 81.0 (80.9, 81.1) | 81.4 (81.3, 81.4) | 81.7 (81.6, 81.8) | 81.7 (81.7, 81.8) | 81.4 (81.3, 81.5) | <0.001 |
| Serine |  |  |  |  |  |  |
| Model 1 | 80.4 (80.3, 80.5) | 81.4 (81.3, 81.5) | 82.1 (82.1, 82.2) | 82.0 (81.9, 82.0) | 81.2 (81.1, 81.2) | <0.001 |
| Model 2 | 80.7 (80.6, 80.8) | 81.5 (81.4, 81.6) | 82.1 (82.0, 82.2) | 81.8 (81.7, 81.9) | 81.1 (81.0, 81.2) | <0.001 |
| Model 3 | 80.7 (80.6, 80.8) | 81.4 (81.4, 81.5) | 81.9 (81.9, 82.0) | 81.8 (81.7, 81.8) | 81.3 (81.2, 81.3) | <0.001 |
| Model 4 | 80.9 (80.8, 81.0) | 81.4 (81.3, 81.5) | 81.9 (81.8, 81.9) | 81.7 (81.6, 81.7) | 81.3 (81.3, 81.4) | <0.001 |
| Tyrosine |  |  |  |  |  |  |
| Model 1 | 80.6 (80.5, 80.7) | 81.5 (81.4, 81.6) | 81.6 (81.5, 81.7) | 81.7 (81.6, 81.8) | 81.6 (81.5, 81.7) | <0.001 |
| Model 2 | 80.8 (80.7, 80.9) | 81.6 (81.5, 81.7) | 81.6 (81.5, 81.7) | 81.6 (81.5, 81.7) | 81.6 (81.5, 81.6) | <0.001 |
| Model 3 | 80.7 (80.6, 80.8) | 81.5 (81.4, 81.6) | 81.6 (81.5, 81.6) | 81.6 (81.6, 81.7) | 81.6 (81.6, 81.7) | <0.001 |
| Model 4 | 81.0 (80.9, 81.1) | 81.5 (81.4, 81.6) | 81.5 (81.4, 81.6) | 81.5 (81.5, 81.6) | 81.6 (81.6, 81.7) | <0.001 |
| Total protein (g/1000 kcal) |  |  |  |  |  |  |
| Model 1 | 81.1 (81.0, 81.2) | 81.3 (81.2, 81.4) | 81.5 (81.4, 81.6) | 81.6 (81.5, 81.7) | 81.9 (81.8, 82.0) | <0.001 |
| Model 2 | 80.9 (80.8, 80.9) | 81.2 (81.1, 81.3) | 81.5 (81.4, 81.6) | 81.7 (81.6, 81.8) | 82.1 (82.0, 82.2) | <0.001 |
| Model 3 | 81.1 (81.0, 81.2) | 81.3 (81.2, 81.3) | 81.5 (81.4, 81.6) | 81.6 (81.5, 81.7) | 81.9 (81.8, 82.0) | <0.001 |
| Model 4 | 81.4 (81.3, 81.5) | 81.4 (81.4, 81.5) | 81.5 (81.4, 81.6) | 81.5 (81.4, 81.6) | 81.5 (81.4, 81.6) | 0.40 |

Model 1 was adjusted for age (continuous), sex, centre
Model 2 was model 1 plus smoking (never, former, current<10, 10-19, 20+ cigarettes/day, unknown), current alcohol consumption (non-drinkers, 0.1-4.9, 5.0-14.9, 15-29.9, 30-59.9, 60+ g/day), physical activity (inactive, moderately inactive, moderately active, active, unknown), employment status (employed or student, neither employed nor student, unknown), highest level of education completed (none or primary, secondary, vocational or university, unknown),calibrated energy intake (continuous)
Model 3 was model 2 plus history of diabetes (yes, no, unknown), prior hypertension (yes, no , unknown), prior hyperlipidaemia (yes, no, unknown)
Model 4 was model 3 plus body mass index (<22.5, 22.5-24.9, 25.0-27.4, 27.5-29.9, ≥30.0 kg/m^2^, unknown)

Supplementary table 12: Hazard ratios (95% confidence intervals)^1^ for **ischaemic** (3603 cases) and **haemorrhagic** stroke (1146 cases) by increments of calibrated intakes of dietary amino acids, with **further adjustment for systolic and diastolic blood pressure**.

|  | HR (95% Cis) for per SD of calibrated intakes | | | | | | |
| --- | --- | --- | --- | --- | --- | --- | --- |
| Amino acids (percent of total protein) | Ischaemic stroke | | |  | Haemorrhagic stroke | | |
|  | Multivariable^1^ | Plus SBP | Plus DBP |  | Multivariable^1^ | Plus SBP | Plus DBP |
| Branched-chain amino acids | 0.92 (0.86, 0.98) | 0.92 (0.87, 0.98) | 0.92 (0.87, 0.98) |  | 0.99 (0.89, 1.10) | 0.99 (0.89, 1.11) | 0.99 (0.89, 1.10) |
| Isoleucine | 0.92 (0.86, 0.98) | 0.92 (0.86, 0.98) | 0.92 (0.86, 0.98) |  | 0.99 (0.89, 1.11) | 1.00 (0.89, 1.12) | 1.00 (0.89, 1.11) |
| Leucine | 0.92 (0.86, 0.98) | 0.92 (0.86, 0.98) | 0.92 (0.86, 0.98) |  | 0.99 (0.89, 1.11) | 1.00 (0.89, 1.11) | 0.99 (0.89, 1.11) |
| Valine | 0.92 (0.87, 0.97) | 0.92 (0.87, 0.97) | 0.92 (0.87, 0.97) |  | 0.98 (0.89, 1.09) | 0.99 (0.89, 1.10) | 0.99 (0.89, 1.09) |
| Other essential amino acids | 0.92 (0.86, 0.98) | 0.92 (0.87, 0.98) | 0.92 (0.86, 0.98) |  | 1.00 (0.90, 1.13) | 1.01 (0.90, 1.13) | 1.01 (0.90, 1.13) |
| Histidine | 0.93 (0.87, 0.99) | 0.93 (0.87, 0.99) | 0.93 (0.87, 0.99) |  | 1.00 (0.89, 1.13) | 1.00 (0.89, 1.13) | 1.00 (0.89, 1.13) |
| Lysine | 0.92 (0.86, 0.99) | 0.92 (0.86, 0.99) | 0.92 (0.86, 0.99) |  | 1.02 (0.90, 1.16) | 1.02 (0.90, 1.16) | 1.02 (0.90, 1.16) |
| Methionine | 0.92 (0.86, 0.99) | 0.92 (0.86, 0.99) | 0.92 (0.86, 0.99) |  | 1.02 (0.90, 1.15) | 1.02 (0.90, 1.16) | 1.02 (0.90, 1.15) |
| Phenylalanine | 0.92 (0.87, 0.98) | 0.92 (0.87, 0.98) | 0.92 (0.87, 0.98) |  | 0.98 (0.88, 1.10) | 0.99 (0.89, 1.10) | 0.99 (0.89, 1.10) |
| Threonine | 0.92 (0.86, 0.98) | 0.92 (0.87, 0.98) | 0.92 (0.86, 0.98) |  | 1.00 (0.90, 1.12) | 1.00 (0.90, 1.12) | 1.00 (0.90, 1.12) |
| Tryptophan | 0.93 (0.88, 0.99) | 0.93 (0.88, 0.99) | 0.93 (0.88, 0.99) |  | 1.01 (0.91, 1.12) | 1.01 (0.91, 1.12) | 1.01 (0.91, 1.12) |
| Non-essential amino acids | 0.92 (0.86, 0.98) | 0.92 (0.86, 0.98) | 0.92 (0.86, 0.98) |  | 0.99 (0.88, 1.11) | 1.00 (0.89, 1.12) | 0.99 (0.88, 1.12) |
| Alanine | 0.94 (0.87, 1.01) | 0.94 (0.87, 1.01) | 0.94 (0.87, 1.01) |  | 1.00 (0.88, 1.13) | 1.00 (0.88, 1.14) | 1.00 (0.88, 1.13) |
| Arginine | 0.96 (0.91, 1.01) | 0.96 (0.91, 1.01) | 0.96 (0.91, 1.01) |  | 0.97 (0.88, 1.08) | 0.98 (0.88, 1.08) | 0.97 (0.88, 1.08) |
| Aspartic acid | 0.95 (0.90, 0.99) | 0.95 (0.90, 0.99) | 0.95 (0.90, 0.99) |  | 1.02 (0.93, 1.11) | 1.02 (0.94, 1.11) | 1.02 (0.94, 1.11) |
| Cystine | 0.97 (0.90, 1.04) | 0.97 (0.90, 1.05) | 0.97 (0.90, 1.05) |  | 0.93 (0.81, 1.07) | 0.94 (0.81, 1.08) | 0.93 (0.81, 1.08) |
| Glutamic acid | 0.89 (0.83, 0.96) | 0.89 (0.83, 0.96) | 0.89 (0.83, 0.96) |  | 0.98 (0.86, 1.12) | 0.99 (0.87, 1.13) | 0.99 (0.87, 1.13) |
| Glycine | 0.97 (0.90, 1.05) | 0.97 (0.90, 1.04) | 0.97 (0.90, 1.04) |  | 1.00 (0.88, 1.14) | 1.00 (0.87, 1.13) | 1.00 (0.87, 1.13) |
| Hydroxyproline | 1.00 (0.93, 1.07) | 0.99 (0.92, 1.06) | 0.99 (0.92, 1.07) |  | 1.17 (1.04, 1.32) | 1.16 (1.03, 1.31) | 1.16 (1.03, 1.31) |
| Proline | 0.88 (0.82, 0.94) | 0.88 (0.82, 0.94) | 0.88 (0.82, 0.94) |  | 0.96 (0.85, 1.09) | 0.97 (0.86, 1.10) | 0.97 (0.85, 1.09) |
| Serine | 0.92 (0.87, 0.98) | 0.92 (0.87, 0.98) | 0.92 (0.87, 0.98) |  | 1.01 (0.91, 1.12) | 1.01 (0.91, 1.12) | 1.01 (0.91, 1.12) |
| Tyrosine | 0.92 (0.87, 0.98) | 0.92 (0.87, 0.98) | 0.92 (0.87, 0.98) |  | 1.01 (0.91, 1.12) | 1.01 (0.91, 1.13) | 1.01 (0.91, 1.12) |
| Total protein (g/1000 kcal) | 0.97 (0.94, 1.00) | 0.97 (0.94, 1.00) | 0.97 (0.94, 1.00) |  | 1.00 (0.95, 1.06) | 1.00 (0.95, 1.06) | 1.00 (0.95, 1.06) |

1. Restricted to participants with measures of systolic and diastolic blood pressure (267,642 participants, 3,603 cases of ischaemic stroke and 1,146 cases of haemorrhagic stroke). Hazard ratios modelled per 1 sex-specific SD increment in dietary amino acids or total protein. Based on multivariable model stratified by sex and centre, and adjusted for age (continuous), smoking (never, former, current<10, 10-19, 20+ cigarettes/day, unknown), calibrated alcohol consumption (non-drinkers (<0.1), 0.1-4.9, 5.0-14.9, 15-29.9, 30-59.9, 60+ g/day), physical activity (inactive, moderately inactive, moderately active, active, unknown), employment status (employed or student, neither employed nor student, unknown), highest level of education completed (none or primary, secondary, vocational or university, unknown), history of diabetes (yes, no, unknown), prior hypertension (yes, no , unknown), prior hyperlipidaemia (yes, no, unknown), calibrated energy intake (continuous), body mass index (<22.5, 22.5-24.9, 25.0-27.4, 27.5-29.9, ≥30.0 kg/m^2^, unknown).

Supplementary table 13: Hazard ratios (95% confidence intervals)^1^ for **ischaemic** stroke by increments of calibrated intakes of dietary amino acids, stratified by **age at recruitment**

| Amino acids (percent of total protein) | <55 years  (1192 cases) | |  | 55-64 years  (2209 cases) | |  | ≥65 years  (894 cases) | |  |  |
| --- | --- | --- | --- | --- | --- | --- | --- | --- | --- | --- |
|  | HR (95% CI) | P for trend^2^ |  | HR (95% CI) | P for trend^2^ |  | HR (95% CI) | P for trend^2^ |  | P-het^3^ |
| Branched-chain amino acids | 0.93 (0.84, 1.03) | 0.19 |  | 0.89 (0.82, 0.96) | 0.002 |  | 1.04 (0.91, 1.18) | 0.58 |  | 0.13 |
| Isoleucine | 0.94 (0.85, 1.04) | 0.22 |  | 0.89 (0.82, 0.96) | 0.003 |  | 1.04 (0.91, 1.18) | 0.57 |  | 0.13 |
| Leucine | 0.93 (0.84, 1.04) | 0.20 |  | 0.89 (0.82, 0.96) | 0.003 |  | 1.04 (0.92, 1.19) | 0.52 |  | 0.11 |
| Valine | 0.93 (0.84, 1.02) | 0.14 |  | 0.89 (0.83, 0.96) | 0.002 |  | 1.03 (0.91, 1.16) | 0.68 |  | 0.15 |
| Other essential amino acids | 0.95 (0.85, 1.06) | 0.34 |  | 0.89 (0.82, 0.97) | 0.006 |  | 1.04 (0.91, 1.19) | 0.56 |  | 0.15 |
| Histidine | 0.96 (0.86, 1.07) | 0.47 |  | 0.90 (0.83, 0.98) | 0.01 |  | 1.04 (0.91, 1.19) | 0.59 |  | 0.20 |
| Lysine | 0.95 (0.85, 1.07) | 0.38 |  | 0.90 (0.82, 0.98) | 0.02 |  | 1.04 (0.91, 1.20) | 0.55 |  | 0.19 |
| Methionine | 0.95 (0.84, 1.07) | 0.39 |  | 0.89 (0.81, 0.97) | 0.01 |  | 1.04 (0.91, 1.20) | 0.55 |  | 0.16 |
| Phenylalanine | 0.94 (0.84, 1.03) | 0.19 |  | 0.89 (0.82, 0.96) | 0.002 |  | 1.04 (0.91, 1.18) | 0.57 |  | 0.12 |
| Threonine | 0.95 (0.86, 1.06) | 0.36 |  | 0.89 (0.82, 0.96) | 0.003 |  | 1.04 (0.91, 1.18) | 0.58 |  | 0.12 |
| Tryptophan | 0.96 (0.88, 1.06) | 0.46 |  | 0.89 (0.83, 0.96) | 0.002 |  | 1.05 (0.93, 1.19) | 0.43 |  | 0.06 |
| Non-essential amino acids | 0.95 (0.85, 1.05) | 0.31 |  | 0.88 (0.81, 0.95) | 0.001 |  | 1.04 (0.91, 1.20) | 0.54 |  | 0.09 |
| Alanine | 1.00 (0.89, 1.12) | 0.99 |  | 0.92 (0.84, 1.00) | 0.06 |  | 1.05 (0.91, 1.21) | 0.50 |  | 0.23 |
| Arginine | 1.01 (0.92, 1.10) | 0.90 |  | 0.93 (0.87, 1.00) | 0.04 |  | 1.04 (0.93, 1.17) | 0.46 |  | 0.16 |
| Aspartic acid | 0.99 (0.92, 1.07) | 0.87 |  | 0.91 (0.86, 0.97) | 0.003 |  | 1.04 (0.95, 1.15) | 0.39 |  | 0.05 |
| Cystine | 1.05 (0.92, 1.19) | 0.48 |  | 0.92 (0.84, 1.02) | 0.11 |  | 1.01 (0.85, 1.20) | 0.92 |  | 0.28 |
| Glutamic acid | 0.92 (0.81, 1.04) | 0.17 |  | 0.85 (0.77, 0.93) | 0.001 |  | 1.04 (0.88, 1.23) | 0.62 |  | 0.10 |
| Glycine | 1.03 (0.92, 1.16) | 0.55 |  | 0.95 (0.87, 1.04) | 0.28 |  | 1.06 (0.92, 1.23) | 0.41 |  | 0.33 |
| Hydroxyproline | 1.02 (0.89, 1.16) | 0.80 |  | 0.99 (0.90, 1.10) | 0.92 |  | 1.01 (0.86, 1.18) | 0.91 |  | 0.96 |
| Proline | 0.86 (0.76, 0.97) | 0.02 |  | 0.85 (0.77, 0.93) | <0.001 |  | 1.03 (0.88, 1.21) | 0.70 |  | 0.10 |
| Serine | 0.94 (0.85, 1.03) | 0.19 |  | 0.89 (0.82, 0.95) | 0.001 |  | 1.04 (0.92, 1.18) | 0.55 |  | 0.09 |
| Tyrosine | 0.93 (0.84, 1.02) | 0.14 |  | 0.89 (0.82, 0.96) | 0.002 |  | 1.04 (0.92, 1.18) | 0.55 |  | 0.11 |
| Total protein (g/1000 kcal) | 0.98 (0.93, 1.03) | 0.39 |  | 0.97 (0.93, 1.00) | 0.08 |  | 0.99 (0.93, 1.06) | 0.75 |  | 0.81 |

1. Hazard ratios modelled per 1 sex-specific SD increment in dietary amino acids or total protein. Based on multivariable model stratified by sex and centre, and adjusted for age (continuous), smoking (never, former, current<10, 10-19, 20+ cigarettes/day, unknown), calibrated alcohol consumption (non-drinkers (<0.1), 0.1-4.9, 5.0-14.9, 15-29.9, 30-59.9, 60+ g/day), physical activity (inactive, moderately inactive, moderately active, active, unknown), employment status (employed or student, neither employed nor student, unknown), highest level of education completed (none or primary, secondary, vocational or university, unknown), history of diabetes (yes, no, unknown), prior hypertension (yes, no , unknown), prior hyperlipidaemia (yes, no, unknown), calibrated energy intake (continuous), body mass index (<22.5, 22.5-24.9, 25.0-27.4, 27.5-29.9, ≥30.0 kg/m^2^, unknown).
2. Tests of trend were performed using the calibrated intake (continuous).
3. Tests of heterogeneity of trend by age at recruitment were obtained assuming independence of risk by age at recruitment using a meta-analysis method.

Supplementary table 14: Hazard ratios (95% confidence intervals)^1^ for **haemorrhagic** stroke by increments of calibrated intakes of dietary amino acids, stratified by **age at recruitment**

| Amino acids (percent of total protein) | <55 years  (516 cases) | |  | 55-64 years  (625 cases) | |  | ≥65 years  (234 cases) | |  |  |
| --- | --- | --- | --- | --- | --- | --- | --- | --- | --- | --- |
|  | HR (95% CI) | P for trend^2^ |  | HR (95% CI) | P for trend^2^ |  | HR (95% CI) | P for trend^2^ |  | P-het^3^ |
| Branched-chain amino acids | 0.93 (0.79, 1.09) | 0.38 |  | 1.00 (0.87, 1.16) | 0.98 |  | 0.98 (0.76, 1.26) | 0.87 |  | 0.80 |
| Isoleucine | 0.92 (0.78, 1.09) | 0.36 |  | 1.00 (0.86, 1.16) | 0.98 |  | 0.98 (0.76, 1.27) | 0.90 |  | 0.77 |
| Leucine | 0.93 (0.79, 1.10) | 0.40 |  | 1.00 (0.86, 1.16) | 0.99 |  | 0.99 (0.76, 1.28) | 0.91 |  | 0.81 |
| Valine | 0.93 (0.80, 1.09) | 0.39 |  | 1.00 (0.87, 1.15) | 0.97 |  | 0.96 (0.76, 1.23) | 0.77 |  | 0.80 |
| Other essential amino acids | 0.93 (0.79, 1.11) | 0.43 |  | 1.01 (0.86, 1.18) | 0.91 |  | 0.98 (0.76, 1.28) | 0.90 |  | 0.80 |
| Histidine | 0.93 (0.78, 1.11) | 0.43 |  | 0.99 (0.84, 1.16) | 0.89 |  | 0.99 (0.75, 1.29) | 0.92 |  | 0.88 |
| Lysine | 0.95 (0.79, 1.14) | 0.56 |  | 1.02 (0.87, 1.21) | 0.78 |  | 0.97 (0.74, 1.27) | 0.82 |  | 0.82 |
| Methionine | 0.94 (0.78, 1.14) | 0.53 |  | 1.02 (0.86, 1.20) | 0.85 |  | 0.97 (0.74, 1.27) | 0.81 |  | 0.83 |
| Phenylalanine | 0.93 (0.79, 1.09) | 0.37 |  | 1.00 (0.86, 1.16) | 0.99 |  | 1.00 (0.76, 1.30) | 0.98 |  | 0.78 |
| Threonine | 0.93 (0.79, 1.09) | 0.35 |  | 1.00 (0.87, 1.16) | 0.98 |  | 0.98 (0.76, 1.26) | 0.88 |  | 0.77 |
| Tryptophan | 0.94 (0.81, 1.10) | 0.45 |  | 1.01 (0.88, 1.16) | 0.85 |  | 1.00 (0.78, 1.29) | 0.97 |  | 0.78 |
| Non-essential amino acids | 0.94 (0.79, 1.12) | 0.47 |  | 1.00 (0.85, 1.16) | 0.95 |  | 1.01 (0.76, 1.33) | 0.96 |  | 0.85 |
| Alanine | 0.90 (0.75, 1.08) | 0.27 |  | 0.97 (0.81, 1.15) | 0.69 |  | 1.01 (0.76, 1.34) | 0.96 |  | 0.78 |
| Arginine | 0.90 (0.78, 1.04) | 0.16 |  | 0.95 (0.83, 1.08) | 0.41 |  | 1.06 (0.85, 1.33) | 0.61 |  | 0.49 |
| Aspartic acid | 0.97 (0.85, 1.10) | 0.61 |  | 1.02 (0.91, 1.14) | 0.78 |  | 1.03 (0.84, 1.26) | 0.78 |  | 0.81 |
| Cystine | 0.83 (0.67, 1.04) | 0.10 |  | 0.96 (0.79, 1.16) | 0.68 |  | 0.99 (0.69, 1.42) | 0.95 |  | 0.57 |
| Glutamic acid | 0.97 (0.80, 1.19) | 0.80 |  | 0.99 (0.83, 1.18) | 0.88 |  | 0.98 (0.70, 1.38) | 0.91 |  | 1.00 |
| Glycine | 0.91 (0.75, 1.09) | 0.29 |  | 0.95 (0.79, 1.13) | 0.53 |  | 1.03 (0.76, 1.38) | 0.86 |  | 0.78 |
| Hydroxyproline | 1.23 (1.03, 1.47) | 0.02 |  | 1.07 (0.90, 1.27) | 0.46 |  | 1.15 (0.89, 1.48) | 0.29 |  | 0.53 |
| Proline | 0.95 (0.79, 1.15) | 0.61 |  | 0.99 (0.84, 1.17) | 0.89 |  | 0.92 (0.67, 1.28) | 0.63 |  | 0.92 |
| Serine | 0.95 (0.81, 1.11) | 0.50 |  | 1.03 (0.90, 1.18) | 0.69 |  | 1.02 (0.80, 1.32) | 0.86 |  | 0.73 |
| Tyrosine | 0.96 (0.82, 1.12) | 0.59 |  | 1.03 (0.90, 1.19) | 0.68 |  | 0.98 (0.76, 1.25) | 0.87 |  | 0.79 |
| Total protein (g/1000 kcal) | 0.96 (0.88, 1.04) | 0.31 |  | 0.99 (0.92, 1.07) | 0.85 |  | 1.05 (0.92, 1.19) | 0.50 |  | 0.53 |

1. Hazard ratios modelled per 1 sex-specific SD increment in dietary amino acids or total protein. Based on multivariable model stratified by sex and centre, and adjusted for age (continuous), smoking (never, former, current<10, 10-19, 20+ cigarettes/day, unknown), calibrated alcohol consumption (non-drinkers (<0.1), 0.1-4.9, 5.0-14.9, 15-29.9, 30-59.9, 60+ g/day), physical activity (inactive, moderately inactive, moderately active, active, unknown), employment status (employed or student, neither employed nor student, unknown), highest level of education completed (none or primary, secondary, vocational or university, unknown), history of diabetes (yes, no, unknown), prior hypertension (yes, no , unknown), prior hyperlipidaemia (yes, no, unknown), calibrated energy intake (continuous), body mass index (<22.5, 22.5-24.9, 25.0-27.4, 27.5-29.9, ≥30.0 kg/m^2^, unknown).
2. Tests of trend were performed using the calibrated intake (continuous).
3. Tests of heterogeneity of trend by age at recruitment were obtained assuming independence of risk by age at recruitment using a meta-analysis method.

Supplementary table 15: Hazard ratios (95% confidence intervals)^1^ for **ischaemic** stroke by increments of calibrated intakes of dietary amino acids, stratified by **sex**

| Amino acids (percent of total protein) | Men  (2235 cases) | |  | Women  (2060 cases) | |  |  |  |
| --- | --- | --- | --- | --- | --- | --- | --- | --- |
|  | HR (95% CI) | P for trend^2^ |  | HR (95% CI) | P for trend^2^ |  |  | P-het^3^ |
| Branched-chain amino acids | 0.93 (0.87, 1.00) | 0.04 |  | 0.92 (0.84, 1.01) | 0.08 |  |  | 0.84 |
| Isoleucine | 0.93 (0.87, 1.00) | 0.05 |  | 0.93 (0.84, 1.02) | 0.10 |  |  | 0.89 |
| Leucine | 0.93 (0.87, 1.00) | 0.05 |  | 0.92 (0.84, 1.01) | 0.09 |  |  | 0.86 |
| Valine | 0.93 (0.87, 1.00) | 0.04 |  | 0.92 (0.84, 1.00) | 0.05 |  |  | 0.78 |
| Other essential amino acids | 0.94 (0.87, 1.01) | 0.09 |  | 0.93 (0.84, 1.03) | 0.15 |  |  | 0.88 |
| Histidine | 0.94 (0.88, 1.02) | 0.13 |  | 0.94 (0.85, 1.04) | 0.23 |  |  | 0.94 |
| Lysine | 0.94 (0.87, 1.02) | 0.14 |  | 0.93 (0.83, 1.04) | 0.19 |  |  | 0.82 |
| Methionine | 0.94 (0.87, 1.02) | 0.14 |  | 0.92 (0.83, 1.03) | 0.15 |  |  | 0.78 |
| Phenylalanine | 0.93 (0.87, 1.00) | 0.04 |  | 0.92 (0.84, 1.01) | 0.08 |  |  | 0.86 |
| Threonine | 0.94 (0.87, 1.00) | 0.06 |  | 0.93 (0.85, 1.02) | 0.14 |  |  | 0.97 |
| Tryptophan | 0.93 (0.87, 0.99) | 0.03 |  | 0.95 (0.88, 1.04) | 0.29 |  |  | 0.62 |
| Non-essential amino acids | 0.93 (0.86, 1.00) | 0.06 |  | 0.92 (0.84, 1.01) | 0.09 |  |  | 0.87 |
| Alanine | 0.97 (0.89, 1.05) | 0.41 |  | 0.96 (0.87, 1.07) | 0.50 |  |  | 0.97 |
| Arginine | 0.97 (0.92, 1.03) | 0.37 |  | 0.97 (0.89, 1.05) | 0.48 |  |  | 0.97 |
| Aspartic acid | 0.95 (0.91, 1.01) | 0.09 |  | 0.97 (0.90, 1.04) | 0.42 |  |  | 0.71 |
| Cystine | 0.97 (0.89, 1.06) | 0.55 |  | 0.97 (0.86, 1.09) | 0.61 |  |  | 0.96 |
| Glutamic acid | 0.90 (0.83, 0.99) | 0.03 |  | 0.89 (0.80, 1.00) | 0.04 |  |  | 0.86 |
| Glycine | 1.00 (0.92, 1.08) | 0.98 |  | 0.99 (0.90, 1.10) | 0.85 |  |  | 0.90 |
| Hydroxyproline | 1.02 (0.93, 1.12) | 0.65 |  | 0.98 (0.88, 1.10) | 0.79 |  |  | 0.62 |
| Proline | 0.90 (0.82, 0.98) | 0.01 |  | 0.86 (0.78, 0.96) | 0.005 |  |  | 0.57 |
| Serine | 0.93 (0.87, 0.99) | 0.03 |  | 0.92 (0.84, 1.00) | 0.06 |  |  | 0.84 |
| Tyrosine | 0.93 (0.87, 1.00) | 0.04 |  | 0.91 (0.84, 1.00) | 0.06 |  |  | 0.74 |
| Total protein (g/1000 kcal) | 0.98 (0.94, 1.01) | 0.16 |  | 0.97 (0.92, 1.01) | 0.17 |  |  | 0.77 |

1. Hazard ratios modelled per 1 sex-specific SD increment in dietary amino acids or total protein. Based on multivariable model stratified by sex and centre, and adjusted for age (continuous), smoking (never, former, current<10, 10-19, 20+ cigarettes/day, unknown), calibrated alcohol consumption (non-drinkers (<0.1), 0.1-4.9, 5.0-14.9, 15-29.9, 30-59.9, 60+ g/day), physical activity (inactive, moderately inactive, moderately active, active, unknown), employment status (employed or student, neither employed nor student, unknown), highest level of education completed (none or primary, secondary, vocational or university, unknown), history of diabetes (yes, no, unknown), prior hypertension (yes, no , unknown), prior hyperlipidaemia (yes, no, unknown), calibrated energy intake (continuous), body mass index (<22.5, 22.5-24.9, 25.0-27.4, 27.5-29.9, ≥30.0 kg/m^2^, unknown).
2. Tests of trend were performed using the calibrated intake (continuous).
3. Tests of heterogeneity of trend by sex were obtained assuming independence of risk by age at recruitment using a meta-analysis method.

Supplementary table 16: Hazard ratios (95% confidence intervals)^1^ for **haemorrhagic** stroke by increments of calibrated intakes of dietary amino acids, stratified by **sex**

| Amino acids (percent of total protein) | Men  (565 cases) | |  | Women  (810 cases) | |  |  |  |
| --- | --- | --- | --- | --- | --- | --- | --- | --- |
|  | HR (95% CI) | P for trend^2^ |  | HR (95% CI) | P for trend^2^ |  |  | P-het^3^ |
| Branched-chain amino acids | 0.95 (0.83, 1.09) | 0.46 |  | 0.98 (0.85, 1.13) | 0.78 |  |  | 0.76 |
| Isoleucine | 0.94 (0.82, 1.08) | 0.41 |  | 0.98 (0.85, 1.14) | 0.84 |  |  | 0.68 |
| Leucine | 0.95 (0.83, 1.09) | 0.48 |  | 0.98 (0.85, 1.14) | 0.80 |  |  | 0.76 |
| Valine | 0.95 (0.83, 1.09) | 0.46 |  | 0.97 (0.85, 1.12) | 0.70 |  |  | 0.82 |
| Other essential amino acids | 0.95 (0.82, 1.10) | 0.47 |  | 0.99 (0.85, 1.16) | 0.93 |  |  | 0.67 |
| Histidine | 0.92 (0.80, 1.08) | 0.31 |  | 0.99 (0.85, 1.17) | 0.94 |  |  | 0.52 |
| Lysine | 0.96 (0.83, 1.12) | 0.61 |  | 1.00 (0.85, 1.18) | 0.99 |  |  | 0.73 |
| Methionine | 0.96 (0.82, 1.12) | 0.62 |  | 0.99 (0.84, 1.16) | 0.87 |  |  | 0.82 |
| Phenylalanine | 0.95 (0.82, 1.09) | 0.45 |  | 0.98 (0.85, 1.13) | 0.79 |  |  | 0.74 |
| Threonine | 0.93 (0.81, 1.07) | 0.33 |  | 1.00 (0.86, 1.15) | 0.96 |  |  | 0.53 |
| Tryptophan | 0.96 (0.84, 1.10) | 0.53 |  | 0.99 (0.86, 1.14) | 0.90 |  |  | 0.73 |
| Non-essential amino acids | 0.94 (0.81, 1.10) | 0.45 |  | 0.98 (0.85, 1.15) | 0.84 |  |  | 0.70 |
| Alanine | 0.89 (0.76, 1.05) | 0.16 |  | 0.99 (0.84, 1.17) | 0.94 |  |  | 0.36 |
| Arginine | 0.91 (0.80, 1.03) | 0.13 |  | 0.98 (0.86, 1.12) | 0.81 |  |  | 0.38 |
| Aspartic acid | 0.98 (0.88, 1.09) | 0.65 |  | 1.02 (0.90, 1.14) | 0.79 |  |  | 0.61 |
| Cystine | 0.86 (0.71, 1.03) | 0.10 |  | 0.98 (0.81, 1.19) | 0.84 |  |  | 0.33 |
| Glutamic acid | 0.95 (0.79, 1.13) | 0.56 |  | 0.98 (0.82, 1.17) | 0.81 |  |  | 0.81 |
| Glycine | 0.87 (0.73, 1.03) | 0.10 |  | 1.00 (0.85, 1.18) | 1.00 |  |  | 0.24 |
| Hydroxyproline | 1.01 (0.85, 1.19) | 0.93 |  | 1.29 (1.10, 1.51) | 0.002 |  |  | 0.04 |
| Proline | 0.97 (0.82, 1.15) | 0.74 |  | 0.92 (0.79, 1.08) | 0.32 |  |  | 0.67 |
| Serine | 0.98 (0.86, 1.13) | 0.82 |  | 1.00 (0.87, 1.14) | 0.95 |  |  | 0.91 |
| Tyrosine | 0.99 (0.87, 1.13) | 0.88 |  | 0.98 (0.85, 1.13) | 0.83 |  |  | 0.95 |
| Total protein (g/1000 kcal) | 0.96 (0.90, 1.03) | 0.29 |  | 1.01 (0.94, 1.09) | 0.73 |  |  | 0.33 |

1. Hazard ratios modelled per 1 sex-specific SD increment in dietary amino acids or total protein. Based on multivariable model stratified by sex and centre, and adjusted for age (continuous), smoking (never, former, current<10, 10-19, 20+ cigarettes/day, unknown), calibrated alcohol consumption (non-drinkers (<0.1), 0.1-4.9, 5.0-14.9, 15-29.9, 30-59.9, 60+ g/day), physical activity (inactive, moderately inactive, moderately active, active, unknown), employment status (employed or student, neither employed nor student, unknown), highest level of education completed (none or primary, secondary, vocational or university, unknown), history of diabetes (yes, no, unknown), prior hypertension (yes, no , unknown), prior hyperlipidaemia (yes, no, unknown), calibrated energy intake (continuous), body mass index (<22.5, 22.5-24.9, 25.0-27.4, 27.5-29.9, ≥30.0 kg/m^2^, unknown).
2. Tests of trend were performed using the calibrated intake (continuous).
3. Tests of heterogeneity of trend by sex were obtained assuming independence of risk by age at recruitment using a meta-analysis method.

Supplementary table 17: Hazard ratios (95% confidence intervals)^1^ for **ischaemic** stroke by increments of calibrated intakes of dietary amino acids, stratified by **body mass index**

| Amino acids (percent of total protein) | BMI <25 kg/m^2^  (1479 cases) | |  | BMI 25-29.9 kg/m^2^  (1947 cases) | |  | BMI ≥30 kg/m^2^  (853 cases) | |  |  |
| --- | --- | --- | --- | --- | --- | --- | --- | --- | --- | --- |
|  | HR (95% CI) | P for trend^2^ |  | HR (95% CI) | P for trend^2^ |  | HR (95% CI) | P for trend^2^ |  | P-het^3^ |
| Branched-chain amino acids | 0.94 (0.85, 1.03) | 0.19 |  | 0.88 (0.82, 0.96) | 0.003 |  | 1.00 (0.89, 1.13) | 0.98 |  | 0.22 |
| Isoleucine | 0.94 (0.85, 1.04) | 0.20 |  | 0.89 (0.82, 0.96) | 0.003 |  | 1.00 (0.89, 1.13) | 0.95 |  | 0.24 |
| Leucine | 0.94 (0.85, 1.04) | 0.20 |  | 0.88 (0.81, 0.96) | 0.003 |  | 1.00 (0.89, 1.13) | 0.98 |  | 0.24 |
| Valine | 0.94 (0.85, 1.03) | 0.18 |  | 0.88 (0.82, 0.95) | 0.001 |  | 1.00 (0.89, 1.12) | 0.99 |  | 0.20 |
| Other essential amino acids | 0.95 (0.85, 1.05) | 0.31 |  | 0.89 (0.81, 0.97) | 0.006 |  | 1.01 (0.89, 1.15) | 0.82 |  | 0.21 |
| Histidine | 0.97 (0.87, 1.08) | 0.56 |  | 0.89 (0.82, 0.97) | 0.01 |  | 1.01 (0.88, 1.15) | 0.94 |  | 0.25 |
| Lysine | 0.95 (0.85, 1.06) | 0.35 |  | 0.89 (0.81, 0.98) | 0.01 |  | 1.02 (0.89, 1.17) | 0.75 |  | 0.25 |
| Methionine | 0.94 (0.84, 1.05) | 0.28 |  | 0.89 (0.81, 0.97) | 0.01 |  | 1.03 (0.90, 1.18) | 0.67 |  | 0.20 |
| Phenylalanine | 0.94 (0.85, 1.04) | 0.22 |  | 0.88 (0.81, 0.95) | 0.002 |  | 1.00 (0.89, 1.13) | 0.98 |  | 0.19 |
| Threonine | 0.94 (0.85, 1.04) | 0.26 |  | 0.89 (0.82, 0.96) | 0.004 |  | 1.01 (0.90, 1.14) | 0.83 |  | 0.19 |
| Tryptophan | 0.96 (0.87, 1.05) | 0.34 |  | 0.89 (0.82, 0.96) | 0.003 |  | 1.01 (0.90, 1.13) | 0.89 |  | 0.19 |
| Non-essential amino acids | 0.94 (0.85, 1.04) | 0.24 |  | 0.87 (0.80, 0.95) | 0.002 |  | 1.01 (0.89, 1.15) | 0.88 |  | 0.17 |
| Alanine | 0.97 (0.87, 1.09) | 0.67 |  | 0.91 (0.83, 0.99) | 0.04 |  | 1.06 (0.93, 1.21) | 0.37 |  | 0.15 |
| Arginine | 1.00 (0.91, 1.09) | 0.95 |  | 0.92 (0.86, 0.99) | 0.02 |  | 1.03 (0.93, 1.15) | 0.52 |  | 0.14 |
| Aspartic acid | 0.97 (0.90, 1.05) | 0.46 |  | 0.92 (0.86, 0.98) | 0.006 |  | 1.03 (0.94, 1.13) | 0.56 |  | 0.12 |
| Cystine | 1.00 (0.88, 1.14) | 0.97 |  | 0.89 (0.81, 0.99) | 0.03 |  | 1.10 (0.94, 1.28) | 0.24 |  | 0.08 |
| Glutamic acid | 0.91 (0.81, 1.03) | 0.13 |  | 0.85 (0.77, 0.94) | 0.001 |  | 0.99 (0.85, 1.15) | 0.91 |  | 0.23 |
| Glycine | 1.01 (0.90, 1.13) | 0.87 |  | 0.93 (0.85, 1.03) | 0.15 |  | 1.09 (0.95, 1.24) | 0.21 |  | 0.17 |
| Hydroxyproline | 0.96 (0.85, 1.09) | 0.55 |  | 1.00 (0.90, 1.11) | 0.96 |  | 1.10 (0.94, 1.29) | 0.24 |  | 0.42 |
| Proline | 0.89 (0.80, 1.00) | 0.05 |  | 0.85 (0.77, 0.93) | 0.001 |  | 0.94 (0.81, 1.08) | 0.38 |  | 0.50 |
| Serine | 0.94 (0.85, 1.03) | 0.18 |  | 0.88 (0.81, 0.95) | 0.001 |  | 1.01 (0.90, 1.13) | 0.91 |  | 0.16 |
| Tyrosine | 0.93 (0.85, 1.03) | 0.15 |  | 0.89 (0.82, 0.96) | 0.003 |  | 1.00 (0.89, 1.13) | 0.99 |  | 0.24 |
| Total protein (g/1000 kcal) | 0.99 (0.94, 1.03) | 0.56 |  | 0.95 (0.91, 0.99) | 0.007 |  | 1.01 (0.95, 1.07) | 0.69 |  | 0.15 |

1. Hazard ratios modelled per 1 sex-specific SD increment in dietary amino acids or total protein. Based on multivariable model stratified by sex and centre, and adjusted for age (continuous), smoking (never, former, current<10, 10-19, 20+ cigarettes/day, unknown), calibrated alcohol consumption (non-drinkers (<0.1), 0.1-4.9, 5.0-14.9, 15-29.9, 30-59.9, 60+ g/day), physical activity (inactive, moderately inactive, moderately active, active, unknown), employment status (employed or student, neither employed nor student, unknown), highest level of education completed (none or primary, secondary, vocational or university, unknown), history of diabetes (yes, no, unknown), prior hypertension (yes, no , unknown), prior hyperlipidaemia (yes, no, unknown), calibrated energy intake (continuous), body mass index (<22.5, 22.5-24.9, 25.0-27.4, 27.5-29.9, ≥30.0 kg/m^2^, unknown).
2. Tests of trend were performed using the calibrated intake (continuous).
3. Tests of heterogeneity of trend by body mass index were obtained assuming independence of risk by age at recruitment using a meta-analysis method.

Supplementary table 18: Hazard ratios (95% confidence intervals)^1^ for **haemorrhagic** stroke by increments of calibrated intakes of dietary amino acids, stratified by **body mass index**

| Amino acids (percent of total protein) | BMI <25 kg/m^2^  (583 cases) | |  | BMI 25-29.9 kg/m^2^  (560 cases) | |  | BMI ≥30 kg/m^2^  (226 cases) | |  |  |
| --- | --- | --- | --- | --- | --- | --- | --- | --- | --- | --- |
|  | HR (95% CI) | P for trend^2^ |  | HR (95% CI) | P for trend^2^ |  | HR (95% CI) | P for trend^2^ |  | P-het^3^ |
| Branched-chain amino acids | 0.93 (0.80, 1.09) | 0.39 |  | 1.03 (0.88, 1.20) | 0.73 |  | 0.94 (0.74, 1.19) | 0.61 |  | 0.66 |
| Isoleucine | 0.94 (0.80, 1.11) | 0.45 |  | 1.02 (0.88, 1.19) | 0.77 |  | 0.93 (0.73, 1.18) | 0.55 |  | 0.69 |
| Leucine | 0.93 (0.79, 1.09) | 0.38 |  | 1.03 (0.88, 1.20) | 0.71 |  | 0.95 (0.74, 1.20) | 0.66 |  | 0.65 |
| Valine | 0.93 (0.80, 1.09) | 0.37 |  | 1.03 (0.89, 1.19) | 0.74 |  | 0.94 (0.75, 1.18) | 0.60 |  | 0.65 |
| Other essential amino acids | 0.96 (0.81, 1.14) | 0.65 |  | 1.02 (0.87, 1.20) | 0.83 |  | 0.92 (0.72, 1.18) | 0.53 |  | 0.79 |
| Histidine | 0.96 (0.81, 1.15) | 0.66 |  | 0.99 (0.83, 1.17) | 0.87 |  | 0.93 (0.72, 1.20) | 0.58 |  | 0.93 |
| Lysine | 0.99 (0.83, 1.18) | 0.91 |  | 1.02 (0.86, 1.21) | 0.83 |  | 0.90 (0.69, 1.19) | 0.46 |  | 0.76 |
| Methionine | 0.95 (0.79, 1.14) | 0.58 |  | 1.03 (0.87, 1.22) | 0.74 |  | 0.94 (0.72, 1.24) | 0.68 |  | 0.78 |
| Phenylalanine | 0.93 (0.79, 1.09) | 0.35 |  | 1.03 (0.88, 1.21) | 0.68 |  | 0.95 (0.75, 1.21) | 0.68 |  | 0.62 |
| Threonine | 0.96 (0.82, 1.13) | 0.62 |  | 1.01 (0.87, 1.18) | 0.88 |  | 0.91 (0.72, 1.15) | 0.43 |  | 0.74 |
| Tryptophan | 0.94 (0.81, 1.09) | 0.42 |  | 1.03 (0.89, 1.20) | 0.68 |  | 0.99 (0.79, 1.24) | 0.92 |  | 0.69 |
| Non-essential amino acids | 0.94 (0.79, 1.11) | 0.46 |  | 1.03 (0.87, 1.22) | 0.74 |  | 0.95 (0.74, 1.22) | 0.69 |  | 0.73 |
| Alanine | 0.97 (0.81, 1.17) | 0.76 |  | 0.98 (0.82, 1.16) | 0.78 |  | 0.85 (0.65, 1.11) | 0.24 |  | 0.67 |
| Arginine | 0.98 (0.84, 1.13) | 0.76 |  | 0.97 (0.84, 1.12) | 0.68 |  | 0.85 (0.69, 1.05) | 0.13 |  | 0.52 |
| Aspartic acid | 1.04 (0.92, 1.18) | 0.54 |  | 1.02 (0.90, 1.15) | 0.76 |  | 0.88 (0.74, 1.06) | 0.19 |  | 0.34 |
| Cystine | 0.85 (0.69, 1.05) | 0.14 |  | 1.00 (0.81, 1.23) | 0.98 |  | 0.92 (0.68, 1.26) | 0.62 |  | 0.58 |
| Glutamic acid | 0.89 (0.74, 1.09) | 0.26 |  | 1.05 (0.86, 1.27) | 0.64 |  | 1.06 (0.79, 1.41) | 0.70 |  | 0.45 |
| Glycine | 0.97 (0.80, 1.17) | 0.76 |  | 0.95 (0.80, 1.14) | 0.61 |  | 0.86 (0.66, 1.13) | 0.28 |  | 0.77 |
| Hydroxyproline | 1.23 (1.05, 1.43) | 0.008 |  | 1.00 (0.83, 1.20) | 1.00 |  | 1.18 (0.89, 1.56) | 0.25 |  | 0.23 |
| Proline | 0.87 (0.72, 1.04) | 0.13 |  | 1.02 (0.85, 1.23) | 0.81 |  | 1.07 (0.82, 1.41) | 0.61 |  | 0.33 |
| Serine | 0.94 (0.81, 1.10) | 0.44 |  | 1.07 (0.92, 1.24) | 0.40 |  | 0.98 (0.78, 1.23) | 0.89 |  | 0.51 |
| Tyrosine | 0.95 (0.81, 1.10) | 0.48 |  | 1.05 (0.91, 1.22) | 0.51 |  | 0.99 (0.78, 1.24) | 0.91 |  | 0.62 |
| Total protein (g/1000 kcal) | 1.00 (0.92, 1.08) | 1.00 |  | 0.98 (0.91, 1.06) | 0.62 |  | 0.98 (0.88, 1.11) | 0.79 |  | 0.94 |

1. Hazard ratios modelled per 1 sex-specific SD increment in dietary amino acids or total protein. Based on multivariable model stratified by sex and centre, and adjusted for age (continuous), smoking (never, former, current<10, 10-19, 20+ cigarettes/day, unknown), calibrated alcohol consumption (non-drinkers (<0.1), 0.1-4.9, 5.0-14.9, 15-29.9, 30-59.9, 60+ g/day), physical activity (inactive, moderately inactive, moderately active, active, unknown), employment status (employed or student, neither employed nor student, unknown), highest level of education completed (none or primary, secondary, vocational or university, unknown), history of diabetes (yes, no, unknown), prior hypertension (yes, no , unknown), prior hyperlipidaemia (yes, no, unknown), calibrated energy intake (continuous), body mass index (<22.5, 22.5-24.9, 25.0-27.4, 27.5-29.9, ≥30.0 kg/m^2^, unknown).
2. Tests of trend were performed using the calibrated intake (continuous).
3. Tests of heterogeneity of trend by body mass index were obtained assuming independence of risk by age at recruitment using a meta-analysis method.

Supplementary table 19: Hazard ratios (95% confidence intervals)^1^ for **ischaemic** stroke by increments of calibrated intakes of dietary amino acids, stratified by **smoking status**

| Amino acids (percent of total protein) | Never smoker  (1631 cases) | |  | Former smoker  (1225 cases) | |  | Current smoker  (1407 cases) | |  |  |
| --- | --- | --- | --- | --- | --- | --- | --- | --- | --- | --- |
|  | HR (95% CI) | P for trend^2^ |  | HR (95% CI) | P for trend^2^ |  | HR (95% CI) | P for trend^2^ |  | P-het^3^ |
| Branched-chain amino acids | 0.95 (0.86, 1.04) | 0.27 |  | 0.92 (0.84, 1.02) | 0.12 |  | 0.90 (0.83, 0.99) | 0.03 |  | 0.79 |
| Isoleucine | 0.95 (0.86, 1.05) | 0.32 |  | 0.93 (0.84, 1.03) | 0.15 |  | 0.90 (0.82, 0.99) | 0.03 |  | 0.75 |
| Leucine | 0.95 (0.86, 1.04) | 0.26 |  | 0.93 (0.84, 1.03) | 0.15 |  | 0.90 (0.82, 0.99) | 0.03 |  | 0.81 |
| Valine | 0.95 (0.86, 1.04) | 0.24 |  | 0.92 (0.83, 1.01) | 0.07 |  | 0.91 (0.83, 0.99) | 0.03 |  | 0.78 |
| Other essential amino acids | 0.96 (0.87, 1.06) | 0.43 |  | 0.94 (0.84, 1.04) | 0.21 |  | 0.90 (0.82, 1.00) | 0.04 |  | 0.71 |
| Histidine | 0.97 (0.87, 1.08) | 0.55 |  | 0.94 (0.84, 1.05) | 0.26 |  | 0.91 (0.82, 1.01) | 0.07 |  | 0.71 |
| Lysine | 0.97 (0.87, 1.08) | 0.57 |  | 0.93 (0.83, 1.05) | 0.24 |  | 0.91 (0.82, 1.01) | 0.06 |  | 0.68 |
| Methionine | 0.96 (0.86, 1.07) | 0.47 |  | 0.93 (0.83, 1.05) | 0.23 |  | 0.91 (0.81, 1.01) | 0.07 |  | 0.76 |
| Phenylalanine | 0.95 (0.86, 1.04) | 0.27 |  | 0.93 (0.84, 1.03) | 0.15 |  | 0.90 (0.82, 0.98) | 0.02 |  | 0.73 |
| Threonine | 0.96 (0.87, 1.05) | 0.36 |  | 0.94 (0.85, 1.04) | 0.21 |  | 0.91 (0.83, 0.99) | 0.04 |  | 0.72 |
| Tryptophan | 0.95 (0.87, 1.05) | 0.31 |  | 0.94 (0.85, 1.04) | 0.21 |  | 0.92 (0.84, 1.00) | 0.05 |  | 0.83 |
| Non-essential amino acids | 0.95 (0.86, 1.05) | 0.33 |  | 0.93 (0.83, 1.04) | 0.19 |  | 0.89 (0.81, 0.98) | 0.02 |  | 0.66 |
| Alanine | 1.00 (0.90, 1.12) | 0.93 |  | 0.97 (0.86, 1.09) | 0.58 |  | 0.92 (0.83, 1.02) | 0.12 |  | 0.52 |
| Arginine | 1.01 (0.93, 1.10) | 0.81 |  | 0.98 (0.89, 1.07) | 0.65 |  | 0.93 (0.85, 1.00) | 0.06 |  | 0.32 |
| Aspartic acid | 0.99 (0.92, 1.07) | 0.84 |  | 0.96 (0.89, 1.04) | 0.34 |  | 0.92 (0.86, 0.99) | 0.02 |  | 0.36 |
| Cystine | 1.05 (0.93, 1.18) | 0.46 |  | 0.97 (0.85, 1.10) | 0.62 |  | 0.90 (0.80, 1.02) | 0.10 |  | 0.24 |
| Glutamic acid | 0.91 (0.81, 1.03) | 0.15 |  | 0.91 (0.80, 1.03) | 0.12 |  | 0.87 (0.78, 0.98) | 0.02 |  | 0.84 |
| Glycine | 1.04 (0.93, 1.16) | 0.50 |  | 1.01 (0.90, 1.14) | 0.86 |  | 0.94 (0.85, 1.04) | 0.25 |  | 0.42 |
| Hydroxyproline | 1.00 (0.88, 1.13) | 0.99 |  | 1.09 (0.96, 1.23) | 0.20 |  | 0.94 (0.84, 1.06) | 0.34 |  | 0.28 |
| Proline | 0.87 (0.78, 0.98) | 0.02 |  | 0.87 (0.77, 0.98) | 0.02 |  | 0.89 (0.80, 0.99) | 0.03 |  | 0.96 |
| Serine | 0.95 (0.86, 1.04) | 0.24 |  | 0.93 (0.84, 1.02) | 0.14 |  | 0.90 (0.82, 0.98) | 0.02 |  | 0.74 |
| Tyrosine | 0.94 (0.86, 1.03) | 0.21 |  | 0.92 (0.83, 1.01) | 0.09 |  | 0.91 (0.83, 0.99) | 0.04 |  | 0.87 |
| Total protein (g/1000 kcal) | 0.98 (0.94, 1.03) | 0.44 |  | 0.98 (0.93, 1.03) | 0.34 |  | 0.96 (0.92, 1.01) | 0.10 |  | 0.85 |

1. Hazard ratios modelled per 1 sex-specific SD increment in dietary amino acids or total protein. Based on multivariable model stratified by sex and centre, and adjusted for age (continuous), smoking (never, former, current<10, 10-19, 20+ cigarettes/day, unknown), calibrated alcohol consumption (non-drinkers (<0.1), 0.1-4.9, 5.0-14.9, 15-29.9, 30-59.9, 60+ g/day), physical activity (inactive, moderately inactive, moderately active, active, unknown), employment status (employed or student, neither employed nor student, unknown), highest level of education completed (none or primary, secondary, vocational or university, unknown), history of diabetes (yes, no, unknown), prior hypertension (yes, no , unknown), prior hyperlipidaemia (yes, no, unknown), calibrated energy intake (continuous), body mass index (<22.5, 22.5-24.9, 25.0-27.4, 27.5-29.9, ≥30.0 kg/m^2^, unknown).
2. Tests of trend were performed using the calibrated intake (continuous).
3. Tests of heterogeneity of trend by smoking status were obtained assuming independence of risk by age at recruitment using a meta-analysis method.

Supplementary table 20: Hazard ratios (95% confidence intervals)^1^ for **haemorrhagic** stroke by increments of calibrated intakes of dietary amino acids, stratified by **smoking status**

| Amino acids (percent of total protein) | Never smoker  (522 cases) | |  | Former smoker  (399 cases) | |  | Current smoker  (446 cases) | |  |  |
| --- | --- | --- | --- | --- | --- | --- | --- | --- | --- | --- |
|  | HR (95% CI) | P for trend^2^ |  | HR (95% CI) | P for trend^2^ |  | HR (95% CI) | P for trend^2^ |  | P-het^3^ |
| Branched-chain amino acids | 0.98 (0.83, 1.17) | 0.85 |  | 1.08 (0.91, 1.29) | 0.39 |  | 0.88 (0.74, 1.04) | 0.13 |  | 0.24 |
| Isoleucine | 0.99 (0.83, 1.18) | 0.94 |  | 1.07 (0.89, 1.27) | 0.49 |  | 0.88 (0.74, 1.04) | 0.13 |  | 0.29 |
| Leucine | 0.98 (0.82, 1.17) | 0.82 |  | 1.09 (0.91, 1.30) | 0.36 |  | 0.88 (0.74, 1.04) | 0.13 |  | 0.23 |
| Valine | 0.98 (0.83, 1.16) | 0.82 |  | 1.08 (0.91, 1.28) | 0.37 |  | 0.88 (0.75, 1.03) | 0.12 |  | 0.22 |
| Other essential amino acids | 0.98 (0.82, 1.18) | 0.84 |  | 1.10 (0.92, 1.32) | 0.30 |  | 0.87 (0.73, 1.04) | 0.12 |  | 0.19 |
| Histidine | 0.98 (0.81, 1.18) | 0.82 |  | 1.08 (0.89, 1.30) | 0.45 |  | 0.86 (0.72, 1.04) | 0.12 |  | 0.27 |
| Lysine | 0.98 (0.81, 1.19) | 0.86 |  | 1.13 (0.94, 1.37) | 0.20 |  | 0.86 (0.70, 1.04) | 0.12 |  | 0.13 |
| Methionine | 0.96 (0.79, 1.17) | 0.72 |  | 1.11 (0.92, 1.35) | 0.28 |  | 0.88 (0.72, 1.07) | 0.19 |  | 0.23 |
| Phenylalanine | 0.99 (0.83, 1.18) | 0.93 |  | 1.06 (0.89, 1.27) | 0.50 |  | 0.89 (0.75, 1.05) | 0.17 |  | 0.35 |
| Threonine | 0.98 (0.83, 1.17) | 0.84 |  | 1.09 (0.91, 1.30) | 0.35 |  | 0.87 (0.73, 1.03) | 0.10 |  | 0.19 |
| Tryptophan | 0.98 (0.83, 1.16) | 0.80 |  | 1.09 (0.92, 1.30) | 0.32 |  | 0.91 (0.78, 1.07) | 0.25 |  | 0.32 |
| Non-essential amino acids | 1.00 (0.83, 1.21) | 1.00 |  | 1.05 (0.87, 1.28) | 0.60 |  | 0.90 (0.75, 1.08) | 0.25 |  | 0.48 |
| Alanine | 0.98 (0.80, 1.19) | 0.83 |  | 1.02 (0.83, 1.25) | 0.85 |  | 0.86 (0.71, 1.05) | 0.14 |  | 0.48 |
| Arginine | 1.00 (0.85, 1.17) | 0.99 |  | 0.99 (0.84, 1.16) | 0.86 |  | 0.87 (0.75, 1.02) | 0.08 |  | 0.40 |
| Aspartic acid | 1.03 (0.90, 1.18) | 0.70 |  | 1.07 (0.93, 1.23) | 0.34 |  | 0.93 (0.81, 1.06) | 0.27 |  | 0.33 |
| Cystine | 0.91 (0.73, 1.15) | 0.44 |  | 0.96 (0.75, 1.22) | 0.72 |  | 0.89 (0.71, 1.11) | 0.29 |  | 0.90 |
| Glutamic acid | 0.99 (0.80, 1.23) | 0.96 |  | 1.05 (0.84, 1.32) | 0.66 |  | 0.92 (0.75, 1.13) | 0.45 |  | 0.70 |
| Glycine | 1.02 (0.84, 1.25) | 0.84 |  | 0.92 (0.74, 1.13) | 0.42 |  | 0.90 (0.74, 1.10) | 0.29 |  | 0.64 |
| Hydroxyproline | 1.19 (0.97, 1.46) | 0.10 |  | 1.09 (0.89, 1.34) | 0.39 |  | 1.14 (0.96, 1.35) | 0.13 |  | 0.85 |
| Proline | 0.94 (0.77, 1.16) | 0.58 |  | 1.05 (0.85, 1.30) | 0.66 |  | 0.93 (0.77, 1.13) | 0.46 |  | 0.68 |
| Serine | 1.01 (0.85, 1.20) | 0.90 |  | 1.11 (0.94, 1.32) | 0.22 |  | 0.90 (0.77, 1.06) | 0.22 |  | 0.22 |
| Tyrosine | 0.99 (0.84, 1.17) | 0.92 |  | 1.12 (0.94, 1.32) | 0.20 |  | 0.90 (0.77, 1.07) | 0.23 |  | 0.21 |
| Total protein (g/1000 kcal) | 1.00 (0.91, 1.09) | 0.96 |  | 1.07 (0.98, 1.17) | 0.13 |  | 0.92 (0.84, 1.00) | 0.04 |  | 0.04 |

1. Hazard ratios modelled per 1 sex-specific SD increment in dietary amino acids or total protein. Based on multivariable model stratified by sex and centre, and adjusted for age (continuous), smoking (never, former, current<10, 10-19, 20+ cigarettes/day, unknown), calibrated alcohol consumption (non-drinkers (<0.1), 0.1-4.9, 5.0-14.9, 15-29.9, 30-59.9, 60+ g/day), physical activity (inactive, moderately inactive, moderately active, active, unknown), employment status (employed or student, neither employed nor student, unknown), highest level of education completed (none or primary, secondary, vocational or university, unknown), history of diabetes (yes, no, unknown), prior hypertension (yes, no , unknown), prior hyperlipidaemia (yes, no, unknown), calibrated energy intake (continuous), body mass index (<22.5, 22.5-24.9, 25.0-27.4, 27.5-29.9, ≥30.0 kg/m^2^, unknown).
2. Tests of trend were performed using the calibrated intake (continuous).
3. Tests of heterogeneity of trend by smoking status were obtained assuming independence of risk by age at recruitment using a meta-analysis method.

Supplementary table 21: Hazard ratios (95% confidence intervals)^1^ for **ischaemic** stroke by increments of calibrated intakes of dietary amino acids, stratified by **alcohol drinking**

| Amino acids (percent of total protein) | Non drinkers  (466 cases) | |  | Moderate drinkers <15.0 g/day  (2217 cases) | |  | Heavy drinkers ≥15.0 g/day  (1612 cases) | |  |  |
| --- | --- | --- | --- | --- | --- | --- | --- | --- | --- | --- |
|  | HR (95% CI) | P for trend^2^ |  | HR (95% CI) | P for trend^2^ |  | HR (95% CI) | P for trend^2^ |  | P-het^3^ |
| Branched-chain amino acids | 0.86 (0.73, 1.01) | 0.07 |  | 0.96 (0.89, 1.05) | 0.38 |  | 0.89 (0.82, 0.97) | 0.008 |  | 0.31 |
| Isoleucine | 0.86 (0.73, 1.02) | 0.08 |  | 0.96 (0.89, 1.05) | 0.40 |  | 0.90 (0.83, 0.98) | 0.01 |  | 0.35 |
| Leucine | 0.86 (0.72, 1.02) | 0.08 |  | 0.97 (0.89, 1.05) | 0.42 |  | 0.89 (0.82, 0.97) | 0.008 |  | 0.29 |
| Valine | 0.86 (0.73, 1.01) | 0.06 |  | 0.96 (0.89, 1.04) | 0.32 |  | 0.89 (0.83, 0.97) | 0.006 |  | 0.30 |
| Other essential amino acids | 0.87 (0.73, 1.03) | 0.11 |  | 0.97 (0.89, 1.06) | 0.55 |  | 0.90 (0.82, 0.98) | 0.02 |  | 0.32 |
| Histidine | 0.87 (0.73, 1.04) | 0.13 |  | 0.99 (0.90, 1.08) | 0.80 |  | 0.90 (0.82, 0.99) | 0.02 |  | 0.25 |
| Lysine | 0.88 (0.73, 1.06) | 0.16 |  | 0.97 (0.88, 1.07) | 0.55 |  | 0.91 (0.82, 1.00) | 0.04 |  | 0.47 |
| Methionine | 0.86 (0.71, 1.04) | 0.11 |  | 0.97 (0.88, 1.06) | 0.48 |  | 0.91 (0.82, 1.00) | 0.05 |  | 0.45 |
| Phenylalanine | 0.86 (0.73, 1.01) | 0.06 |  | 0.97 (0.89, 1.06) | 0.49 |  | 0.88 (0.81, 0.96) | 0.004 |  | 0.20 |
| Threonine | 0.88 (0.74, 1.03) | 0.11 |  | 0.97 (0.89, 1.05) | 0.46 |  | 0.90 (0.83, 0.98) | 0.01 |  | 0.36 |
| Tryptophan | 0.88 (0.75, 1.03) | 0.11 |  | 0.99 (0.92, 1.08) | 0.89 |  | 0.88 (0.81, 0.95) | 0.002 |  | 0.07 |
| Non-essential amino acids | 0.85 (0.72, 1.01) | 0.07 |  | 0.97 (0.89, 1.06) | 0.49 |  | 0.88 (0.80, 0.96) | 0.006 |  | 0.22 |
| Alanine | 0.92 (0.77, 1.11) | 0.38 |  | 0.99 (0.90, 1.09) | 0.87 |  | 0.93 (0.85, 1.03) | 0.16 |  | 0.61 |
| Arginine | 0.93 (0.81, 1.07) | 0.32 |  | 1.00 (0.93, 1.08) | 0.95 |  | 0.94 (0.87, 1.01) | 0.09 |  | 0.40 |
| Aspartic acid | 0.93 (0.82, 1.05) | 0.22 |  | 0.99 (0.93, 1.05) | 0.68 |  | 0.93 (0.87, 0.99) | 0.03 |  | 0.40 |
| Cystine | 0.86 (0.70, 1.06) | 0.16 |  | 1.03 (0.93, 1.14) | 0.56 |  | 0.91 (0.82, 1.02) | 0.11 |  | 0.16 |
| Glutamic acid | 0.81 (0.66, 0.99) | 0.04 |  | 0.96 (0.86, 1.06) | 0.39 |  | 0.85 (0.76, 0.94) | 0.002 |  | 0.16 |
| Glycine | 0.95 (0.79, 1.14) | 0.56 |  | 1.02 (0.93, 1.13) | 0.63 |  | 0.96 (0.87, 1.06) | 0.44 |  | 0.60 |
| Hydroxyproline | 0.82 (0.66, 1.01) | 0.06 |  | 1.02 (0.91, 1.13) | 0.78 |  | 1.04 (0.94, 1.16) | 0.42 |  | 0.12 |
| Proline | 0.80 (0.65, 0.97) | 0.03 |  | 0.92 (0.83, 1.01) | 0.08 |  | 0.85 (0.77, 0.94) | 0.001 |  | 0.37 |
| Serine | 0.85 (0.73, 1.00) | 0.05 |  | 0.97 (0.89, 1.05) | 0.42 |  | 0.89 (0.82, 0.96) | 0.004 |  | 0.21 |
| Tyrosine | 0.85 (0.73, 1.01) | 0.06 |  | 0.96 (0.89, 1.04) | 0.33 |  | 0.90 (0.83, 0.97) | 0.008 |  | 0.32 |
| Total protein (g/1000 kcal) | 0.99 (0.92, 1.07) | 0.86 |  | 0.98 (0.94, 1.02) | 0.43 |  | 0.95 (0.91, 0.99) | 0.02 |  | 0.46 |

1. Hazard ratios modelled per 1 sex-specific SD increment in dietary amino acids or total protein. Based on multivariable model stratified by sex and centre, and adjusted for age (continuous), smoking (never, former, current<10, 10-19, 20+ cigarettes/day, unknown), calibrated alcohol consumption (non-drinkers (<0.1), 0.1-4.9, 5.0-14.9, 15-29.9, 30-59.9, 60+ g/day), physical activity (inactive, moderately inactive, moderately active, active, unknown), employment status (employed or student, neither employed nor student, unknown), highest level of education completed (none or primary, secondary, vocational or university, unknown), history of diabetes (yes, no, unknown), prior hypertension (yes, no , unknown), prior hyperlipidaemia (yes, no, unknown), calibrated energy intake (continuous), body mass index (<22.5, 22.5-24.9, 25.0-27.4, 27.5-29.9, ≥30.0 kg/m^2^, unknown).
2. Tests of trend were performed using the calibrated intake (continuous).
3. Tests of heterogeneity of trend by alcohol drinking were obtained assuming independence of risk by age at recruitment using a meta-analysis method.

Supplementary table 22: Hazard ratios (95% confidence intervals)^1^ for **haemorrhagic** stroke by increments of calibrated intakes of dietary amino acids, stratified by **alcohol drinking**

| Amino acids (percent of total protein) | Non drinkers  (122 cases) | |  | Moderate drinkers <15.0 g/day  (728 cases) | |  | Heavy drinkers ≥15.0 g/day  (525 cases) | |  |  |
| --- | --- | --- | --- | --- | --- | --- | --- | --- | --- | --- |
|  | HR (95% CI) | P for trend^2^ |  | HR (95% CI) | P for trend^2^ |  | HR (95% CI) | P for trend^2^ |  | P-het^3^ |
| Branched-chain amino acids | 0.87 (0.63, 1.20) | 0.39 |  | 1.04 (0.90, 1.20) | 0.58 |  | 0.93 (0.80, 1.08) | 0.33 |  | 0.42 |
| Isoleucine | 0.85 (0.61, 1.18) | 0.32 |  | 1.05 (0.90, 1.22) | 0.53 |  | 0.93 (0.80, 1.08) | 0.33 |  | 0.36 |
| Leucine | 0.88 (0.63, 1.23) | 0.44 |  | 1.04 (0.90, 1.21) | 0.57 |  | 0.92 (0.79, 1.08) | 0.32 |  | 0.44 |
| Valine | 0.87 (0.63, 1.18) | 0.37 |  | 1.03 (0.90, 1.19) | 0.65 |  | 0.93 (0.81, 1.08) | 0.35 |  | 0.45 |
| Other essential amino acids | 0.86 (0.61, 1.20) | 0.38 |  | 1.06 (0.91, 1.24) | 0.42 |  | 0.92 (0.78, 1.08) | 0.29 |  | 0.30 |
| Histidine | 0.86 (0.61, 1.22) | 0.40 |  | 1.07 (0.91, 1.25) | 0.41 |  | 0.89 (0.75, 1.05) | 0.16 |  | 0.22 |
| Lysine | 0.82 (0.57, 1.17) | 0.28 |  | 1.09 (0.93, 1.28) | 0.30 |  | 0.92 (0.77, 1.10) | 0.35 |  | 0.21 |
| Methionine | 0.82 (0.57, 1.17) | 0.27 |  | 1.07 (0.91, 1.26) | 0.42 |  | 0.93 (0.78, 1.11) | 0.41 |  | 0.29 |
| Phenylalanine | 0.90 (0.65, 1.26) | 0.55 |  | 1.03 (0.89, 1.20) | 0.67 |  | 0.93 (0.80, 1.08) | 0.34 |  | 0.55 |
| Threonine | 0.88 (0.64, 1.21) | 0.44 |  | 1.06 (0.91, 1.22) | 0.47 |  | 0.91 (0.78, 1.06) | 0.22 |  | 0.31 |
| Tryptophan | 0.92 (0.68, 1.25) | 0.60 |  | 1.05 (0.91, 1.21) | 0.50 |  | 0.94 (0.81, 1.08) | 0.37 |  | 0.48 |
| Non-essential amino acids | 0.92 (0.65, 1.30) | 0.64 |  | 1.05 (0.90, 1.23) | 0.55 |  | 0.91 (0.77, 1.08) | 0.27 |  | 0.46 |
| Alanine | 0.89 (0.63, 1.25) | 0.50 |  | 1.05 (0.89, 1.24) | 0.56 |  | 0.86 (0.72, 1.03) | 0.10 |  | 0.25 |
| Arginine | 0.98 (0.75, 1.29) | 0.90 |  | 1.02 (0.89, 1.16) | 0.79 |  | 0.87 (0.75, 1.00) | 0.05 |  | 0.26 |
| Aspartic acid | 0.95 (0.75, 1.21) | 0.69 |  | 1.07 (0.95, 1.20) | 0.27 |  | 0.95 (0.84, 1.07) | 0.37 |  | 0.33 |
| Cystine | 0.95 (0.62, 1.46) | 0.82 |  | 0.99 (0.82, 1.20) | 0.94 |  | 0.84 (0.68, 1.03) | 0.09 |  | 0.50 |
| Glutamic acid | 0.94 (0.62, 1.43) | 0.79 |  | 1.05 (0.87, 1.25) | 0.64 |  | 0.92 (0.77, 1.11) | 0.41 |  | 0.65 |
| Glycine | 0.91 (0.64, 1.29) | 0.60 |  | 1.06 (0.89, 1.25) | 0.52 |  | 0.84 (0.70, 1.01) | 0.06 |  | 0.18 |
| Hydroxyproline | 1.01 (0.71, 1.44) | 0.94 |  | 1.36 (1.17, 1.58) | <0.001 |  | 0.96 (0.81, 1.15) | 0.68 |  | 0.01 |
| Proline | 0.81 (0.53, 1.22) | 0.31 |  | 0.99 (0.83, 1.17) | 0.90 |  | 0.97 (0.81, 1.15) | 0.70 |  | 0.67 |
| Serine | 0.94 (0.68, 1.29) | 0.69 |  | 1.05 (0.91, 1.21) | 0.49 |  | 0.96 (0.83, 1.11) | 0.58 |  | 0.63 |
| Tyrosine | 0.86 (0.62, 1.19) | 0.36 |  | 1.06 (0.92, 1.22) | 0.41 |  | 0.96 (0.83, 1.11) | 0.61 |  | 0.41 |
| Total protein (g/1000 kcal) | 0.98 (0.84, 1.15) | 0.81 |  | 1.05 (0.97, 1.12) | 0.23 |  | 0.93 (0.86, 1.00) | 0.06 |  | 0.09 |

1. Hazard ratios modelled per 1 sex-specific SD increment in dietary amino acids or total protein. Based on multivariable model stratified by sex and centre, and adjusted for age (continuous), smoking (never, former, current<10, 10-19, 20+ cigarettes/day, unknown), calibrated alcohol consumption (non-drinkers (<0.1), 0.1-4.9, 5.0-14.9, 15-29.9, 30-59.9, 60+ g/day), physical activity (inactive, moderately inactive, moderately active, active, unknown), employment status (employed or student, neither employed nor student, unknown), highest level of education completed (none or primary, secondary, vocational or university, unknown), history of diabetes (yes, no, unknown), prior hypertension (yes, no , unknown), prior hyperlipidaemia (yes, no, unknown), calibrated energy intake (continuous), body mass index (<22.5, 22.5-24.9, 25.0-27.4, 27.5-29.9, ≥30.0 kg/m^2^, unknown).
2. Tests of trend were performed using the calibrated intake (continuous).
3. Tests of heterogeneity of trend by alcohol drinking were obtained assuming independence of risk by age at recruitment using a meta-analysis method.

Supplementary table 23: Hazard ratios (95% confidence intervals)^1^ for **ischaemic** stroke by increments of calibrated intakes of dietary amino acids, stratified by **prior disease status**

| Amino acids (percent of total protein) | No disease history  (1300 cases) | |  | History of diabetes, hypertension or hyperlipidemia (1826 cases) | |  |  |
| --- | --- | --- | --- | --- | --- | --- | --- |
|  | HR (95% CI) | P for trend^2^ |  | HR (95% CI) | P for trend^2^ |  | P-het^3^ |
| Branched-chain amino acids | 0.96 (0.87, 1.07) | 0.48 |  | 0.91 (0.84, 0.99) | 0.03 |  | 0.40 |
| Isoleucine | 0.96 (0.87, 1.07) | 0.47 |  | 0.91 (0.83, 0.99) | 0.03 |  | 0.42 |
| Leucine | 0.97 (0.87, 1.08) | 0.56 |  | 0.91 (0.83, 0.99) | 0.03 |  | 0.35 |
| Valine | 0.96 (0.87, 1.06) | 0.39 |  | 0.91 (0.84, 0.99) | 0.03 |  | 0.45 |
| Other essential amino acids | 0.99 (0.89, 1.10) | 0.81 |  | 0.91 (0.84, 1.00) | 0.05 |  | 0.29 |
| Histidine | 1.01 (0.90, 1.13) | 0.87 |  | 0.92 (0.84, 1.01) | 0.07 |  | 0.20 |
| Lysine | 1.00 (0.89, 1.12) | 1.00 |  | 0.91 (0.83, 1.01) | 0.07 |  | 0.24 |
| Methionine | 0.98 (0.87, 1.10) | 0.77 |  | 0.91 (0.83, 1.01) | 0.06 |  | 0.33 |
| Phenylalanine | 0.96 (0.86, 1.07) | 0.47 |  | 0.91 (0.84, 0.99) | 0.03 |  | 0.44 |
| Threonine | 0.98 (0.88, 1.09) | 0.71 |  | 0.91 (0.84, 0.99) | 0.03 |  | 0.28 |
| Tryptophan | 0.97 (0.88, 1.07) | 0.56 |  | 0.92 (0.85, 1.00) | 0.06 |  | 0.46 |
| Non-essential amino acids | 0.98 (0.87, 1.10) | 0.69 |  | 0.90 (0.82, 0.98) | 0.02 |  | 0.25 |
| Alanine | 1.05 (0.93, 1.18) | 0.44 |  | 0.91 (0.83, 1.01) | 0.06 |  | 0.08 |
| Arginine | 1.05 (0.96, 1.16) | 0.26 |  | 0.93 (0.86, 1.00) | 0.05 |  | 0.04 |
| Aspartic acid | 1.01 (0.93, 1.10) | 0.80 |  | 0.94 (0.88, 1.00) | 0.05 |  | 0.15 |
| Cystine | 1.06 (0.93, 1.22) | 0.37 |  | 0.90 (0.81, 1.00) | 0.06 |  | 0.06 |
| Glutamic acid | 0.93 (0.82, 1.07) | 0.32 |  | 0.88 (0.79, 0.98) | 0.02 |  | 0.52 |
| Glycine | 1.09 (0.97, 1.23) | 0.14 |  | 0.93 (0.85, 1.02) | 0.14 |  | 0.04 |
| Hydroxyproline | 1.00 (0.89, 1.13) | 0.93 |  | 0.98 (0.88, 1.10) | 0.77 |  | 0.79 |
| Proline | 0.88 (0.78, 1.00) | 0.05 |  | 0.88 (0.79, 0.98) | 0.02 |  | 0.96 |
| Serine | 0.95 (0.86, 1.06) | 0.37 |  | 0.92 (0.85, 1.00) | 0.05 |  | 0.57 |
| Tyrosine | 0.95 (0.86, 1.05) | 0.30 |  | 0.92 (0.85, 1.01) | 0.07 |  | 0.70 |
| Total protein (g/1000 kcal) | 1.00 (0.95, 1.06) | 0.89 |  | 0.96 (0.92, 1.00) | 0.04 |  | 0.16 |

1. Hazard ratios modelled per 1 sex-specific SD increment in dietary amino acids or total protein. Based on multivariable model stratified by sex and centre, and adjusted for age (continuous), smoking (never, former, current<10, 10-19, 20+ cigarettes/day, unknown), calibrated alcohol consumption (non-drinkers (<0.1), 0.1-4.9, 5.0-14.9, 15-29.9, 30-59.9, 60+ g/day), physical activity (inactive, moderately inactive, moderately active, active, unknown), employment status (employed or student, neither employed nor student, unknown), highest level of education completed (none or primary, secondary, vocational or university, unknown), history of diabetes (yes, no, unknown), prior hypertension (yes, no , unknown), prior hyperlipidaemia (yes, no, unknown), calibrated energy intake (continuous), body mass index (<22.5, 22.5-24.9, 25.0-27.4, 27.5-29.9, ≥30.0 kg/m^2^, unknown).
2. Tests of trend were performed using the calibrated intake (continuous).
3. Tests of heterogeneity of trend by prior disease status were obtained assuming independence of risk by age at recruitment using a meta-analysis method.

Supplementary table 24: Hazard ratios (95% confidence intervals)^1^ for **haemorrhagic** stroke by increments of calibrated intakes of dietary amino acids, stratified by **prior disease status**

| Amino acids (percent of total protein) | No disease history  (541 cases) | |  | History of diabetes, hypertension or hyperlipidemia (493 cases) | |  |  |  |
| --- | --- | --- | --- | --- | --- | --- | --- | --- |
|  | HR (95% CI) | P for trend^2^ |  | HR (95% CI) | P for trend^2^ |  |  | P-het^3^ |
| Branched-chain amino acids | 0.92 (0.78, 1.08) | 0.30 |  | 0.88 (0.75, 1.05) | 0.15 |  |  | 0.76 |
| Isoleucine | 0.91 (0.77, 1.07) | 0.25 |  | 0.89 (0.75, 1.06) | 0.18 |  |  | 0.87 |
| Leucine | 0.92 (0.78, 1.09) | 0.33 |  | 0.87 (0.73, 1.04) | 0.13 |  |  | 0.69 |
| Valine | 0.92 (0.79, 1.08) | 0.32 |  | 0.89 (0.76, 1.05) | 0.18 |  |  | 0.78 |
| Other essential amino acids | 0.92 (0.77, 1.09) | 0.33 |  | 0.87 (0.73, 1.04) | 0.13 |  |  | 0.67 |
| Histidine | 0.91 (0.76, 1.08) | 0.28 |  | 0.84 (0.70, 1.01) | 0.06 |  |  | 0.54 |
| Lysine | 0.92 (0.77, 1.10) | 0.37 |  | 0.88 (0.72, 1.07) | 0.19 |  |  | 0.73 |
| Methionine | 0.92 (0.77, 1.10) | 0.36 |  | 0.87 (0.72, 1.06) | 0.16 |  |  | 0.69 |
| Phenylalanine | 0.93 (0.78, 1.10) | 0.38 |  | 0.87 (0.73, 1.04) | 0.12 |  |  | 0.63 |
| Threonine | 0.91 (0.77, 1.07) | 0.27 |  | 0.87 (0.74, 1.04) | 0.12 |  |  | 0.72 |
| Tryptophan | 0.92 (0.78, 1.08) | 0.32 |  | 0.89 (0.75, 1.05) | 0.16 |  |  | 0.75 |
| Non-essential amino acids | 0.94 (0.78, 1.12) | 0.48 |  | 0.87 (0.72, 1.04) | 0.12 |  |  | 0.54 |
| Alanine | 0.89 (0.74, 1.08) | 0.23 |  | 0.84 (0.69, 1.02) | 0.07 |  |  | 0.63 |
| Arginine | 0.90 (0.78, 1.05) | 0.18 |  | 0.85 (0.73, 0.99) | 0.04 |  |  | 0.59 |
| Aspartic acid | 0.95 (0.84, 1.08) | 0.46 |  | 0.93 (0.81, 1.06) | 0.30 |  |  | 0.82 |
| Cystine | 0.91 (0.73, 1.14) | 0.41 |  | 0.82 (0.65, 1.02) | 0.08 |  |  | 0.50 |
| Glutamic acid | 0.96 (0.78, 1.18) | 0.69 |  | 0.87 (0.70, 1.07) | 0.19 |  |  | 0.52 |
| Glycine | 0.89 (0.73, 1.08) | 0.23 |  | 0.85 (0.70, 1.03) | 0.10 |  |  | 0.73 |
| Hydroxyproline | 1.12 (0.95, 1.33) | 0.17 |  | 0.95 (0.77, 1.17) | 0.65 |  |  | 0.23 |
| Proline | 0.95 (0.78, 1.15) | 0.59 |  | 0.90 (0.74, 1.10) | 0.30 |  |  | 0.70 |
| Serine | 0.95 (0.81, 1.12) | 0.56 |  | 0.91 (0.77, 1.07) | 0.25 |  |  | 0.68 |
| Tyrosine | 0.94 (0.80, 1.10) | 0.44 |  | 0.91 (0.77, 1.08) | 0.28 |  |  | 0.81 |
| Total protein (g/1000 kcal) | 0.95 (0.88, 1.04) | 0.29 |  | 0.95 (0.88, 1.04) | 0.28 |  |  | 0.99 |

1. Hazard ratios modelled per 1 sex-specific SD increment in dietary amino acids or total protein. Based on multivariable model stratified by sex and centre, and adjusted for age (continuous), smoking (never, former, current<10, 10-19, 20+ cigarettes/day, unknown), calibrated alcohol consumption (non-drinkers (<0.1), 0.1-4.9, 5.0-14.9, 15-29.9, 30-59.9, 60+ g/day), physical activity (inactive, moderately inactive, moderately active, active, unknown), employment status (employed or student, neither employed nor student, unknown), highest level of education completed (none or primary, secondary, vocational or university, unknown), history of diabetes (yes, no, unknown), prior hypertension (yes, no , unknown), prior hyperlipidaemia (yes, no, unknown), calibrated energy intake (continuous), body mass index (<22.5, 22.5-24.9, 25.0-27.4, 27.5-29.9, ≥30.0 kg/m^2^, unknown).
2. Tests of trend were performed using the calibrated intake (continuous).
3. Tests of heterogeneity of trend by prior disease status were obtained assuming independence of risk by age at recruitment using a meta-analysis method.

Supplementary table 25: **Country-specific** and **pooled hazard ratios** (95% confidence intervals) for **ischaemic** stroke by increments of observed intakes of dietary amino acids.

| Amino acids (percent of total protein) | Country-specific hazard ratios (95% confidence intervals) ^1^ | | | | | | |  | Meta-analyses | |  |
| --- | --- | --- | --- | --- | --- | --- | --- | --- | --- | --- | --- |
|  | Denmark  (770 cases) | Germany  (398 cases) | Italy  (141 cases) | Netherlands  (325 cases) | Spain  (458 cases) | Sweden  (1807 cases) | UK  (396 cases) | | Fixed effects | Random effects | p-het^2^ |
| Branched-chain amino acids | 0.96 (0.91, 1.00) | 0.98 (0.91, 1.07) | 0.98 (0.85, 1.12) | 0.97 (0.89, 1.07) | 0.96 (0.90, 1.02) | 0.97 (0.93, 1.00) | 1.05 (0.98, 1.12) | | 0.97 (0.95, 0.99) | 0.97 (0.95, 0.99) | 0.51 |
| Isoleucine | 0.96 (0.91, 1.01) | 0.98 (0.91, 1.07) | 0.97 (0.85, 1.11) | 0.98 (0.89, 1.07) | 0.96 (0.91, 1.02) | 0.97 (0.93, 1.00) | 1.04 (0.98, 1.11) | | 0.97 (0.95, 1.00) | 0.97 (0.95, 1.00) | 0.58 |
| Leucine | 0.95 (0.91, 1.00) | 0.98 (0.90, 1.07) | 0.98 (0.86, 1.13) | 0.97 (0.88, 1.07) | 0.96 (0.90, 1.02) | 0.97 (0.93, 1.00) | 1.05 (0.98, 1.12) | | 0.97 (0.95, 0.99) | 0.97 (0.95, 0.99) | 0.49 |
| Valine | 0.95 (0.91, 1.00) | 0.99 (0.91, 1.07) | 0.97 (0.85, 1.12) | 0.97 (0.88, 1.06) | 0.95 (0.89, 1.01) | 0.96 (0.93, 1.00) | 1.05 (0.98, 1.12) | | 0.97 (0.95, 0.99) | 0.97 (0.95, 0.99) | 0.47 |
| Other essential amino acids | 0.96 (0.92, 1.01) | 1.00 (0.92, 1.08) | 0.97 (0.85, 1.10) | 0.98 (0.89, 1.07) | 0.97 (0.91, 1.03) | 0.97 (0.93, 1.00) | 1.04 (0.98, 1.11) | | 0.98 (0.95, 1.00) | 0.98 (0.95, 1.00) | 0.55 |
| Histidine | 0.97 (0.91, 1.02) | 1.00 (0.92, 1.09) | 0.97 (0.85, 1.11) | 0.98 (0.88, 1.08) | 0.98 (0.92, 1.05) | 0.97 (0.93, 1.00) | 1.05 (0.98, 1.12) | | 0.98 (0.96, 1.00) | 0.98 (0.96, 1.00) | 0.53 |
| Lysine | 0.97 (0.92, 1.03) | 1.01 (0.92, 1.10) | 0.95 (0.83, 1.09) | 0.98 (0.88, 1.09) | 0.97 (0.91, 1.04) | 0.96 (0.93, 1.00) | 1.05 (0.98, 1.12) | | 0.98 (0.95, 1.00) | 0.98 (0.95, 1.00) | 0.56 |
| Methionine | 0.96 (0.91, 1.02) | 1.01 (0.92, 1.10) | 0.96 (0.83, 1.12) | 0.97 (0.86, 1.08) | 0.97 (0.91, 1.04) | 0.96 (0.92, 1.00) | 1.05 (0.97, 1.12) | | 0.98 (0.95, 1.00) | 0.98 (0.95, 1.00) | 0.59 |
| Phenylalanine | 0.94 (0.90, 0.99) | 0.98 (0.90, 1.07) | 0.99 (0.86, 1.15) | 0.97 (0.88, 1.07) | 0.95 (0.89, 1.01) | 0.97 (0.93, 1.00) | 1.06 (0.98, 1.14) | | 0.97 (0.95, 0.99) | 0.97 (0.95, 1.00) | 0.34 |
| Threonine | 0.96 (0.92, 1.01) | 0.99 (0.92, 1.08) | 0.97 (0.85, 1.11) | 0.98 (0.89, 1.07) | 0.97 (0.92, 1.03) | 0.96 (0.93, 1.00) | 1.04 (0.98, 1.10) | | 0.98 (0.96, 1.00) | 0.98 (0.96, 1.00) | 0.57 |
| Tryptophan | 0.94 (0.89, 0.99) | 0.98 (0.90, 1.07) | 0.96 (0.84, 1.11) | 0.99 (0.88, 1.10) | 0.95 (0.89, 1.01) | 0.98 (0.94, 1.01) | 1.03 (0.96, 1.10) | | 0.97 (0.95, 0.99) | 0.97 (0.95, 0.99) | 0.64 |
| Non-essential amino acids | 0.94 (0.90, 1.00) | 0.98 (0.89, 1.07) | 1.00 (0.86, 1.15) | 0.96 (0.87, 1.07) | 0.96 (0.90, 1.03) | 0.97 (0.93, 1.00) | 1.06 (0.99, 1.14) | | 0.97 (0.95, 0.99) | 0.97 (0.95, 1.00) | 0.36 |
| Alanine | 0.98 (0.92, 1.03) | 1.01 (0.92, 1.11) | 0.98 (0.84, 1.14) | 0.96 (0.86, 1.08) | 1.01 (0.95, 1.08) | 0.97 (0.93, 1.01) | 1.06 (0.99, 1.13) | | 0.99 (0.97, 1.01) | 0.99 (0.97, 1.01) | 0.50 |
| Arginine | 0.97 (0.92, 1.02) | 1.01 (0.92, 1.10) | 0.97 (0.84, 1.12) | 0.97 (0.88, 1.08) | 1.01 (0.95, 1.08) | 0.98 (0.94, 1.02) | 1.06 (1.00, 1.13) | | 0.99 (0.97, 1.02) | 0.99 (0.97, 1.02) | 0.40 |
| Aspartic acid | 0.96 (0.92, 1.01) | 0.99 (0.91, 1.08) | 0.95 (0.82, 1.10) | 0.99 (0.90, 1.09) | 0.98 (0.92, 1.04) | 0.97 (0.94, 1.00) | 1.05 (0.99, 1.12) | | 0.98 (0.96, 1.00) | 0.98 (0.96, 1.00) | 0.44 |
| Cystine | 0.91 (0.84, 0.98) | 0.97 (0.84, 1.11) | 1.08 (0.87, 1.33) | 0.93 (0.79, 1.10) | 1.00 (0.90, 1.11) | 1.01 (0.96, 1.06) | 1.10 (0.99, 1.23) | | 0.99 (0.95, 1.02) | 0.99 (0.94, 1.04) | 0.14 |
| Glutamic acid | 0.91 (0.85, 0.97) | 0.94 (0.84, 1.06) | 1.04 (0.86, 1.24) | 0.94 (0.83, 1.07) | 0.93 (0.86, 1.02) | 0.96 (0.92, 1.01) | 1.07 (0.97, 1.18) | | 0.96 (0.93, 0.98) | 0.96 (0.92, 1.00) | 0.21 |
| Glycine | 0.99 (0.93, 1.05) | 1.02 (0.92, 1.14) | 1.01 (0.85, 1.20) | 0.97 (0.86, 1.09) | 1.04 (0.96, 1.13) | 0.98 (0.94, 1.03) | 1.07 (0.99, 1.16) | | 1.00 (0.98, 1.03) | 1.00 (0.98, 1.03) | 0.54 |
| Hydroxyproline | 1.04 (0.95, 1.13) | 1.03 (0.91, 1.16) | 1.19 (0.95, 1.49) | 0.98 (0.88, 1.10) | 1.14 (0.98, 1.31) | 0.99 (0.94, 1.05) | 1.10 (0.95, 1.28) | | 1.02 (0.98, 1.06) | 1.02 (0.98, 1.06) | 0.45 |
| Proline | 0.90 (0.84, 0.97) | 0.94 (0.83, 1.07) | 1.01 (0.83, 1.23) | 0.94 (0.82, 1.07) | 0.86 (0.78, 0.94) | 0.95 (0.90, 1.00) | 1.06 (0.94, 1.20) | | 0.93 (0.90, 0.96) | 0.93 (0.89, 0.98) | 0.20 |
| Serine | 0.95 (0.90, 1.00) | 0.98 (0.90, 1.08) | 1.00 (0.86, 1.16) | 0.97 (0.88, 1.07) | 0.93 (0.87, 1.00) | 0.96 (0.93, 1.00) | 1.07 (0.99, 1.15) | | 0.97 (0.94, 0.99) | 0.97 (0.94, 1.00) | 0.23 |
| Tyrosine | 0.95 (0.90, 1.00) | 0.98 (0.90, 1.08) | 0.96 (0.83, 1.10) | 0.98 (0.88, 1.08) | 0.94 (0.87, 1.00) | 0.96 (0.93, 1.00) | 1.05 (0.97, 1.13) | | 0.96 (0.94, 0.99) | 0.96 (0.94, 0.99) | 0.51 |
| Total protein (g/1000 kcal) | 0.98 (0.94, 1.01) | 1.00 (0.95, 1.06) | 0.99 (0.90, 1.09) | 0.96 (0.90, 1.03) | 0.99 (0.95, 1.04) | 0.98 (0.96, 1.01) | 1.00 (0.95, 1.05) | | 0.98 (0.97, 1.00) | 0.98 (0.97, 1.00) | 0.96 |

1. Hazard ratios modelled per 1 sex-specific SD increment in dietary amino acids or protein. Based on multivariable model stratified by sex and centre, and adjusted for age (continuous), smoking (never, former, current<10, 10-19, 20+ cigarettes/day, unknown), observed alcohol consumption (non-drinkers (<0.1), 0.1-4.9, 5.0-14.9, 15-29.9, 30-59.9, 60+ g/day), physical activity (inactive, moderately inactive, moderately active, active, unknown), employment status (employed or student, neither employed nor student, unknown), highest level of education completed (none or primary, secondary, vocational or university, unknown), history of diabetes (yes, no, unknown), prior hypertension (yes, no , unknown), prior hyperlipidaemia (yes, no, unknown), observed energy intake (continuous), body mass index (<22.5, 22.5-24.9, 25.0-27.4, 27.5-29.9, ≥30.0 kg/m^2^, unknown).
2. Tests of heterogeneity by country from the meta-analysis.

Supplementary table 26: **Country-specific** and **pooled hazard ratios** (95% confidence intervals) **for haemorrhagic** stroke by increments of observed intakes of dietary amino acids.

| Amino acids (percent of total protein) | Country-specific hazard ratios (95% confidence intervals) ^1^ | | | | | | |  | | Meta-analyses | |  |
| --- | --- | --- | --- | --- | --- | --- | --- | --- | --- | --- | --- | --- |
|  | Denmark  (309 cases) | Germany  (89 cases) | Italy  (84 cases) | Netherlands  (127 cases) | Spain  (128 cases) | Sweden  (382 cases) | UK  (256 cases) | | Fixed effects | | Random effects | p-het^2^ |
| Branched-chain amino acids | 1.02 (0.95, 1.10) | 1.16 (0.99, 1.35) | 0.75 (0.63, 0.89) | 0.88 (0.75, 1.02) | 0.89 (0.79, 1.00) | 1.02 (0.95, 1.10) | 0.98 (0.90, 1.07) | | 0.97 (0.93, 1.01) | | 0.96 (0.88, 1.03) | <0.001 |
| Isoleucine | 1.03 (0.95, 1.11) | 1.17 (1.00, 1.36) | 0.76 (0.64, 0.90) | 0.88 (0.76, 1.03) | 0.89 (0.79, 0.99) | 1.02 (0.95, 1.10) | 0.99 (0.91, 1.07) | | 0.97 (0.94, 1.01) | | 0.96 (0.88, 1.03) | <0.001 |
| Leucine | 1.02 (0.95, 1.11) | 1.16 (0.98, 1.36) | 0.75 (0.63, 0.89) | 0.87 (0.74, 1.02) | 0.89 (0.79, 1.01) | 1.03 (0.95, 1.11) | 0.99 (0.90, 1.08) | | 0.97 (0.93, 1.01) | | 0.96 (0.88, 1.03) | <0.001 |
| Valine | 1.02 (0.95, 1.11) | 1.15 (0.98, 1.35) | 0.74 (0.62, 0.88) | 0.87 (0.75, 1.02) | 0.89 (0.79, 1.01) | 1.02 (0.94, 1.10) | 0.98 (0.90, 1.07) | | 0.97 (0.93, 1.01) | | 0.95 (0.88, 1.03) | 0.001 |
| Other essential amino acids | 1.02 (0.95, 1.11) | 1.17 (1.00, 1.37) | 0.76 (0.64, 0.90) | 0.88 (0.75, 1.03) | 0.88 (0.78, 1.00) | 1.04 (0.96, 1.12) | 0.99 (0.91, 1.07) | | 0.97 (0.94, 1.01) | | 0.96 (0.88, 1.04) | <0.001 |
| Histidine | 1.02 (0.94, 1.11) | 1.16 (0.98, 1.36) | 0.75 (0.63, 0.88) | 0.89 (0.76, 1.05) | 0.88 (0.77, 1.00) | 1.04 (0.97, 1.13) | 0.98 (0.90, 1.07) | | 0.97 (0.93, 1.01) | | 0.96 (0.87, 1.04) | <0.001 |
| Lysine | 1.03 (0.95, 1.12) | 1.20 (1.02, 1.41) | 0.75 (0.63, 0.90) | 0.89 (0.75, 1.06) | 0.87 (0.76, 1.00) | 1.05 (0.96, 1.14) | 0.99 (0.91, 1.08) | | 0.98 (0.94, 1.02) | | 0.96 (0.88, 1.05) | <0.001 |
| Methionine | 1.03 (0.95, 1.13) | 1.20 (1.01, 1.43) | 0.73 (0.60, 0.89) | 0.87 (0.72, 1.05) | 0.88 (0.77, 1.01) | 1.04 (0.95, 1.13) | 0.98 (0.89, 1.07) | | 0.97 (0.93, 1.01) | | 0.96 (0.87, 1.05) | <0.001 |
| Phenylalanine | 1.02 (0.95, 1.11) | 1.15 (0.96, 1.36) | 0.75 (0.62, 0.89) | 0.86 (0.74, 1.01) | 0.89 (0.78, 1.01) | 1.03 (0.95, 1.11) | 0.99 (0.90, 1.08) | | 0.97 (0.93, 1.01) | | 0.95 (0.87, 1.03) | 0.002 |
| Threonine | 1.02 (0.95, 1.10) | 1.18 (1.02, 1.37) | 0.78 (0.66, 0.92) | 0.87 (0.75, 1.02) | 0.89 (0.79, 1.00) | 1.03 (0.95, 1.11) | 0.99 (0.91, 1.07) | | 0.98 (0.94, 1.01) | | 0.96 (0.89, 1.04) | 0.001 |
| Tryptophan | 1.02 (0.94, 1.11) | 1.19 (1.01, 1.41) | 0.75 (0.63, 0.90) | 0.86 (0.71, 1.02) | 0.88 (0.78, 1.00) | 1.04 (0.95, 1.13) | 0.98 (0.90, 1.08) | | 0.97 (0.93, 1.01) | | 0.96 (0.87, 1.04) | <0.001 |
| Non-essential amino acids | 1.02 (0.94, 1.11) | 1.17 (0.98, 1.39) | 0.75 (0.63, 0.91) | 0.86 (0.73, 1.02) | 0.89 (0.78, 1.02) | 1.03 (0.95, 1.12) | 0.99 (0.90, 1.08) | | 0.97 (0.93, 1.01) | | 0.96 (0.88, 1.03) | 0.003 |
| Alanine | 1.02 (0.94, 1.11) | 1.19 (0.99, 1.42) | 0.80 (0.66, 0.97) | 0.88 (0.73, 1.06) | 0.86 (0.75, 0.99) | 1.03 (0.94, 1.12) | 0.99 (0.90, 1.08) | | 0.97 (0.93, 1.02) | | 0.96 (0.89, 1.04) | 0.01 |
| Arginine | 1.02 (0.94, 1.11) | 1.20 (1.01, 1.42) | 0.82 (0.68, 0.99) | 0.83 (0.70, 0.99) | 0.86 (0.76, 0.98) | 1.04 (0.95, 1.13) | 1.00 (0.92, 1.09) | | 0.98 (0.93, 1.02) | | 0.96 (0.89, 1.04) | 0.004 |
| Aspartic acid | 1.02 (0.95, 1.10) | 1.23 (1.05, 1.44) | 0.81 (0.68, 0.98) | 0.87 (0.74, 1.02) | 0.91 (0.80, 1.02) | 1.04 (0.96, 1.12) | 1.01 (0.93, 1.09) | | 0.99 (0.95, 1.03) | | 0.98 (0.91, 1.05) | 0.006 |
| Cystine | 1.01 (0.90, 1.14) | 1.16 (0.87, 1.54) | 0.70 (0.52, 0.95) | 0.84 (0.64, 1.10) | 0.82 (0.67, 1.01) | 0.98 (0.88, 1.10) | 0.96 (0.84, 1.11) | | 0.94 (0.88, 1.00) | | 0.93 (0.84, 1.01) | 0.09 |
| Glutamic acid | 1.01 (0.91, 1.12) | 1.16 (0.92, 1.47) | 0.69 (0.55, 0.87) | 0.84 (0.69, 1.03) | 0.91 (0.77, 1.08) | 1.03 (0.93, 1.14) | 0.98 (0.87, 1.11) | | 0.96 (0.91, 1.01) | | 0.94 (0.85, 1.04) | 0.005 |
| Glycine | 1.02 (0.93, 1.12) | 1.16 (0.94, 1.43) | 0.83 (0.67, 1.03) | 0.90 (0.74, 1.09) | 0.84 (0.72, 0.98) | 1.02 (0.92, 1.12) | 0.99 (0.89, 1.09) | | 0.97 (0.92, 1.02) | | 0.96 (0.89, 1.03) | 0.08 |
| Hydroxyproline | 1.01 (0.88, 1.15) | 1.09 (0.89, 1.34) | 0.87 (0.63, 1.22) | 1.14 (0.97, 1.35) | 0.82 (0.59, 1.14) | 1.14 (1.01, 1.28) | 1.05 (0.85, 1.29) | | 1.05 (0.98, 1.12) | | 1.05 (0.97, 1.13) | 0.3 |
| Proline | 1.01 (0.90, 1.14) | 1.09 (0.85, 1.40) | 0.66 (0.52, 0.85) | 0.85 (0.69, 1.06) | 0.87 (0.73, 1.05) | 1.01 (0.90, 1.12) | 0.94 (0.81, 1.10) | | 0.93 (0.88, 0.99) | | 0.92 (0.82, 1.02) | 0.01 |
| Serine | 1.03 (0.95, 1.12) | 1.16 (0.97, 1.38) | 0.74 (0.61, 0.90) | 0.85 (0.73, 1.00) | 0.90 (0.78, 1.03) | 1.04 (0.96, 1.12) | 0.99 (0.90, 1.09) | | 0.98 (0.94, 1.02) | | 0.96 (0.87, 1.04) | 0.001 |
| Tyrosine | 1.03 (0.95, 1.12) | 1.17 (0.99, 1.39) | 0.73 (0.60, 0.87) | 0.86 (0.73, 1.02) | 0.90 (0.78, 1.03) | 1.04 (0.96, 1.13) | 0.98 (0.89, 1.08) | | 0.97 (0.93, 1.01) | | 0.96 (0.87, 1.05) | <0.001 |
| Total protein (g/1000 kcal) | 1.02 (0.96, 1.08) | 1.14 (1.02, 1.27) | 0.80 (0.70, 0.91) | 0.91 (0.81, 1.01) | 0.92 (0.84, 1.01) | 1.02 (0.97, 1.08) | 1.02 (0.96, 1.08) | | 0.99 (0.96, 1.02) | | 0.98 (0.91, 1.04) | <0.001 |

1. Hazard ratios modelled per 1 sex-specific SD increment in dietary amino acids or protein. Based on multivariable model stratified by sex and centre, and adjusted for age (continuous), smoking (never, former, current<10, 10-19, 20+ cigarettes/day, unknown), observed alcohol consumption (non-drinkers (<0.1), 0.1-4.9, 5.0-14.9, 15-29.9, 30-59.9, 60+ g/day), physical activity (inactive, moderately inactive, moderately active, active, unknown), employment status (employed or student, neither employed nor student, unknown), highest level of education completed (none or primary, secondary, vocational or university, unknown), history of diabetes (yes, no, unknown), prior hypertension (yes, no , unknown), prior hyperlipidaemia (yes, no, unknown), observed energy intake (continuous), body mass index (<22.5, 22.5-24.9, 25.0-27.4, 27.5-29.9, ≥30.0 kg/m^2^, unknown).
2. Tests of heterogeneity by country from the meta-analysis.

Supplementary table 27: Hazard ratios (95% confidence intervals)^1^ for **ischaemic** (3507 cases) and **haemorrhagic** stroke (1080 cases) by increments of calibrated intakes of dietary amino acids, **excluding the first four years of follow-up**.

| Amino acids (percent of total protein) | Ischaemic stroke | |  | Haemorrhage stroke | |
| --- | --- | --- | --- | --- | --- |
|  | HR (95% CI) | p-trend^2^ |  | HR (95% CI) | p-trend^2^ |
| Branched-chain amino acids | 0.91 (0.86, 0.97) | 0.003 |  | 0.96 (0.86, 1.07) | 0.43 |
| Isoleucine | 0.92 (0.86, 0.97) | 0.005 |  | 0.95 (0.85, 1.07) | 0.40 |
| Leucine | 0.91 (0.85, 0.97) | 0.003 |  | 0.96 (0.85, 1.07) | 0.45 |
| Valine | 0.91 (0.86, 0.96) | 0.001 |  | 0.96 (0.86, 1.07) | 0.45 |
| Other essential amino acids | 0.92 (0.86, 0.98) | 0.008 |  | 0.96 (0.85, 1.08) | 0.49 |
| Histidine | 0.92 (0.86, 0.99) | 0.02 |  | 0.95 (0.84, 1.07) | 0.39 |
| Lysine | 0.92 (0.86, 0.99) | 0.02 |  | 0.97 (0.85, 1.10) | 0.61 |
| Methionine | 0.92 (0.86, 0.98) | 0.02 |  | 0.96 (0.85, 1.09) | 0.54 |
| Phenylalanine | 0.91 (0.85, 0.96) | 0.001 |  | 0.96 (0.85, 1.07) | 0.44 |
| Threonine | 0.92 (0.87, 0.98) | 0.009 |  | 0.95 (0.85, 1.07) | 0.40 |
| Tryptophan | 0.92 (0.87, 0.97) | 0.004 |  | 0.97 (0.87, 1.08) | 0.59 |
| Non-essential amino acids | 0.91 (0.85, 0.97) | 0.003 |  | 0.95 (0.84, 1.07) | 0.40 |
| Alanine | 0.96 (0.90, 1.03) | 0.27 |  | 0.91 (0.80, 1.03) | 0.14 |
| Arginine | 0.97 (0.92, 1.02) | 0.24 |  | 0.91 (0.82, 1.01) | 0.07 |
| Aspartic acid | 0.95 (0.91, 1.00) | 0.05 |  | 0.97 (0.89, 1.06) | 0.52 |
| Cystine | 0.96 (0.88, 1.03) | 0.25 |  | 0.88 (0.76, 1.02) | 0.09 |
| Glutamic acid | 0.87 (0.81, 0.94) | <0.001 |  | 0.96 (0.84, 1.11) | 0.59 |
| Glycine | 1.00 (0.93, 1.07) | 0.94 |  | 0.88 (0.77, 1.01) | 0.07 |
| Hydroxyproline | 1.02 (0.95, 1.10) | 0.60 |  | 1.06 (0.93, 1.21) | 0.41 |
| Proline | 0.86 (0.80, 0.92) | <0.001 |  | 0.97 (0.85, 1.10) | 0.61 |
| Serine | 0.90 (0.85, 0.96) | 0.001 |  | 0.99 (0.89, 1.10) | 0.81 |
| Tyrosine | 0.90 (0.85, 0.96) | 0.001 |  | 0.99 (0.89, 1.10) | 0.85 |
| Total protein (g/1000 kcal) | 0.96 (0.94, 0.99) | 0.02 |  | 0.98 (0.92, 1.03) | 0.42 |

1. Hazard ratios modelled per 1 sex-specific SD increment in dietary amino acids or total protein. Based on multivariable model stratified by sex and centre, and adjusted for age (continuous), calibrated energy intake (continuous), smoking (never, former, current<10, 10-19, 20+ cigarettes/day, unknown), calibrated alcohol consumption (non-drinkers (<0.1), 0.1-4.9, 5.0-14.9, 15-29.9, 30-59.9, 60+ g/day), physical activity (inactive, moderately inactive, moderately active, active, unknown), employment status (employed or student, neither employed nor student, unknown), highest level of education completed (none or primary, secondary, vocational or university, unknown), history of diabetes (yes, no, unknown), prior hypertension (yes, no , unknown), prior hyperlipidaemia (yes, no, unknown), body mass index (<22.5, 22.5-24.9, 25.0-27.4, 27.5-29.9, ≥30.0 kg/m^2^, unknown).
2. Linear trends estimated by including the exposures as continuous variables in the Cox regression.

Supplementary table 28: Hazard ratios (95% confidence intervals)^1^ for **ischaemic** (1966 cases) and **haemorrhagic** stroke (721 cases) by increments of calibrated intakes of dietary amino acids, based on **complete case analyses**.

| Amino acids (percent of total protein) | Ischaemic stroke | |  | Haemorrhage stroke | |
| --- | --- | --- | --- | --- | --- |
|  | HR (95% CI) | p-trend^2^ |  | HR (95% CI) | p-trend^2^ |
| Branched-chain amino acids | 0.91 (0.83, 0.99) | 0.03 |  | 0.99 (0.86, 1.15) | 0.93 |
| Isoleucine | 0.91 (0.83, 0.99) | 0.04 |  | 1.01 (0.87, 1.17) | 0.89 |
| Leucine | 0.91 (0.83, 0.99) | 0.04 |  | 0.99 (0.85, 1.15) | 0.88 |
| Valine | 0.91 (0.83, 0.99) | 0.02 |  | 0.99 (0.86, 1.14) | 0.88 |
| Other essential amino acids | 0.92 (0.83, 1.01) | 0.07 |  | 1.01 (0.87, 1.19) | 0.85 |
| Histidine | 0.92 (0.84, 1.02) | 0.10 |  | 1.00 (0.85, 1.17) | 1.00 |
| Lysine | 0.92 (0.83, 1.02) | 0.11 |  | 1.04 (0.89, 1.23) | 0.61 |
| Methionine | 0.91 (0.82, 1.01) | 0.07 |  | 1.03 (0.87, 1.22) | 0.71 |
| Phenylalanine | 0.91 (0.83, 0.99) | 0.04 |  | 0.98 (0.84, 1.13) | 0.75 |
| Threonine | 0.92 (0.84, 1.01) | 0.07 |  | 1.02 (0.88, 1.18) | 0.82 |
| Tryptophan | 0.92 (0.85, 1.00) | 0.06 |  | 1.00 (0.86, 1.15) | 0.95 |
| Non-essential amino acids | 0.91 (0.82, 1.00) | 0.05 |  | 1.00 (0.85, 1.18) | 0.98 |
| Alanine | 0.95 (0.85, 1.05) | 0.31 |  | 1.04 (0.88, 1.24) | 0.62 |
| Arginine | 0.97 (0.89, 1.05) | 0.44 |  | 1.00 (0.87, 1.15) | 0.97 |
| Aspartic acid | 0.96 (0.89, 1.03) | 0.26 |  | 1.05 (0.93, 1.18) | 0.42 |
| Cystine | 0.97 (0.86, 1.08) | 0.54 |  | 1.01 (0.83, 1.23) | 0.89 |
| Glutamic acid | 0.87 (0.78, 0.97) | 0.02 |  | 0.97 (0.80, 1.16) | 0.72 |
| Glycine | 1.00 (0.90, 1.11) | 0.98 |  | 1.05 (0.88, 1.25) | 0.58 |
| Hydroxyproline | 0.98 (0.89, 1.08) | 0.66 |  | 1.17 (1.01, 1.35) | 0.04 |
| Proline | 0.85 (0.77, 0.94) | 0.002 |  | 0.95 (0.81, 1.13) | 0.57 |
| Serine | 0.91 (0.84, 0.99) | 0.04 |  | 1.00 (0.87, 1.15) | 0.98 |
| Tyrosine | 0.90 (0.83, 0.99) | 0.02 |  | 0.99 (0.86, 1.14) | 0.92 |
| Total protein (g/1000 kcal) | 0.96 (0.92, 1.01) | 0.09 |  | 1.01 (0.93, 1.09) | 0.83 |

1. Hazard ratios modelled per 1 sex-specific SD increment in dietary amino acids or total protein. Based on multivariable model stratified by sex and centre, and adjusted for age (continuous), calibrated energy intake (continuous), smoking (never, former, current<10, 10-19, 20+ cigarettes/day, unknown), calibrated alcohol consumption (non-drinkers (<0.1), 0.1-4.9, 5.0-14.9, 15-29.9, 30-59.9, 60+ g/day), physical activity (inactive, moderately inactive, moderately active, active, unknown), employment status (employed or student, neither employed nor student, unknown), highest level of education completed (none or primary, secondary, vocational or university, unknown), history of diabetes (yes, no, unknown), prior hypertension (yes, no , unknown), prior hyperlipidaemia (yes, no, unknown), body mass index (<22.5, 22.5-24.9, 25.0-27.4, 27.5-29.9, ≥30.0 kg/m^2^, unknown).
2. Linear trends estimated by including the exposures as continuous variables in the Cox regression.

518,502 participants recruited from 10 countries

Excluded the following countries:

- France ( n=74,472)
- Greece (n=27,539)
- Norway (n=37,200)

379,291 participants
from 7 countries

Exclusions based on the following reasons:

- Known or possible history of myocardial infarction or stroke or had unknown prior disease status at recruitment (n=9,803)
- Case event date after the centre-specific censoring date (n=16)
- No follow-up information (n=10)
- No dietary data (n=5,995)
- Missing data on all major covariates (n=60)
- Extreme energy intake defined as <800 kcal/d or >4200 kcal/d in men and <500 kcal/d or >3500 kcal/d in women (n=7,265)

356,142 participants

Supplementary figure 1: Participant flow chart of the study

Further details on the exclusions can be found in the Supplementary methods.

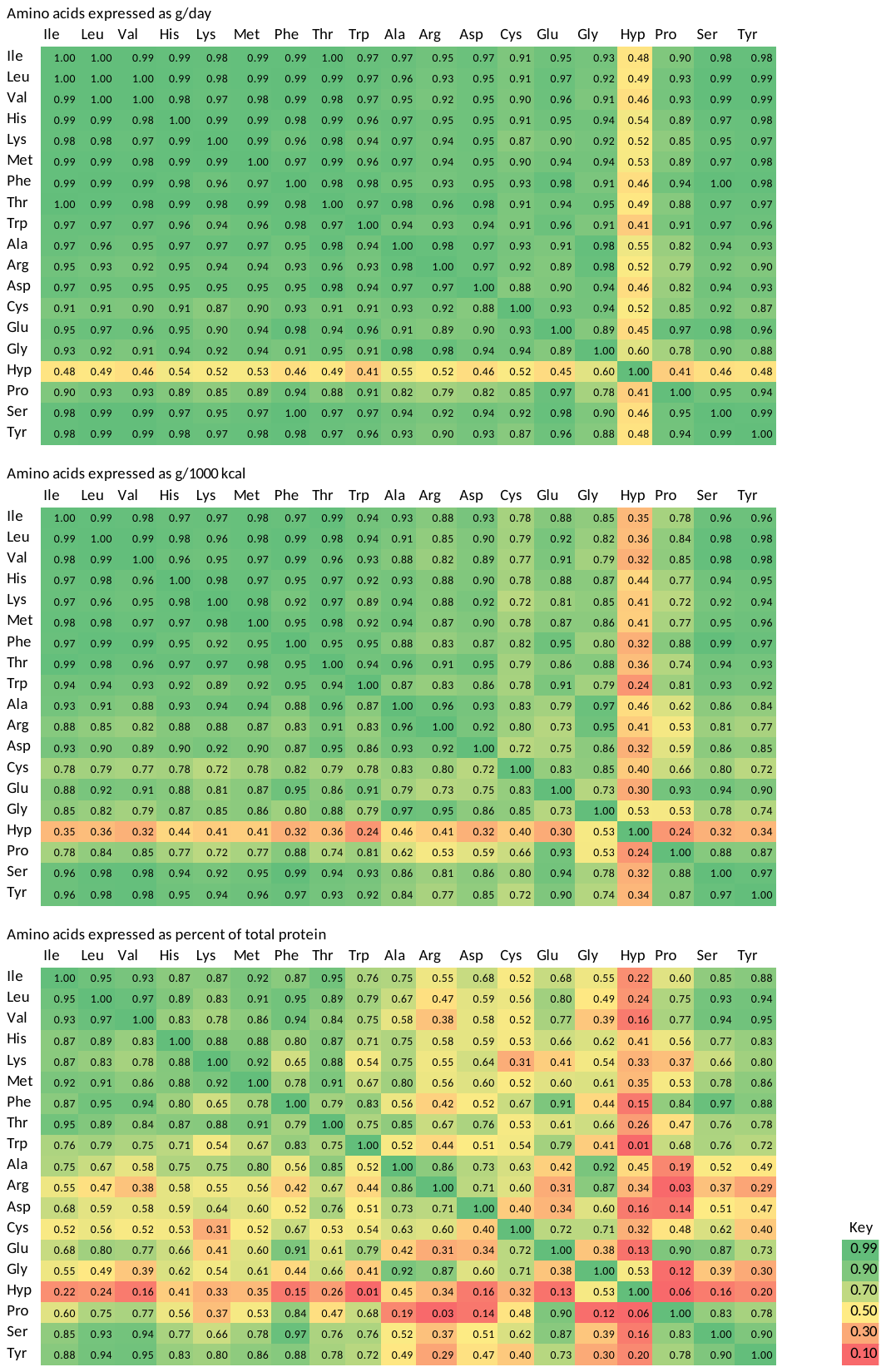

Supplementary figure 2: Spearman correlation coefficients **between individual dietary amino acids**, expressed as g/day, g/1000kcal and percent of total protein

Ile= isoleucine, Leu=leucine, Val=valine, His=histidine, Lys=lysine, Met=Methionine, Phe=phenylalanine, Thr=threonine, Trp=tryptophan, Ala=alanine, Arg=arginine, Asp=aspartic acid, Cys=cystine, Glu=glutamic acid, Gly=glycine, Hyp=hydroxyproline, Pro=proline, Ser=serine, Tyr=tyrosine

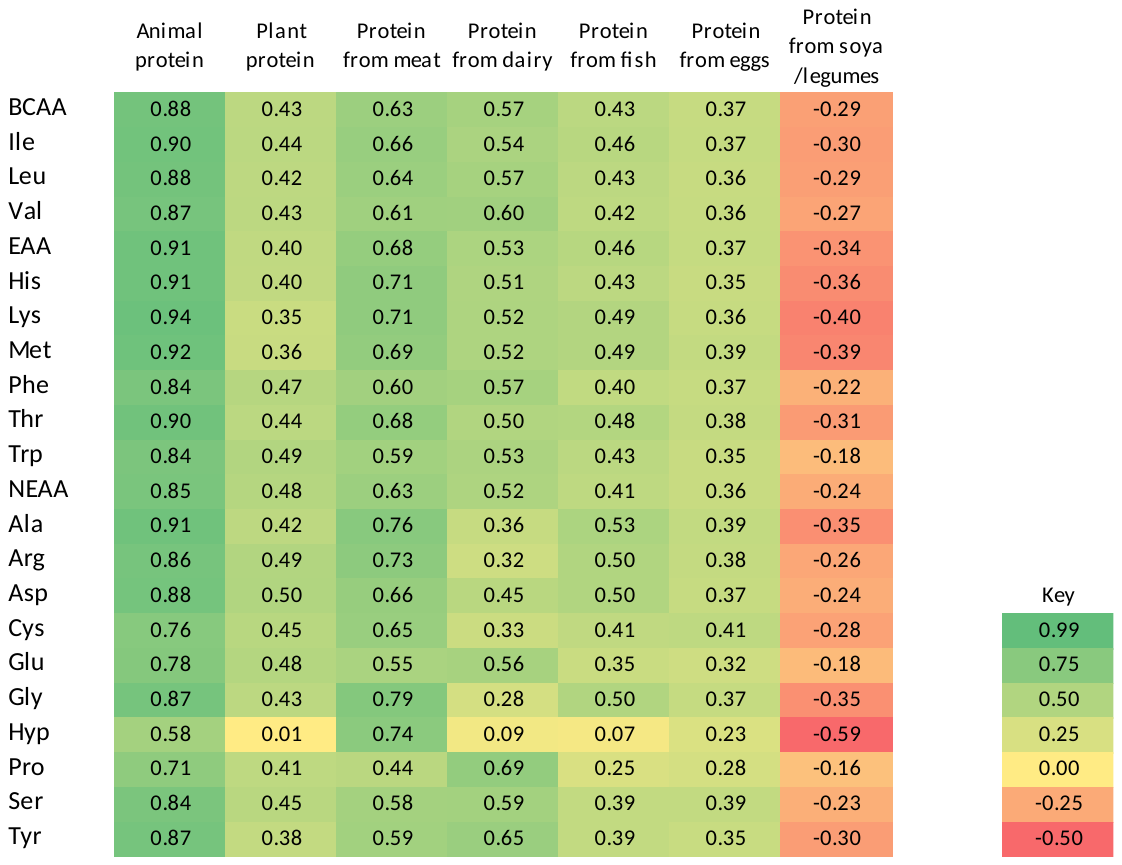

Supplementary figure 3: Spearman correlation coefficients of dietary amino acids (g/day) with **dietary protein from different sources**

BCAA= branched-chain amino acids, Ile= isoleucine, Leu=leucine, Val=valine, EAA=other essential amino acids, His=histidine, Lys=lysine, Met=Methionine, Phe=phenylalanine, Thr=threonine, Trp=tryptophan, NEAA= non-essential amino acids, Ala=alanine, Arg=arginine, Asp=aspartic acid, Cys=cystine, Glu=glutamic acid, Gly=glycine, Hyp=hydroxyproline, Pro=proline, Ser=serine, Tyr=tyrosine

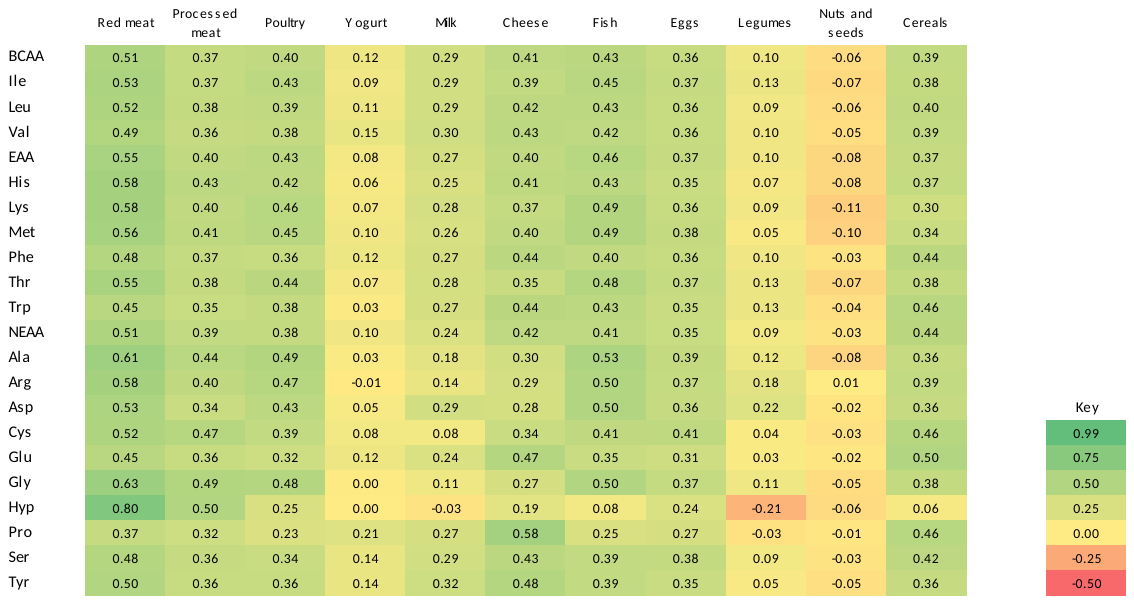

Supplementary figure 4: Spearman correlation coefficients of dietary amino acids (g/day) with **food sources of protein**

BCAA= branched-chain amino acids, Ile= isoleucine, Leu=leucine, Val=valine, EAA=other essential amino acids, His=histidine, Lys=lysine, Met=Methionine, Phe=phenylalanine, Thr=threonine, Trp=tryptophan, NEAA= non-essential amino acids, Ala=alanine, Arg=arginine, Asp=aspartic acid, Cys=cystine, Glu=glutamic acid, Gly=glycine, Hyp=hydroxyproline, Pro=proline, Ser=serine, Tyr=tyrosine

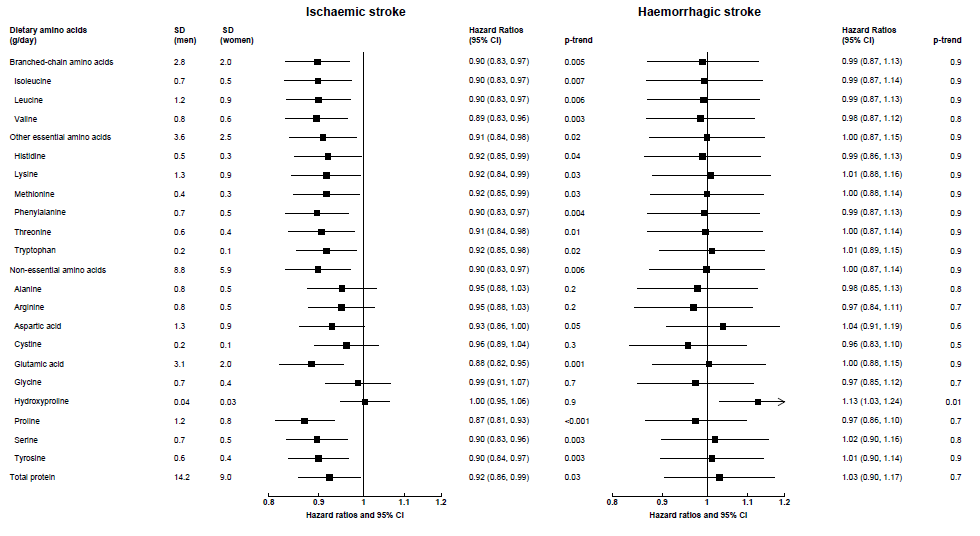

Supplementary figure 5 Hazard ratios (95% confidence intervals) for **ischaemic** (4295 cases) and **haemorrhagic** stroke (1375 cases) by increments of **calibrated** intakes of dietary amino acids (**as g/day**).

Hazard ratios modelled per 1 sex-specific SD increment in dietary amino acids, expressed as grammes per day. The model was stratified by sex and centre, and adjusted for age (continuous), calibrated energy intake (continuous), smoking (never, former, current<10, 10-19, 20+ cigarettes/day, unknown), calibrated alcohol consumption (non-drinkers (<0.1), 0.1-4.9, 5.0-14.9, 15-29.9, 30-59.9, 60+ g/day), physical activity (inactive, moderately inactive, moderately active, active, unknown), employment status (employed or student, neither employed nor student, unknown), highest level of education completed (none or primary, secondary, vocational or university, unknown), history of diabetes (yes, no, unknown), prior hypertension (yes, no , unknown), prior hyperlipidaemia (yes, no, unknown), body mass index (<22.5, 22.5-24.9, 25.0-27.4, 27.5-29.9, ≥30.0 kg/m^2^, unknown).

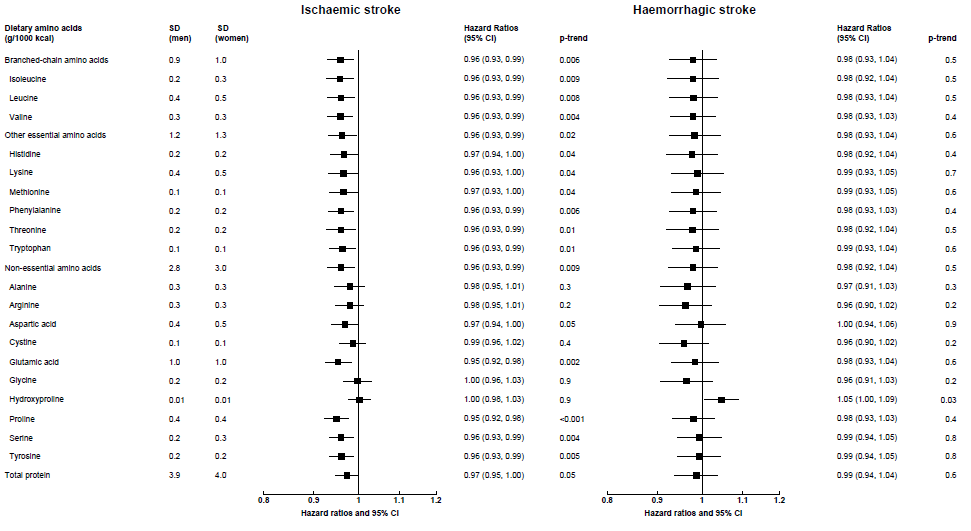

Supplementary figure 6 Hazard ratios (95% confidence intervals) for **ischaemic** (4295 cases) and **haemorrhagic** stroke (1375 cases) by increments of **calibrated** intakes of dietary amino acids (**as g/1000 kcal**).

Hazard ratios modelled per 1 sex-specific SD increment in dietary amino acids, expressed as grammes per 1000 kcal. The model was stratified by sex and centre, and adjusted for age (continuous), calibrated energy intake (continuous), smoking (never, former, current<10, 10-19, 20+ cigarettes/day, unknown), calibrated alcohol consumption (non-drinkers (<0.1), 0.1-4.9, 5.0-14.9, 15-29.9, 30-59.9, 60+ g/day), physical activity (inactive, moderately inactive, moderately active, active, unknown), employment status (employed or student, neither employed nor student, unknown), highest level of education completed (none or primary, secondary, vocational or university, unknown), history of diabetes (yes, no, unknown), prior hypertension (yes, no , unknown), prior hyperlipidaemia (yes, no, unknown), body mass index (<22.5, 22.5-24.9, 25.0-27.4, 27.5-29.9, ≥30.0 kg/m^2^, unknown).

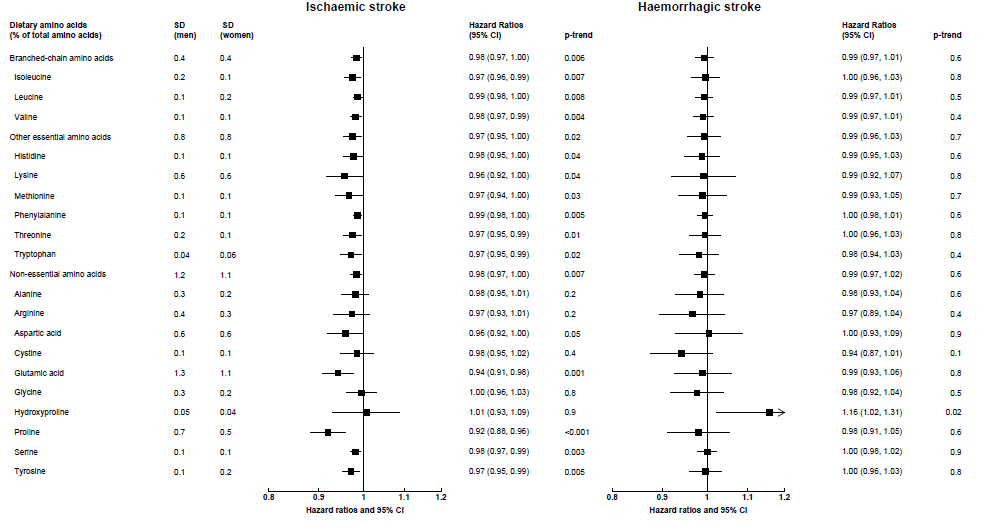

Supplementary figure 7 Hazard ratios (95% confidence intervals) for **ischaemic** (4295 cases) and **haemorrhagic** stroke (1375 cases) by increments of **calibrated** intakes of dietary amino acids (**as percentage of the sum of all available amino acids**).

Hazard ratios modelled per 1 sex-specific SD increment in dietary amino acids, expressed as percent of the sum of all amino acids. The model was stratified by sex and centre, and adjusted for age (continuous), calibrated energy intake (continuous), smoking (never, former, current<10, 10-19, 20+ cigarettes/day, unknown), calibrated alcohol consumption (non-drinkers (<0.1), 0.1-4.9, 5.0-14.9, 15-29.9, 30-59.9, 60+ g/day), physical activity (inactive, moderately inactive, moderately active, active, unknown), employment status (employed or student, neither employed nor student, unknown), highest level of education completed (none or primary, secondary, vocational or university, unknown), history of diabetes (yes, no, unknown), prior hypertension (yes, no , unknown), prior hyperlipidaemia (yes, no, unknown), body mass index (<22.5, 22.5-24.9, 25.0-27.4, 27.5-29.9, ≥30.0 kg/m^2^, unknown).

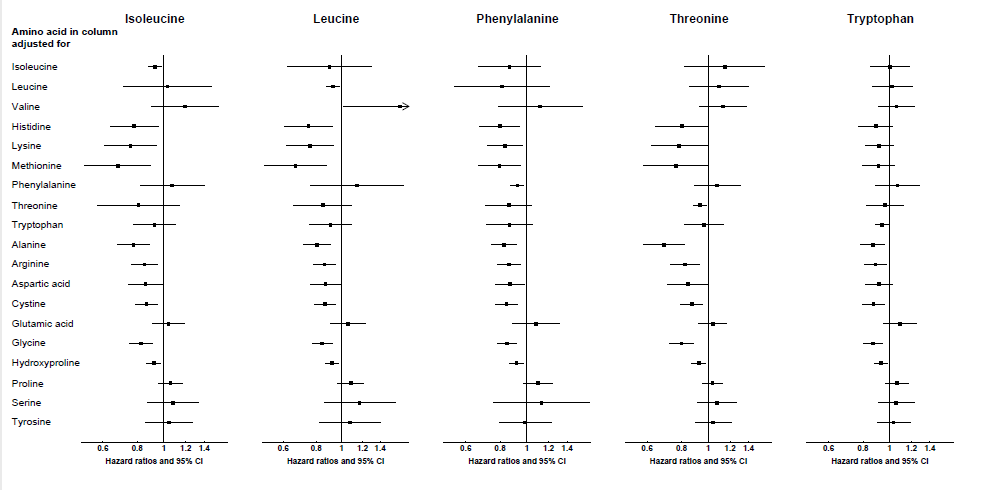

Supplementary figure 8 Hazard ratios (95% confidence intervals) for the association between selected amino acids (isoleucine, leucine, phenylalanine, threonine, tryptophan) and **ischaemic** stroke (4295 cases), **with mutual adjustment for other amino acids**.

Results are shown for amino acids which were statistically significantly associated with ischemic stroke in both calibrated analyses only, but not in the observed analyses. Hazard ratios modelled per 1 sex-specific SD (as shown in Figure 1 and Supplementary table 2) increment in dietary amino acids in column heading, expressed as percent of total protein, with multivariable adjustment plus amino acid on the left panel. If the amino acid in column and in left panel are the same, the result is interpretable as the hazard ratio for the amino acid based on the multivariable adjusted model. The multivariable model was stratified by sex and centre, and adjusted for age (continuous), calibrated energy intake (continuous), smoking (never, former, current<10, 10-19, 20+ cigarettes/day, unknown), calibrated alcohol consumption (non-drinkers (<0.1), 0.1-4.9, 5.0-14.9, 15-29.9, 30-59.9, 60+ g/day), physical activity (inactive, moderately inactive, moderately active, active, unknown), employment status (employed or student, neither employed nor student, unknown), highest level of education completed (none or primary, secondary, vocational or university, unknown), history of diabetes (yes, no, unknown), prior hypertension (yes, no , unknown), prior hyperlipidaemia (yes, no, unknown), body mass index (<22.5, 22.5-24.9, 25.0-27.4, 27.5-29.9, ≥30.0 kg/m^2^, unknown).
